# Subnational HIV incidence trends in Malawi: large, heterogeneous declines across space

**DOI:** 10.1101/2023.02.02.23285334

**Authors:** Timothy M Wolock, Seth Flaxman, Tiwonge Chimpandule, Stone Mbiriyawanda, Andreas Jahn, Rose Nyirenda, Jeffrey W Eaton

## Abstract

The rate of new HIV infections globally has decreased substantially from its peak in the late 1990s, but the epidemic persists and remains highest in many countries in eastern and southern Africa. Previous research hypothesised that, as the epidemic recedes, it will become increasingly concentrated among sub-populations and geographic areas where transmission is the highest and that are least effectively reached by treatment and prevention services. However, empirical data on subnational HIV incidence trends is sparse, and the local transmission rates in the context of effective treatment scale-up are unknown. In this work, we developed a novel Bayesian spatio-temporal epidemic model to estimate adult HIV prevalence, incidence and treatment coverage at the district level in Malawi from 2010 through the end of 2021. We found that HIV incidence decreased in every district of Malawi between 2010 and 2021 but the rate of decline varied by area. National-level treatment coverage more than tripled between 2010 and 2021 and more than doubled in every district. Large increases in treatment coverage were associated with declines in HIV transmission, with 12 districts having incidence-prevalence ratios of 0.03 or less (a previously suggested threshold for epidemic control). Across districts, incidence varied more than HIV prevalence and ART coverage, suggesting that the epidemic is becoming increasingly spatially concentrated. Our results highlight the success of the Malawi HIV treatment programme over the past decade, with large improvements in treatment coverage leading to commensurate declines in incidence. More broadly, we demonstrate the utility of spatially resolved HIV modelling in generalized epidemic settings. By estimating temporal changes in key epidemic indicators at a relatively fine spatial resolution, we were able to directly assess, for the first time, whether the ART scaleup in Malawi resulted in spatial gaps or hotspots. Regular use of this type of analysis will allow HIV program managers to monitor the equity of their treatment and prevention programmes and their subnational progress towards epidemic control.

## Introduction

Globally, new human immunodeficiency virus (HIV) infections have decreased from a peak of 3.0 million in 1997 to 1.5 million in 2020, while AIDS deaths decreased decreased from 1.9 million in 2004 to 680,000 in 2020.^1^ These improvements have resulted, in large part, from the rapid scale-up of life-saving antiretroviral therapy (ART), alongside combination HIV prevention.^2–4^ Although HIV elimination will only be attained in future generations, measuring progress towards epidemic transition, in which low levels of acquired immunodeficiency syndrome (AIDS) mortality are maintained and new infections are continually reduced, is critical for guiding policy-making at global and local levels.^5,6^ The goal of epidemic transition has been encoded into international policy in the UNAIDS Fast Track Strategy, which calls for a 90% reduction in new infections from 2010 levels by 2030.^7^

The most rapid progress in reducing new HIV infections and improving ART coverage has been in Eastern and Southern Africa (ESA), where burden and programme investments have historically been highest. Over the past 25 years, annual new HIV infections have more than halved from 1.6 million in 1997 to 670,000 in 2021.^1^ However, the region remains disproportionately affected: those 670,000 new infections accounted for 45% of all global infections in 2021.

The rate of new HIV infection in a population, or *incidence*, reflects the epidemic’s trajectory in the short- and long-term, making it the most important metric for measuring progress towards epidemic transition.^6,8,9^ For example, high incidence today will necessitate sustained long-term treatment provision far into the future due to the need for lifelong ART. Estimating incidence has therefore been a central task of HIV epidemiology since the beginning of the epidemic. A 2017 UNAIDS consultation recommended several “HIV epidemic transition” metrics to assess whether HIV programmes are on track towards ending the AIDS epidemic.^6^ In addition to the rate of new infections and percent reduction in new infections, the leading recommended indicator was the incidence-to-prevalence ratio (IPR) with target threshold of 0.03, below which the epidemic is on a long-term trajectory of decline. National estimates of IPR have since been regularly reported by UNAIDS and others.^1,10^

However, epidemic transition has not been well-quantified at subnational areas, where HIV dynamics are highly heterogeneous.^11,12^ Sustaining declining incidence will require granular information about where ongoing HIV transmission and new infections occur. Previous work has hypothesized that as the epidemic recedes, incidence will become increasingly concentrated in more vulnerable populations and geographic areas.^13–15^ Areas with persistent high prevalence could become “sources” of new infections that sustain epidemics in otherwise well-managed “sink” areas.^16^ Such dynamics could stall or even reverse progress.

### Estimating HIV incidence

Despite its importance as an epidemiological indicator, population-level HIV incidence is difficult to measure directly, even at the national level.^8,17^ Long survival following HIV infection (fifteen years or more untreated and 30 years or longer with ART) means that prevalent cases or new diagnoses may represent individuals infected many years ago.^18,19^ Therefore, HIV diagnoses data provide little direct information on trends in new infections. In settings with complete HIV case reporting, back-calculation methods can be used to estimate incidence from new diagnoses, but case reporting is only partially implemented in most high HIV-burden settings^20–22^ Additionally, health data systems currently struggle to distinguish new diagnoses from repeat diagnoses, rendering existing back-calculation approaches unsuitable.^23^ Recent biomarker-based algorithms have been deployed to identify recently infected individuals (typically in the previous 4-6 months), providing cross-sectional estimates of HIV incidence in national surveys.^24,25^ However, national surveys are infrequent and require prohibitively large sample sizes to provide reliable estimates of trends in incidence.^26^

Instead, in high HIV burden settings, incidence estimation has relied on fitting mathematical models to data measuring trends in HIV prevalence from national household surveys and from antenatal care (ANC) surveillance systems.^27–29^ These models infer incidence trends consistent with observed prevalence trajectories by combining assumptions about HIV transmission dynamics and survival after infection with and without ART. However, these models assume both statistical and epidemiological independence across regions, making them inappropriate for subnational estimation. Such independence means that HIV transmission dynamics are assumed to not vary systematically over space and that spatial treatment seeking dynamics cannot be accounted for.^30^

Other recent research has focused on quantifying spatial burden of HIV prevalence and ART coverage using spatial smoothing, small-area estimation, Bayesian geostatistical, and machine learning approaches.^11,12,30,31^ Less research has addressed subnational incidence estimation. The Naomi model predicts subnational incidence alongside prevalence and ART coverage but does not estimate trends.^30^ Sartorius et al. fit a compartmental epidemic model to predicted subnational HIV prevalence trends but included incomplete subnational HIV treatment data and did not consider spatial structure in infection dynamics.^12,32^

In this work, we developed a spatio-temporal epidemic model that bridges the gap between spatially resolved models of prevalence and national-level models of incidence. Our model simultaneously infers HIV prevalence, incidence, and treatment coverage by subnational region, sex, and time by fitting spatio-temporally varying HIV transmission and treatment initiation rates within an epidemic model to data from household surveys, ANC facilities, and ART programmes.

### HIV in Malawi

We used our model to estimate district-level HIV prevalence, incidence, and treatment coverage in Malawi from 2010 through 2021. Malawi is a country in Southern Africa with population around 20 million people.^33^ It consists of 28 districts, each having an average population of slightly more than 700,000 people. Its total area of approximately 100,000 squared kilometres makes it one of the smallest countries in the ESA region.

Malawi has experienced a severe HIV epidemic over the past 40 years, similar to nearby countries in the ESA region. Incidence among adults aged 15-49 peaked at 22 new infections per 1,000 people in 1993, and HIV prevalence among adults remains among the highest in the world at 8%.^3^

High national-level prevalence in Malawi masks dramatic subnational spatial variation. UNAIDS estimated that in 2021, district-level adult HIV prevalence ranged from 3% to 17% across districts.^34^ The epidemic disproportionately affects the south of country, which is more densely populated than the north. Even within small regions, urban areas exhibit much higher prevalence than surrounding rural areas.

Despite high HIV burden and health system constraints, Malawi has built one of the most successful HIV treatment programmes in the world by implementing a public health approach to scaling up treatment that focuses on ensuring equitable access to ART across the country.^35–38^ A recent household survey estimated that 87% of adults with HIV were virally suppressed, indicating successful treatment.^39^ Programmatic success has been underpinned by robust, standardised data collection through which HIV testing and ART provision is systematically reported to central health authorities on a quarterly basis.

The confluence of high-quality data, previously well documented spatial variation, and local demand for district-level burden estimation made Malawi an ideal setting in which to develop and demonstrate our model. We used these estimates to quantify district-level progress towards the target incidence-prevalence ratio of 0.03, as well as the incidence thresholds proposed by Galvani et al.^9^ Finally, we investigated whether large improvements in treatment coverage between 2010 and 2021 resulted in spatially equitable changes in district-level HIV incidence and whether, consistent with the “source-sink” theories described above, the epidemic in Malawi was becoming more spatially concentrated.

## Results

### District-level HIV data

Subnational HIV data in Malawi consisted of (1) cross-sectional measures of adult HIV prevalence from four nationally-representative household surveys conducted between 2004 and 2016, with cross-sectional ART coverage and proportion recently infected in the 2015-2016 Malawi PHIA (MPHIA), (2) routinely collected health system data on the HIV status of pregnant women attending ANC services each quarter between 2011 and 2021, and (3) data on the number of patients accessing ART the end of each quarter between 2004 and 2021.^40–43^

Nationally, HIV prevalence among adults aged 15-49 years was estimated as 10.0% (9.2% to 10.8%) in 2015-2016 from the MPHIA and 9.0% (8.2% to 10.0%) from the 2015-16 Malawi Demographic and Household Surveys (MDHS), a large decline from 10.6% (9.7% to 11.6%) in 2004-2005.^40–43^ Declining prevalence was corroborated by HIV prevalence among pregnant women, which declined from 8.5% in 2011 to 6.3% in 2021. ART coverage was 68.6% (65.9% to 71.2%) in the 2015-2016 MPHIA survey, reflecting the dramatic scale-up in treatment since the programme’s start in the early 2000s. Between 2010 and 2021, the number of people receiving ART in Malawi increased nearly four-fold from 247,100 to 895,100.

The high burden and high treatment coverage at national level masks dramatic subnational variation. Prevalence in the Southern region is more than twice that in the Central and Northern regions, at 15.3% (14.1% to 16.6%), 6.0% (5.2% to 6.9%), and 6.8% (5.4% to 8.6%), respectively, in the 2015-2016 MPHIA survey. Across districts, prevalence in the survey ranged from 20.3% (14.4% to 27.8%) in Phalombe to 2.4% (0.7% to 8.0%) in Ntchisi, while ART coverage ranged from 89.8% (58.7% to 98.2%) in Mwanza to 47.7% (29.9% to 66.2%) in Dowa. HIV prevalence among pregnant women at ANC corroborated this wide variation, ranging from 11.1% in Mulanje to 1.9% in Ntchisi. However, between 2011 to 2021, prevalence declined consistently across all 28 districts by a median of 28% (interquartile range [IQR] 24% to 30%). The number of patients accessing ART increased between 2010 to 2021 by a median of 267% (IQR 215% to 311%).

### Model fit and model selection

We used a cross-validation strategy to evaluate potential specifications for the modelling the HIV transmission rate over time. Among 146 combinations considered, no single specification clearly fit better to the data than all others. In general, the best fitting models used five-year spaced spline knots with first-order autoregressive priors in the transmission rate model. The results presented here were generated using a B-spline of order two with autoregressive priors on the first differences between the coefficients (Supplemental Material Sections 1.5 and 2).

Figure 1 presents an example of the model fit to data about multiple outcomes from 1995 through 2021 for Blantyre, a densely populated high-prevalence district in southern Malawi. HIV prevalence in Blantyre declined among both women and men across the four household surveys, and HIV prevalence among pregnant women declined steadily over the whole period. Since ART programme inception in 2005, the number of adults 15-49 receiving ART in Blantyre increased to 79,000, and 92% (86% to 97%) and 80% (72% to 88%) of women and men, respectively, with HIV were on ART by 2021.

**Figure 1:**
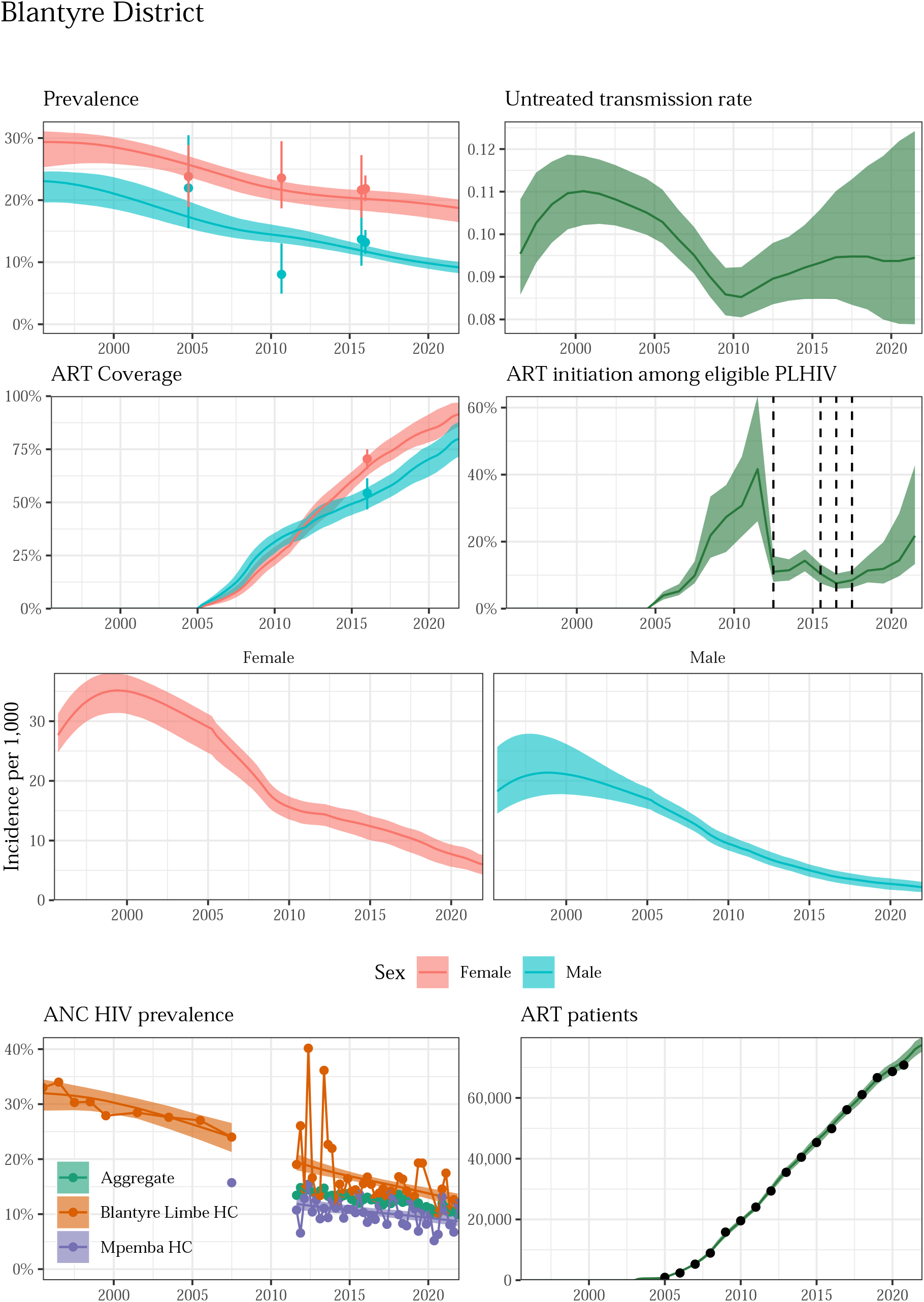
Model fit to HIV data sources in Blantyre District, 1995-2021. Estimated prevalence, ART coverage, untreated transmission rates, annual ART initiation probabilities, ANC prevalence, and ART patient counts in the Blantyre district in southern Malawi with household survey data (HIV prevalence and ART coverage), HIV prevalence among pregnant women attending ANC facilities, and the number of adults 15-49 receiving ART programmatic reporting data (points). Prevalence, ART coverage, incidence rate, and ART patients reflect adults aged 15-49 years. Vertical dashed lines indicate years of ART eligibility changes. Different colours on panel “ANC prevalence” indicate different ANC facilities.

These changes in prevalence and ART coverage resulted from steeply and steadily declining HIV incidence from 2000 to present. During this period, the HIV transmission rate by untreated adults with HIV was stable: 0.11 (0.10 to 0.11) in 2000 and 0.10 (0.09 to 0.11) in 2021. Infectious men transmitted HIV at a 3.9 (2.5 to 6.2) times higher rate than infectious women. The annual probability of ART initiation for an untreated adult reached 21.3% (16.2% to 39.4%) in 2021. Similarly good fits were obtained in all 28 districts (Supplemental Figures 5-32).

### National-level estimates

Aggregating over all districts, at the end of 2021, 7.9% (7.6% to 8.2%) of adults aged 15-49 years in Malawi were living with HIV, of whom 88% (86% to 93%) were on ART. The HIV incidence rate was 2.3 (1.7 to 2.7) new infections per 1,000 at risk. Between 2010 and 2021, HIV prevalence decreased by 25% (22% to 29%) and incidence decreased by 69% (64% to 76%), while ART coverage increased from 26% (26% to 27%) to 88% (86% to 93%), a 3.3 (3.2 to 3.6) times increase.

HIV prevalence among women aged 15-49 in 2021 was 10.4% (10.0% to 10.8%), twice as high as 5.1% (4.7% to 5.7%) among men. ART coverage was also higher among women at 91% (89% to 95%), compared to 81% (76% to 88%) among men. Incidence was 2.5 (1.7 to 3.6) times higher among women than in men: 3.2 (2.6 to 3.8) new infections per 1,000 women compared to 1.3 (0.8 to 1.8) per 1,000 men. For comparison, UNAIDS estimated incidence rates of 2.4 and 1.4 among Malawian women and men, respectively, in 2021.^3^

### Subnational estimates

Across 28 districts of Malawi, median prevalence was 7.1% (6.7% to 7.5%) in 2021. Prevalence ranged from 15.6% (14.3% to 17.4%) in Mulanje in south-east Malawi to 2.0% (1.6% to 2.6%) in Ntchisi in central Malawi (Figure 2, Table 1). Median ART coverage was 91% (87% to 95%) and ranged from 96% (84% to 98%) in Phalombe to 53% (43% to 71%) in Nsanje. Median HIV incidence across districts was 1.9 (1.5 to 2.2) new infections per 1,000 people but varied across district. Incidence was highest in Mulanje at 5.2 (3.7 to 7.2) new infections per 1,000 and lowest in Ntchisi at 0.4 (0.2 to 0.9) (Table 1).

**Figure 2:**
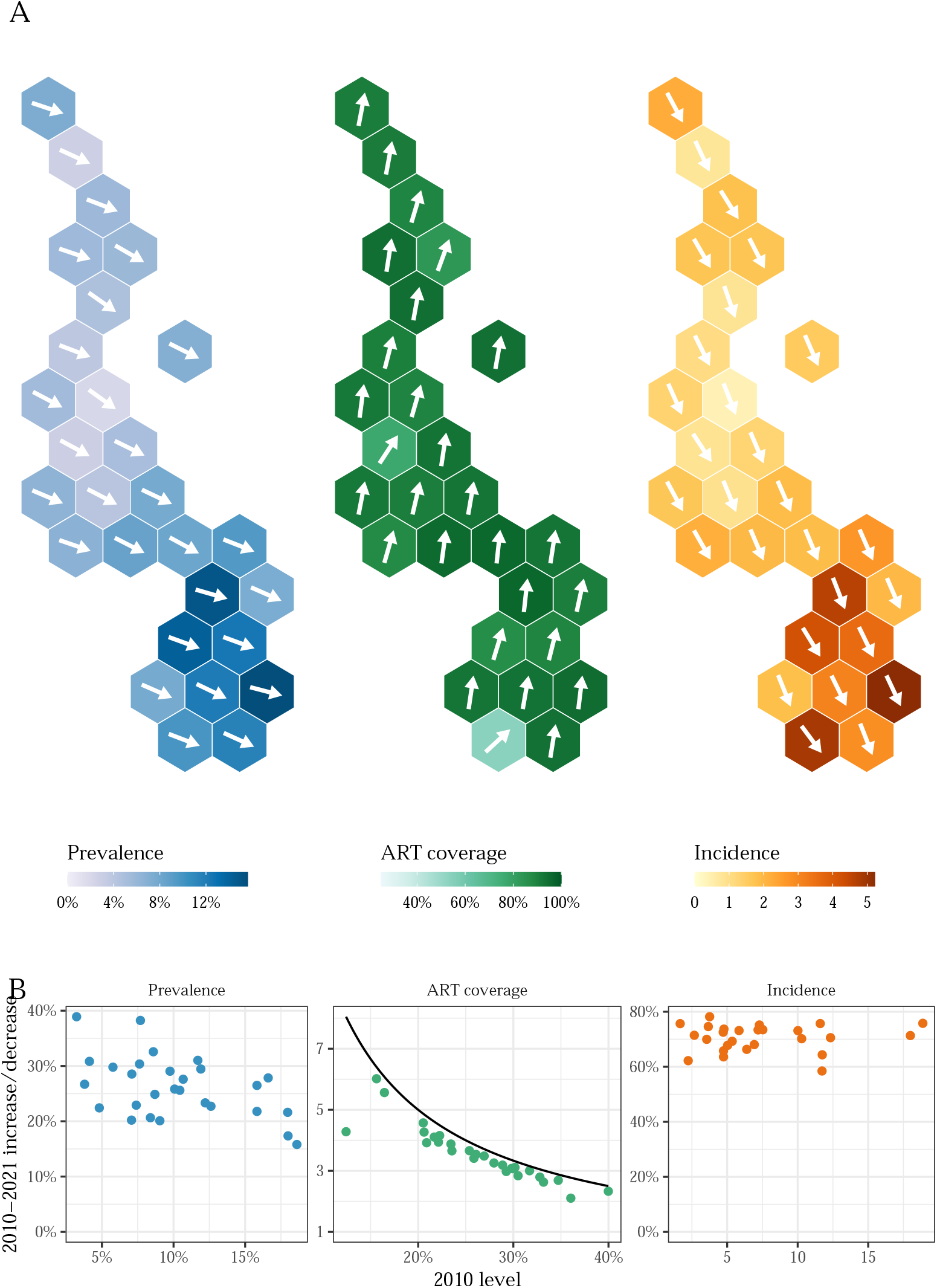
Trends and levels of HIV in Malawi, 2010-2021. A) Hexagonal tile maps present district-level HIV prevalence, ART coverage, and HIV incidence among adults aged 15-49 in Malawi in 2021. The angle of each arrow corresponds to the district-level percent change in each indicator relative to the theoretical maximum change from the 2010 baseline. Upward and downward pointing arrows indicate increases and decreases, respectively. The theoretical maximum change in prevalence and incidence is a 100% decrease, and the maximum change in ART coverage is the percent change needed to reach 100% coverage from the 2010 level. B) Scatter plots comparing the level of each indicator in 2010 to change between 2010 and 2021. Change is *x*-fold increase for ART coverage and percent decrease for prevalence and incidence.

**Table 1:** Estimated national and district-level HIV prevalence, ART coverage, and HIV incidence in Malawi in 2021 and percent changes between 2010 and 2021. Point estimates are posterior medians, and parenthetical estimates are 95% credible intervals.

|  | Prevalence |  | ART Coverage |  | Incidence |  |
| --- | --- | --- | --- | --- | --- | --- |
|  | 2021 value | 2010-2021 decrease | 2021 value | 2010-2021 increase | 2021 value | 2010-2021 decrease |
| National | 8% (8%-8%) | 25% (22%-29%) | 88% (86%-93%) | 3.3 (3.2-3.6) | 2.3 (1.7-2.7) | 69% (64%-76%) |
| **Northern** |  |  |  |  |  |  |
| Chitipa | 3% (2%-4%) | 27% (15%-40%) | 91% (75%-96%) | 3.9 (2.6-6.5) | 0.8 (0.4-1.3) | 71% (59%-80%) |
| Karonga | 7% (7%-8%) | 20% (11%-27%) | 91% (83%-97%) | 3.3 (2.8-3.8) | 2.2 (1.5-3.2) | 68% (58%-76%) |
| Likoma | 7% (6%-9%) | 29% (20%-34%) | 94% (79%-98%) | 3.1 (2.7-3.7) | 1.5 (0.9-3.0) | 73% (61%-80%) |
| Mzimba | 6% (5%-6%) | 20% (13%-29%) | 94% (85%-97%) | 2.7 (2.4-3.1) | 1.7 (1.0-2.6) | 66% (49%-75%) |
| Nkhata Bay | 6% (5%-7%) | 33% (26%-39%) | 83% (69%-94%) | 3.9 (3.1-5.3) | 1.7 (0.9-2.6) | 69% (59%-82%) |
| Rumphi | 6% (4%-7%) | 23% (14%-30%) | 89% (73%-97%) | 2.6 (2.2-3.2) | 1.8 (1.0-2.8) | 64% (46%-72%) |
| **Central** |  |  |  |  |  |  |
| Dedza | 4% (3%-5%) | 30% (24%-40%) | 90% (74%-96%) | 4.3 (3.5-6.3) | 0.9 (0.5-1.7) | 75% (61%-83%) |
| Dowa | 3% (2%-4%) | 31% (23%-40%) | 77% (56%-89%) | 2.1 (1.6-2.7) | 0.8 (0.5-1.5) | 62% (43%-74%) |
| Kasungu | 4% (3%-5%) | 22% (14%-33%) | 90% (76%-96%) | 3.9 (3.0-6.3) | 1.1 (0.6-1.6) | 70% (57%-81%) |
| Lilongwe | 6% (6%-7%) | 25% (17%-32%) | 92% (84%-96%) | 2.8 (2.4-3.3) | 1.6 (1.0-2.9) | 68% (48%-78%) |
| Mchinji | 5% (5%-6%) | 30% (23%-39%) | 92% (77%-97%) | 3.5 (2.5-5.0) | 1.3 (0.7-2.0) | 73% (59%-85%) |
| Nkhotakota | 5% (4%-6%) | 38% (29%-44%) | 94% (73%-97%) | 3.1 (2.3-4.2) | 0.8 (0.5-1.4) | 78% (70%-86%) |
| Ntcheu | 8% (7%-9%) | 28% (21%-34%) | 93% (73%-98%) | 3.2 (2.6-4.1) | 1.9 (1.1-3.3) | 73% (57%-85%) |
| Ntchisi | 2% (2%-3%) | 39% (32%-44%) | 89% (70%-96%) | 3.0 (2.4-4.2) | 0.4 (0.2-0.9) | 76% (63%-81%) |
| Salima | 5% (4%-6%) | 29% (21%-35%) | 93% (83%-97%) | 4.2 (3.1-6.2) | 1.3 (0.8-1.9) | 74% (62%-81%) |
| **Southern** |  |  |  |  |  |  |
| Balaka | 8% (7%-9%) | 31% (25%-35%) | 96% (89%-98%) | 3.0 (2.7-3.6) | 1.9 (1.4-2.5) | 74% (68%-80%) |
| Blantyre | 14% (13%-15%) | 22% (16%-26%) | 87% (80%-93%) | 2.8 (2.6-3.2) | 4.2 (3.1-5.3) | 64% (54%-73%) |
| Chikwawa | 8% (7%-9%) | 26% (18%-34%) | 93% (81%-98%) | 3.7 (2.8-5.0) | 1.8 (1.1-2.9) | 75% (64%-83%) |
| Chiradzulu | 12% (11%-14%) | 28% (24%-33%) | 93% (85%-97%) | 2.3 (2.1-2.9) | 3.1 (2.1-4.0) | 70% (62%-78%) |
| Machinga | 7% (7%-9%) | 26% (18%-32%) | 90% (77%-96%) | 4.1 (3.5-5.1) | 2.0 (1.3-3.6) | 73% (58%-81%) |
| Mangochi | 9% (8%-10%) | 23% (16%-31%) | 93% (81%-97%) | 5.6 (4.6-7.5) | 2.7 (1.6-3.6) | 73% (67%-82%) |
| Mulanje | 16% (14%-17%) | 16% (8%-24%) | 95% (86%-97%) | 4.6 (4.0-5.3) | 5.2 (3.7-7.2) | 71% (62%-77%) |
| Mwanza | 7% (5%-8%) | 21% (11%-27%) | 87% (72%-96%) | 3.7 (2.6-5.3) | 2.2 (1.4-3.4) | 66% (53%-75%) |
| Neno | 8% (7%-10%) | 29% (21%-37%) | 95% (76%-98%) | 3.1 (2.7-3.9) | 2.0 (1.1-3.3) | 74% (59%-85%) |
| Nsanje | 10% (8%-12%) | 23% (13%-30%) | 53% (43%-71%) | 4.3 (3.1-7.2) | 4.8 (2.8-7.4) | 58% (43%-74%) |
| Phalombe | 15% (13%-17%) | 17% (9%-24%) | 96% (84%-98%) | 6.0 (4.5-8.0) | 4.6 (3.1-7.3) | 76% (63%-83%) |
| Thyolo | 12% (10%-13%) | 26% (21%-34%) | 93% (86%-97%) | 3.5 (3.0-4.7) | 2.8 (1.9-3.9) | 76% (69%-83%) |
| Zomba | 12% (11%-14%) | 22% (12%-28%) | 89% (81%-94%) | 3.4 (3.0-4.0) | 3.6 (2.4-5.5) | 71% (62%-79%) |

Incidence decreased by at least 50% in all districts between 2010 and 2021, although declines varied spatially. The smallest decrease was from 11.7 to 4.8 (a 58% (43% to 74%) decline) in Nsanje, while the largest was in Nkhotakota from 3.8 to 0.8 (a 78% (70% to 86%) decline).

Incidence declines corresponded to large increases in ART coverage in every district. Between 2010 and 2021, treatment at least doubled in every district among both men and women (Figure 2, Table 1). The smallest relative increase in ART coverage was an increase of 2.1 (1.6 to 2.7) times in Dowa, and the largest was a 6.0 (4.5 to 8.0) times increase in Phalombe. Phalombe had the second-lowest ART coverage in 2010, while Dowa had the second-highest, illustrating that the largest improvements were in the districts that had the lowest coverage in 2010. Lower ART coverage in 2010 was strongly associated with large increases between 2010 and 2021 (Figure 2).

### Subnational progress towards epidemic transition

In all 28 districts, incidence decreased by at least 20% in every posterior simulation (corresponding to posterior probabilities of 100%). The posterior probability of a 50%-or-greater decrease was above 90% in 26 of 28 districts, with Dowa and Nsanje only reaching 84% and 78%. No districts had 90% or higher posterior probabilities of incidence decreases of at least 75% (Figure 3); only five districts (Thyolo, Chikwawa, Nkhotakota, Ntchisi, and Phalombe) had 50% or higher posterior probabilities of 75% decreases or more.

**Figure 3:**
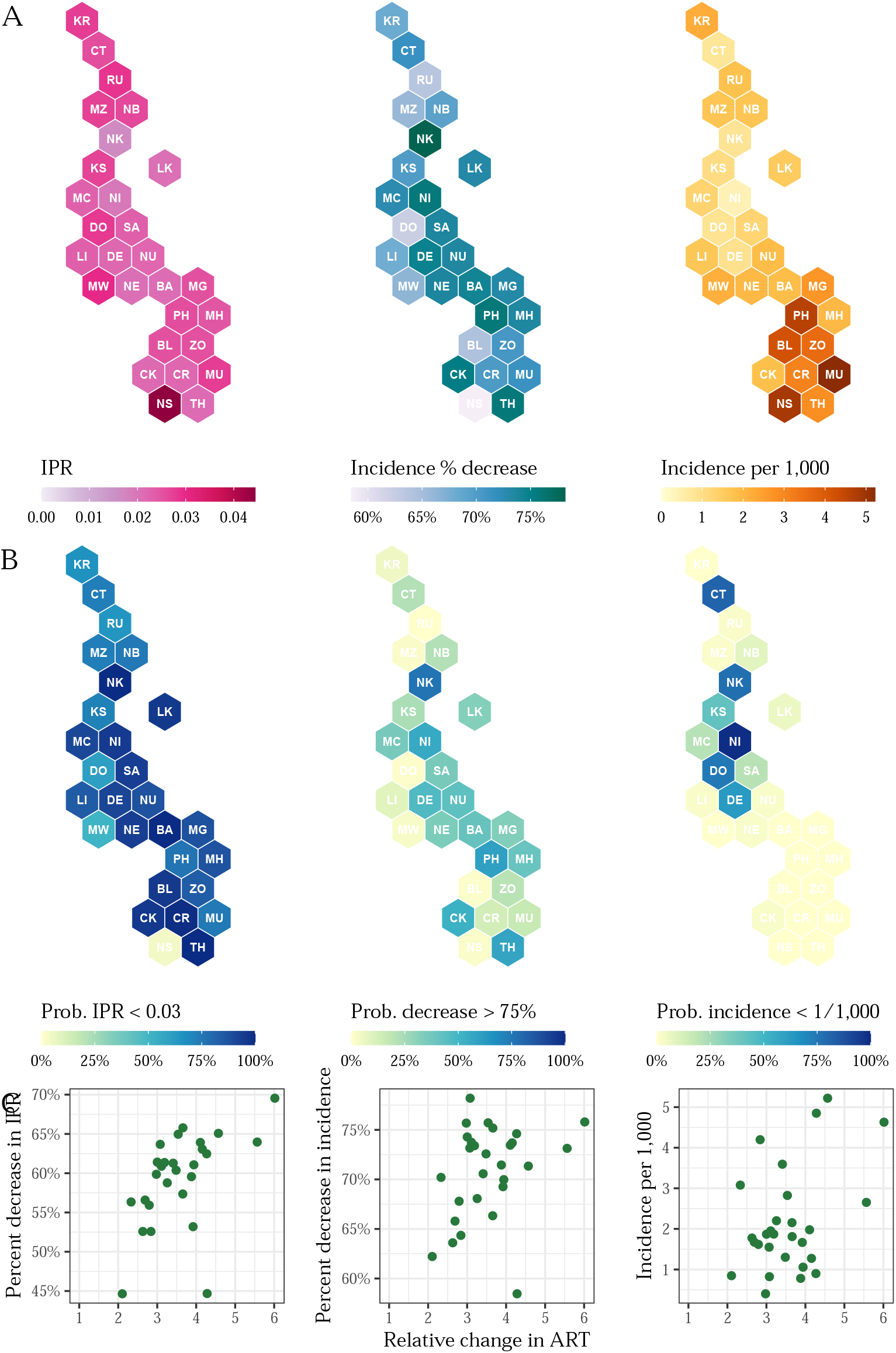
Changing subnational adult HIV incidence dynamics in Malawi. A) HIV IPR in 2021 (left), changes in HIV incidence between 2010 and 2021 (centre), HIV incidence in 2021 (right) among ages 15-49 by district in Malawi. B) Posterior probabilities of IPRs in 2021 less than 0.03 (left), changes in incidence exceeding 75% decreases between 2010 and 2021 (centre), and incidence less than 1 per 1,000 in 2021 (right). C) Scatter plots comparing IPR in 2021, percent change in incidence between 2010 and 2021, and incidence per 1,000 in 2021 to relative changes in ART coverage by district.

In 27 of 28 districts, at least half of the posterior density in incidence change was located between 60% and 80% incidence reductions between 2010 and 2020 (Figure 4), indicating that although no district definitively achieved the UN-targeted 75% reduction, many districts were approaching that threshold. Nationally, the posterior probability of a 75%-or-greater decrease was only 4.9%, but the probability of a 65%-or-greater decrease was 94.8%.

**Figure 4:**
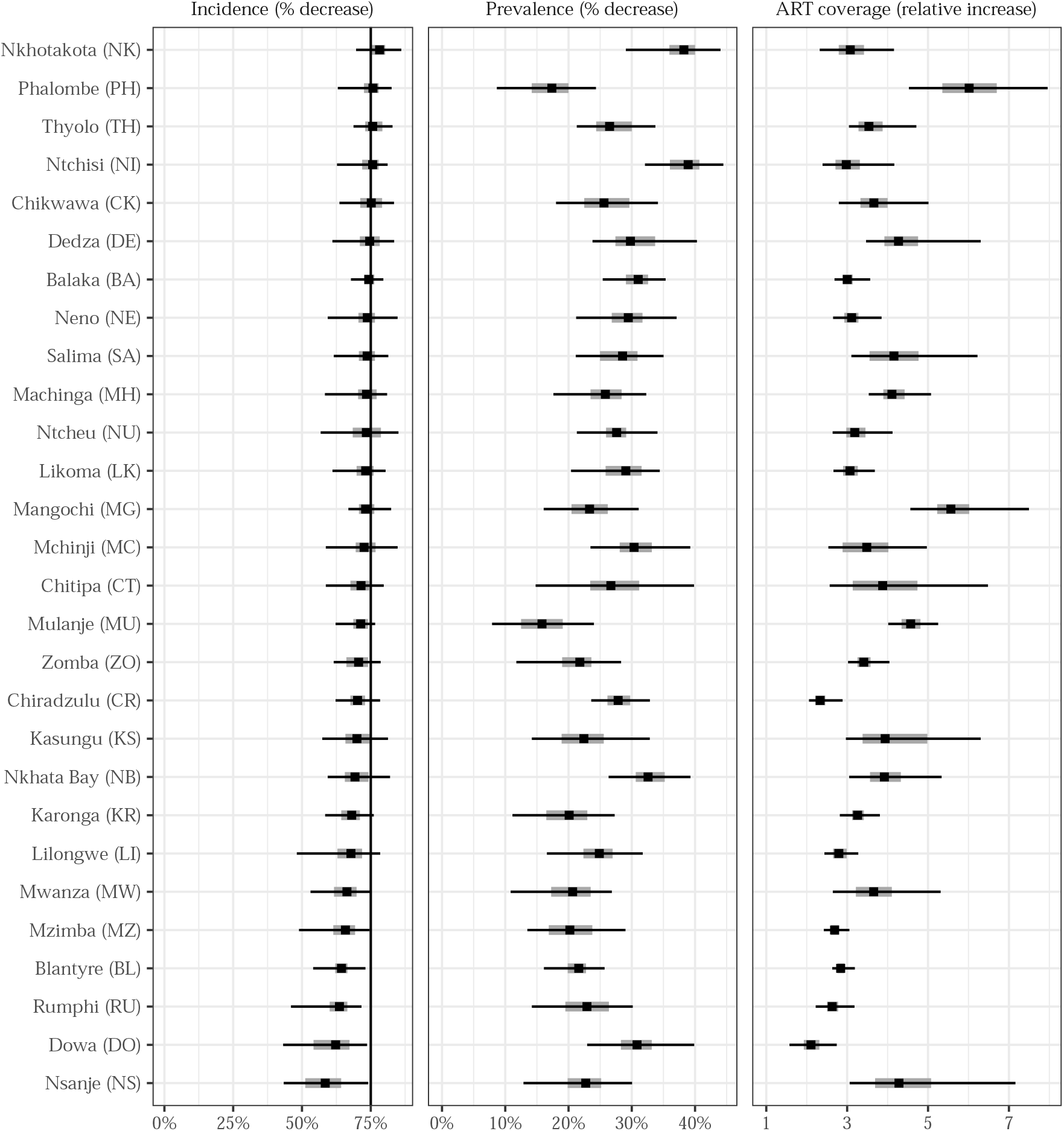
Changes in key HIV indicators among adults in Malawi, 2010-2021. Posterior median (points) changes in incidence risk, ART coverage, and prevalence with 95% and 50% credible intervals (lines and shaded regions, respectively). Districts are sorted vertically from highest median change in incidence to lowest.

The spatial structure of our model allowed us to infer subnational transmission dynamics. Following Ghys et al., we calculated the ratio of new infections to PLHIV (or *incidence-prevalence ratio*) and its inverse, which measures the number of PLHIV per new infection.^6^ The national incidence-prevalence ratio (IPR) was 0.026 (0.021 to 0.030), corresponding to one new infection per 38 (33 to 48) PLHIV. These figures represent considerable improvements from 2010, when the national-level IPR was 0.062 (0.058 to 0.064) or one new infection per 16 (16 to 17) PLHIV. The posterior probability that Malawi had met the 0.03 IPR threshold proposed by Ghys by 2021 was 97.4%.^6^

We estimated substantial spatial heterogeneity in HIV transmission. District-level IPRs in 2021 ranged from a high of 0.044 (0.028 to 0.057) in Nsanje to a low of 0.016 (0.011 to 0.024) in Nkhotakota. These IPRs correspond to one new infection per 22 (17 to 35) PLHIV in Nsanje and per 61 (41 to 91) PLHIV in Nkhotakota. In 2021, 12 of 28 districts had a 90% or greater posterior probability of an IPR less than 0.03. The district-level posterior probability of having reached an IPR of 0.03 in 2021 or lower was correlated with ART coverage in 2021 (Pearson *ρ*: 0.82) but not with the change in ART coverage between 2010 and 2021 (Pearson *ρ*: -0.16).

District-level changes in IPR varied less over space than absolute levels, due in part to uniformly large improvements in ART coverage. The percent decreases in IPR ranged from 45% (20% to 57%) to 70% (57% to 76%) in Dowa and Phalombe, respectively. Figure 3 compares changes in ART coverage and IPRs. Because the model accounts for the population-level impact of treatment on transmission, changes in transmission were closely correlated with changes in ART coverage. The negative correlation between ART coverage change and IPR change indicates that larger improvements in ART coverage were associated with larger declines in IPR. The outlier marked with a red point in this plot is Nsanje, in which large improvements in ART coverage did not result in the expected reductions in HIV transmission.

The right column of Figure 3 also compares estimated incidence to the threshold of one per 1,000 by 2030 proposed by Galvani et al. Nationally, Malawi has not met that threshold in 2021 (posterior probability 0.0%), but the posterior probabilities varied subnationally. Ntchisi was the only district to achieve a 90% or higher posterior probability of one new infection per 1,000 people in 2021. This posterior probability was less than 10% in 20 districts. The posterior probability of incidence less than one per 10,000 people in 2021 was 0.0% in every district.

### Increasing spatial heterogeneity in incidence

We measured spatial variability in the three indicators of interest by calculating the coefficient of variation (CV) across districts for each predicted year. Incidence varied more over space than prevalence and ART coverage (Figure 5). Despite increasing uniformity in ART coverage, the spatial heterogeneity in incidence and prevalence increased slightly between 2010 and 2021. This finding is consistent with our observation that the relative range in incidence across districts increased dramatically over the same period. The posterior probability that the coefficient of variation in incidence increased between 2010 and 2021 was 87%.

**Figure 5:**
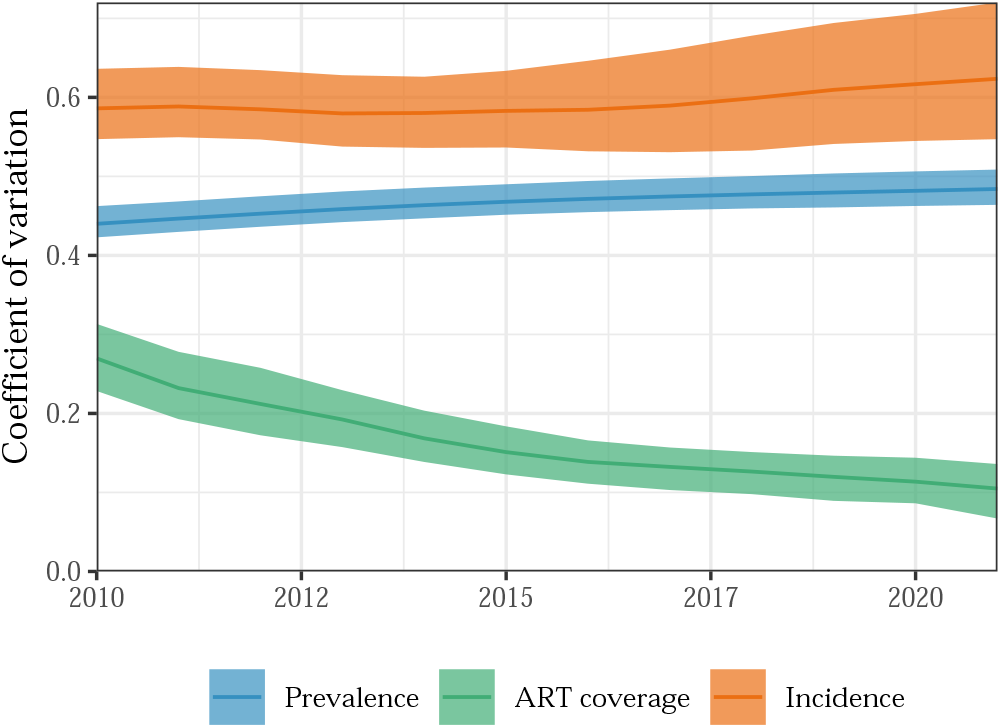
Changes in spatial heterogeneity in HIV indicators in Malawi, 2010-2021. Coefficients of variation (CVs) across districts in HIV incidence rate, prevalence, and ART coverage over time. Larger CVs indicates greater spatial variability. Shaded area represents 95% credible intervals across posterior epidemic draws.

## Discussion

Between 2010 and 2021, HIV incidence and prevalence decreased among both men and women in all 28 districts of Malawi, coinciding with large increases in ART coverage. There was substantial heterogeneity in both levels of and trends in prevalence and incidence, despite rapidly decreasing variability in ART coverage.

Our results highlight the continued success of Malawi’s HIV treatment programme. National ART coverage increased more than three-fold between 2010 and 2021. Districts with the largest increases between 2010 and 2021 were those with the lowest coverage in 2010. Whereas a testing and treatment strategy that prioritised the highest burden areas could have exacerbated existing gaps, the public health approach deployed in Malawi has yielded highly equitable treatment coverage.

Large and widespread improvements in ART coverage resulted in commensurate decreases in incidence across the country. Although we estimated that the incidence rate varied 14-fold across districts in 2021, incidence declined by at least 50% in every district. In Twelve districts, the posterior probability of incidence-prevalence ratio below the 0.03 threshold in 2021 was above 90%.^6^ Changes in district-level IPRs were strongly correlated with improvements in ART coverage, but the posterior probability of having met the 0.03 threshold was most highly correlated with the current ART coverage.

Consistent with simulations presented by Galvani et al., the elimination target based on an incidence-level below one new infection per 1,000 was more difficult to attain than the IPR-based target, with only one district having reached less than one new infection per 1,000 people in 2021.^9^ Both the IPR and absolute incidence level decreased dramatically over the study period, reflecting the progress Malawi has made in reducing HIV burden. We also note that because our model produces internally consistent estimates of prevalence and incidence, it could be used to estimate any number of epidemic control metrics and posterior probabilities of associated targets.

District-level data from the 2020-2021 MPHIA survey were not publicly available at the time of this analysis, so we can use the national level survey estimates as out-of-sample validation for our estimates. The survey found a prevalence of 8.0% (7.5% to 8.5%) among adults aged 15-49, while we estimated national prevalence of 7.9% (7.6% to 8.2%) in 2021.^39^ Our estimated national incidence of 2.3 (1.7 to 2.7) new infections per 1,000 people was also close to the survey estimate of 2.3 (1.1 to 3.6). Currently, these comparisons provide an additional layer of validation for our model, but when the district-level survey data become available, our model will be able to fit directly to them.

Although improvements in incidence and treatment coverage were substantial in every district, the spatial variation in incidence was increasing as of 2021. These results are consistent with theories that the epidemic will recede into harder-to-reach populations and areas as ART coverage reaches high levels. Broad, large-scale treatment provision programmes have succeeded in improving outcomes in the general population, but they can only partially fill treatment gaps among groups that have greater difficulty or hesitancy engaging with centralised healthcare systems.^44,45^ As treatment coverage continues to improve, population-level incidence will increasingly be determined by other, more heterogeneous factors (e.g. prevalence of commercial sex work, education, etc.) and the marginal effects of improved coverage will decrease.^14^ Failing to patch seemingly small treatment gaps could therefore lead to the emergence of *source-sink* dynamics that could stall progress towards control and elimination.^16^ The evidence presented in Figure 5 is consistent with this theory, but the estimated increases in spatial heterogeneity were small relative to the large decreases changes in incidence over time. Analyses of data collected over the next several years will offer a clearer picture of how the impact of population-level ART coverage on incidence in high-prevalence settings changes as treatment coverage nears 100%.

Regarding implications for HIV programming in Malawi, our results confirm the large and equitable impact of improved access to HIV services on high ART coverage and low and declining incidence. Further ART scale-up resources should focus on the districts with relatively higher incidence and lagging ART coverage, while ensuring appropriate levels of testing, linkage, and retention programmes are sustained in all districts to maintain the high ART coverage. The efficiency and cost-effectiveness of other primary prevention interventions depends critically on HIV incidence in the target population. For example, WHO recommend that HIV pre-exposure prophylaxis (PrEP) should be prioritised for locations and population groups with HIV incidence above 30 per 1,000 to be cost-effective.^46^ We estimated that general population incidence was six-fold lower than this in 2021, even in the highest incidence districts. This underscores that PrEP and other effective but expensive primary prevention interventions are unlikely to be an efficient use of health resources to scale-up to the general population in any districts. Instead, access should be prioritised among populations with specific risk factors in high incidence districts, as outlined in the Global AIDS Strategy 2021-2026.^47–49^

This analysis has a number of limitations. First, we did not explicitly model transmission between districts in the model HIV incidence; the district incidence rate was related to prevalence and ART coverage in that district. Model comparisons indicated that omitting spatial transmission yielded slightly better out-of-sample fit than alternative specifications that included spatial mixing (Supplemental Material Section 1.3.1.1). This assumption is consistent with a recent analysis of viral genetic data suggesting that HIV transmission in SSA is highly local.^50^ Second, we omitted age structure from the compartmental model. Age is a critical determinant of HIV infection risk and mortality, but explicitly representing age resulted in computationally intractable number of compartments for our inference framework. Instead we accounted for the effects of age by age-standardising mortality and progression rates (Supplemental Material Section 1.3). Future work is needed to identify computational strategies for efficiently solving joint epidemic-demographic models. Third, we relied on fixed assumptions about HIV disease progression with and without treatment, non-AIDS mortality, and the effect of population-level ART coverage on transmission. These assumptions align with those made by other compartmental models of HIV, but their applicability to subnational regions of Malawi can still be questioned. Of particular importance is the assumption made about the effect of ART coverage on transmission. The observed association between changes in ART coverage is partly determined by this fixed assumption. Fourth, for computational tractability and our focus on estimating HIV incidence trends since 2010, our model started in year 1995 instead of from the start of the epidemic. Between 1995 and 2005 national incidence estimates from our model differed from national HIV estimates published by UNAIDS, but after 2005 national incidence rate estimates from our model were very similar to national UNAIDS estimates (Supplemental Figure 36). Future implementations of this model will include an option to calibrate to external estimates of national-level prevalence and incidence. Finally, for tractability, the model of ART attendance assumed that individuals decide where to seek treatment independently every quarter. In future work, we aim to develop a more realistic model to estimate treatment initiation, retention, and transferring.

Despite these limitations, the estimates presented here shed new light on how HIV incidence has evolved as ART coverage expands and demonstrates a new modelling approach for Malawi, and other countries, to synthesise surveillance data for a more a more spatially granular understanding of HIV dynamics. We found that the rapid and equitable scale-up of treatment in Malawi resulted in large improvements in ART coverage and incidence across the country, with some districts meeting “epidemic control” the threshold proposed by Ghys.^6^ We observed a small increase in the spatial heterogeneity of incidence, consistent with theories that the epidemic is becoming increasingly concentrated in the high-ART era. If the impact of broad, general-population treatment provision on incidence does decrease over the next several years, then the success of HIV policy-making will depend critically on how well it targets the right people in the right places.^13^ Future models used to monitor HIV epidemics must meet these needs.

## Methods

We fit a spatio-temporal Bayesian epidemic model of HIV to district-specific HIV data collected in Malawi between 1995 and 2021. The model infers three components for each district by sex: the HIV transmission rate by untreated adults over time, the probability of ART initiation among untreated adults, and the initial HIV prevalence in 1995. We estimated quarterly HIV prevalence, incidence, and treatment coverage for adults aged 15-49 from 2010 to 2021 for the 28 districts of Malawi. The sections below provide an overview of the data sources, model structure, statistical inference, and analyses. Supplemental Material presents the technical details of the model and results of model comparisons to select the final model specification.

### Data

We incorporated data from three sources into our model: nationally representative household surveys, HIV prevalence among pregnant women accessing HIV testing at public ANC facilities, and reports of the number of patients receiving ART.

### Household survey data

Four nationally representative household surveys with HIV serological testing have been conducted HIV testing in Malawi: the 2004, 2010, and 2015-16 Malawi Demographic and Household Surveys (MDHS), and the 2015-2016 Malawi Population-based HIV Impact Assessment (MPHIA) survey.^40–43^ A second MPHIA survey was conducted in 2020-21, but district-level survey data were not yet available.^39^ From the three DHSs, we extracted district- and sex-specific HIV prevalence, and from MPHIA we extracted district- and sex-specific HIV prevalence, ART coverage, and the proportion recently infected according to a recent infection testing algorithm. HIV positive respondents were classified as using ART if either antiretroviral biomarker was detected or the respondent self-reported using ART, consistent with primary survey reports of ART coverage. For both survey series, we restricted to participants aged 15 to 49 years.

### ANC facility data

We combined data on HIV prevalence among pregnant women attending public ANC from two sources: ANC surveillance conducted at selected sentinel sites between 1994 and 2010 and routinely reported results of HIV testing among all pregnant women attending ANC from 2011 onwards.^51^ ANC sentinel surveillance was conducted in approximately two facilities in each district every 2 to 3 years. Facility-level HIV prevalence observations were extracted from data inputs to the Estimation and Projection Package (EPP) model within the UNAIDS Spectrum estimates software.^52^ Routine ANC testing prevalence for 2011 onward for the same facilities was extracted from the Malawi Department of HIV & AIDS Management Information System (DHAMIS), and aggregate to quarterly temporal resolution. For the 730 facilities not included in ANC sentinel surveillance, we aggregated routine ANC testing data to quarterly, district-level aggregate prevalence observations.

### ART programme data

We aggregated the number of patients receiving ART at health facilities in each district at the end of each quarter from the DHAMIS. Médecins Sans Frontières began operating treatment clinics in Chiradzulu before the national ART scale-up, so we supplemented the DHAMIS data with reported patient counts in Chiradzulu from 2002 to 2004 from a published report.^53^ Data included ART patients of all ages, so we multiplied each count by the share of ART patients that were between 15 and 49 years old in each year from national Spectrum model estimates.^54^

### District-level population

We used population estimates of the district population aged 15-49 years by sex from the National Statistical Office of Malawi, linearly interpolated to obtain quarterly estimates.^55^ We used Beers graduation to disaggregate five-year age categories into single-year ages to obtain estimates of the number of individuals ageing in and out of the 15-49 year-old population each year.^56^

### Bayesian epidemic model

We created a compartmental epidemic model of HIV to simulate HIV incidence, prevalence, and treatment coverage.^57^ The HIV transmission rate, ART initiation rate, and initial HIV prevalence in 1995 are specified by generalised additive models. Given a set of parameters, a single evaluation of this model is executed as follows:

1. Linear models predict region-/sex-/time-specific series of HIV transmission rates, ART initiation rates, and initial prevalence.
2. The epidemic model is initialised at the state determined by the estimated initial prevalence from (1) and simulated using predicted transmission rates, ART initiation rates, and a fixed set of natural history parameters.
3. The likelihood of each data sources is evaluated as a function of predicted HIV prevalence, incidence, and ART coverage from the epidemic model and additional observation model parameters reflecting relevant biases and overdispersion in each data source.

### Compartmental model of HIV

The deterministic compartmental HIV epidemic model tracks the sizes of susceptible, infected without treatment and infected with treatment populations by sex and district. The system of ordinary differential equations that define the model is in Supplemental Material Section 1.3.

Untreated and treated infection compartments are stratified into four disease progression stages defined by CD4 T cell count bins (500 or more, 350-500, 200-350, and less than 200). Susceptible individuals can die or become infected with HIV.Untreated PLHIV can die, begin treatment, or progress to the next CD4 category. Treated PLHIV can die or interrupt treatment.

The initial state of the epidemic model, the transmission rate of HIV, and the rate of treatment initiation are inferred. Other model dynamics are fixed at exogenously defined values. Time- and sex-specific mortality and rates were calculated using time-, sex-, and age-specific death counts from the UNAIDS Spectrum model, allowing us to account for how the changing age age distribution of PLHIV affects average mortality rates.^54^ Progression rates through CD4 categories were calculated using the formulation from and the average age of PLHIV not on treatment from Spectrum. The time- and sex-varying distribution of entrants and exits into each compartment is fixed at values age-aggregated values from EPP-ASM. Supplemental Material Section 1.3 details the calculation and implementation of each assumption.

We calculate time-, sex-, and region-specific incidence as a function of time-, sex-, and region-specific transmission rates and opposite-sex prevalence that has been adjusted for ART coverage. Following EPP, we assume that HIV transmission would be reduced by 80% at 100% ART coverage.^52^

### Generalised additive models for model components

The HIV transmission rate by untreated adults in each quarter is modelled using region-specific intercepts and region-specific autoregressive integrated moving average (ARIMA) terms with respect to time, which allow for flexible changes over time within district.^58^ This model was conceived as a generalisation of the “r-spline” model used in EPP.^59^ The sex ratio of transmission is modelled using a log-linear model with respect to time that is shared across all regions.

In contrast to previous inferential models of HIV incidence, our model infers ART initiation rates and fits to patient counts. The model of ART initiation is similar to the model of HIV transmission rates, predicting region-, sex-, and time-specific initiation with district intercepts, district ARIMA terms, and an inferred sex intercept.

The initial state of the compartmental model is modelled by independent and identically distributed district-specific random effects for logit-transformed HIV prevalence in 1995. The initial prevalence is allocated to each CD4 compartment using pre-calculated distributions from the Spectrum model.^60^ The national-level initial prevalence was constrained to be similar to estimated prevalence in Malawi in 1995 by placing an informative prior on the aggregate of inferred district prevalences.

### Observation models

Solving the epidemic model with the dynamics predicted by the three models described in Section 4.2.2 produces internally consistent estimates of HIV prevalence, incidence, and treatment coverage by region, sex, and calendar quarter. These are related to the data described in the “Data” Section with a series of observation models.

### Household survey data

We assume that household survey data were representative by district and sex over their collection periods. These surveys are collected via complex multi-stage sampling schemes and therefore the estimates we derive from them must be accompanied by design-based variances. For district/sex-specific HIV prevalence and ART coverage observations, we calculate the effective sample size and number of cases based on design-based survey estimates and standard errors. We use a binomial model for the likelihood conditional on predicted rates.^61^ This method has been used in previous HIV mapping exercises.^12,30^

For recency assays, we observed the effective number of people with recent infection. Kassanjee et al. derived an estimator for incidence given a proportion of positive recency assays, which Eaton et al. manipulated to give the expected proportion recently infected as a function of incidence.^30,62^ Let *π*_*i*_ be the true proportion of people who were infected recently, *λ*_*i*_ be the true incidence rate, and *ρ*_*i*_ be true prevalence. Then, following Eaton et al.,

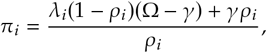

where Ω is the mean duration of recent infection and *γ* is the false positive rate of the recency assay. We assume that Ω = 130/365 and *γ* = 0, consistent with primary analysis of MPHIA 2015-16 survey data.^43^ We treat *π*_*i*_ as the probability of a positive recency assay.

### ANC facility data

Facility-level HIV prevalence at ANC differ from district population prevalence because both selected facilities may not be representative of the district population and because HIV prevalence among pregnant women is systematically different from general population prevalence. Previous HIV models have addressed this by incorporating facility-specific random effects.^63^ For additional district-aggregated facility data not previously included in EPP, we include a random effect capturing deviation between ANC prevalence and population prevalence. We extend the random intercepts model proposed by Alkema, Raftery, and Clark to allow the representativeness of each facility to change linearly over time, reflecting that, as incidence declines and the population of PLHIV ages, HIV prevalence among pregnant women declines more rapidly than general population prevalence.^64^ The details of this model are provided in the Supplemental Material.

We assume that the resulting facility-level predicted prevalence is the true population mean from which the quarterly ANC HIV testing data were sampled. We used a beta-binomial likelihood to capture overdispersion in observed ANC prevalence observations.^65^

### ART programme data

Finally, we fit to data on the number of patients receiving treatment in each district at the end of each quarter. Because household surveys are residency-based and patients may seek treatment in a different district than they live, there is a fundamental disconnect between survey ART coverage estimates and ART programme data. Extending Eaton et al., we implement a model of ART attendance that allocates residents on ART to treatment regions, detailed in the Supplemental Material.^30^ We use a modified negative binomial likelihood for the observed ART patient counts, with a mean equal to total number of allocated patients and both linear and quadratic scaling terms in the variance.^66^

### Model selection

We fit a grid of 146 different transmission rate model specifications data sets that held out data beginning in each year from 2015 through 2020. Between 146 specifications and six hold-out horizons, we fit 876 models. We measured out-of-sample performance by calculating root mean squared errors (RMSEs) on held-out ANC prevalence data and ART programme data.

### Analysis of descriptive results

We predicted quarterly HIV prevalence, incidence, and treatment coverage for adults aged 15-49 years by district and sex for all 28 districts of Malawi from 2010 to 2021. We calculated HIV prevalence as the number of PLHIV divided by the total population, ART coverage as the number of PLHIV on treatment divided by the total number of PLHIV regardless of treatment eligibility, and reported time-, district-, and sex-specific incidence rates directly from the epidemic model. We additionally calculated the percent change in all three metrics from the first quarter of 2010 through the final quarter of 2021. Whenever appropriate, we present median estimates with 95% credible intervals in parentheses.

We calculated the posterior probability of incidence having changed by more than predefined thresholds by finding the share of posteriors samples with percent change values greater or less than predefined levels. To quantify changing spatial heterogeneity, we calculated the coefficient of variation of incidence rate, prevalence, and ART coverage across districts in each year.

Finally, to assess determinants of changes in incidence, we linearly regressed estimated changes in sex-specific district-level incidence between 2010 and 2021 on sex, region, the proportion of adults aged between 15 and 25, incidence in 2010, and the change in ART coverage between 2010 and 2021.

### Implementation

Our model is implemented in C++ using the **Template Model Builder** (TMB) R library.^67^ We used the **tmbstan** library to perform inference with the No-U-Turn Sampler (NUTS), as implemented in Stan.^68,69^ All plots were produced with the **ggplot2** library, and the hexagonal tile maps were produced using the **geogrid** library.^70,71^

## Data Availability

Facility-level aggregate data from the Malawi DHAMIS system are publicly available from the Malawi Ministry of Health (https://dms.hiv.health.gov.mw/dataset/). Data from the DHS Program are available at the DHS website (https://dhsprogram.com/Data/) upon registration. Data from the PHIA surveys are available upon registration from the PHIA website (https://phia-data.icap.columbia.edu/). District population projections are publicly available from the Malawi National Statistics Office (http://www.nsomalawi.mw/). The C++ code for the analysis is available on Github: [https://github.com/twolock/mwi-incidence-code/](https://github.com/twolock/mwi-incidence-code/). The analysis is extremely computationally intensive and built specifically for use on the Imperial College London High Performance Computing cluster, so we cannot provide a reproducible version of this paper.

## Data availability

Facility-level aggregate data from the Malawi DHAMIS system are publicly available from the Malawi Ministry of Health (https://dms.hiv.health.gov.mw/dataset/. Data from the DHS Program are available at the DHS website (https://dhsprogram.com/Data/) upon registration. Data from the PHIA surveys are available upon registration from the PHIA website (https://phia-data.icap.columbia.edu/). District population projections are publicly available from the Malawi National Statistics Office (http://www.nsomalawi.mw/).

## Code availability

The C++ code for the analysis is available on Github: https://github.com/twolock/mwi-incidence-code/. The analysis is extremely computationally intensive and built specifically for use on the Imperial College London High Performance Computing cluster, so we cannot provide a reproducible version of this paper.

## Acknowledgements

We thank the investigators of and participants in the 2004, 2010, and 2015-16 Malawi Demographic and Health Surveys and the 2015-2016 Malawi Population HIV Impact Assessment survey. TMW thanks the members of the Imperial College HIV Inference Research Group for their helpful advice. This work was conducted as part of TMW’s PhD thesis, which was funded by the Imperial College President’s PhD Scholarship. JWE and TMW were supported by the Bill and Melinda Gates Foundation (INV-002606) and the MRC Centre for Global Infectious Disease Analysis (reference MR/R015600/1), jointly funded by the UK Medical Research Council (MRC) and the UK Foreign, Commonwealth & Development Office (FCDO), under the MRC/FCDO Concordat agreement and is also part of the EDCTP2 programme supported by the European Union. JWE was supported by the Bill and Melinda Gates Foundation (INV-006733) and the National Institute of Allergy and Infectious Disease of the National Institutes of Health under award number R01AI136664.

## Author contributions

TMW, SF and JWE conceived of the study. RN, AJ, SM, and TC oversaw implementation of the Malawi HIV programme and management and interpretation Malawi HIV programme data. TMW developed the statistical model, conducted the analysis, and drafted this article. All authors revised the article for intellectual content and approved the final manuscript for submission.

