## Supplemental Material for "Subnational HIV incidence trends in Malawi: large, heterogeneous declines across space"

13 January 2023

**2 Model selection results** **15**

**3 Fit in all districts** **18**

**4 Comparison to UNAIDS 2021 estimates (UNAIDS 2021)** **46**

**References** **47**

### 1 Detailed methodology

This section describes the general model framework and definitions of data sources, without reference to country-specific implementation. Details about specific data sources and inputs for Malawi are described in the main text.

#### 1.1 Data sources

The collated set of data, $\mathcal{D}$, considered by our model consists of a subset of household surveys, ANC facility HIV test results, and ART programme patient counts. Table 1 outlines the data sources and the indicators they measure. Table 2 outlines the exact data sources used in this analysis.

Supplemental Table 1: Taxonomy of population-level HIV data sources included in our model

| Indicator | Data Source | Numerator | Denominator |
|---|---|---|---|
| Prevalence | Household surveys | # of positive HIV tests | # of HIV tests |
| ANC prevalence | Sentinel ANC clinics and routine ANC testing | # of positive HIV tests | # of HIV tests |
| ART coverage | Household surveys | # of positive ART tests | # of ART tests |
| ART patients | Routine health service delivery data | # of ART patients | - |
| Recency status | Household surveys | # of positive recency assays | # of recency assays |

Supplemental Table 2: Population-level data sources from Malawi used in this analysis.

| Name | Type | Years | Prevalence | Treatment | Recency |
|---|---|---|---|---|---|
| 2004 DHS | HH Survey | 2004 | ✓ | | |
| 2010 DHS | HH Survey | 2010 | ✓ | | |
| 2015-2016 DHS | HH Survey | 2015-2016 | ✓ | | |
| MPHIA 2015-2016 | HH Survey | 2015-2016 | ✓ | ✓ | ✓ |
| UNAIDS ANC Data | Sentinel surveillance | 1995-2010 | ✓ | | |
| DHAMIS | Facility reports | 2011-present | ✓ | | |
| DHAMIS | Facility reports | 2005-present | | ✓ | |

#### 1.2 Model overview

The model represents the dynamics necessary to simulate an epidemic model as non-linear functions of time, space, and sex (Hastie and Tibshirani 1986), aggregates the epidemic model's projections to produce estimates, and uses the observation model to relate those estimates to data. For a single draw from the posterior density:

1. A set of process parameters, $\theta_P$, are used to model region-/sex-/time-specific series of HIV transmission rates, ART initiation rates, and initial prevalence.
2. The epidemic model is initialised at the state determined by the estimated initial prevalence from (1) and integrated using the estimated transmission rates, ART initiation rates, and a set of exogenous, fixed parameters.
3. Predictions from the epidemic model are aggregated to produce estimates of HIV prevalence, ART coverage, and ART patients at the same spatio-temporal resolution as each dataset.
4. Predicted HIV prevalence, incidence, and ART coverage are used with an additional set of parameters, $\theta_O$, to evaluate the observation model given a collated dataset $\mathcal{D}$.

Figure 1 presents a simplified representation of the model. The first step from the above list is represented by every node to the left of $M$, the second step is $M$, and the third step is to the right of $M$.

#### 1.3 Compartmental model of HIV

We use a deterministic compartmental model of HIV to simulate HIV prevalence and incidence, ART coverage, and the number of PLHIV receiving treatment ($\rho_{r,g}(t)$,$\lambda_{r,g}(t)$, $\alpha_{r,g}(t)$, and $A_{r,g}(t)$, respectively).

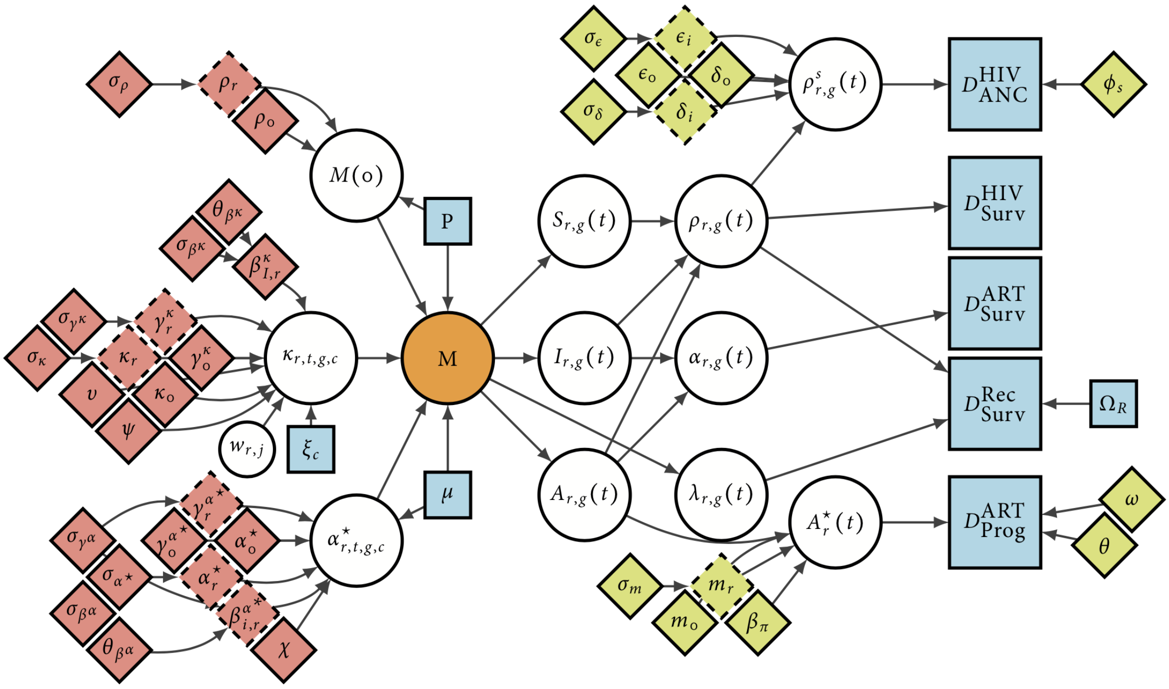

Supplemental Figure 1: A simplified graphical representation of our model of HIV incidence. Diamonds are parameters, white circles are (deterministic) calculations, and blue squares are external data. Red parameters influence the process model, and green/yellow parameters influence the observation model.

We model the number of people of sex $g$ in region $r$ at time $t$ by disease status with the following set of ordinary differential equations:

$$
\begin{aligned}
\frac{\partial S_{r,g}(t)}{\partial t} =& S_{r,g}(t) \cdot (-\lambda_{r,g}(t) - \mu^S_{t,g}) + E^S_{r,t,g} \\
\frac{\partial I_{r,g,c}(t)}{\partial t} =& I_{r,g,c}(t) \cdot (-(\mu^S_{t,g} + \mu^I_{t,g,c}) - \alpha^*_{r,t,g,c} - \iota_{g,c,t}) + \lambda_{r,g,c}(t) S_{r,g}(t) + \\
& \eta A_{r,g,c}(t) + \iota_{g,c-1} I_{r,g,c-1}(t) + E^I_{r,t,g,c} \\
\frac{\partial A_{r,g,c}(t)}{\partial t} =& A_{r,g,c}(t) \cdot (-\mu^S_{t,g} - \mu^A_{t,g,c} - \eta) + \alpha^*_{r,t,g,c} I(t) + E^A_{r,t,g,c}.
\end{aligned}
\tag{1}
$$

This model is solved using the forward Euler method with a step size of 0.25 years. We denote the number of susceptibles $S_{r,g}(t)$, the number infected in disease stage $c$ without treatment $I_{r,g,c}(t)$, and the number infected with treatment who began treatment at disease stage $c$ $A_{r,g,c}(t)$. We define $c$ to be one of four CD4 compartments, consistent with those defined by the Thembisa model (Johnson and Dorrington 2019). Figure 2 outlines the structure of disease progression in the model.

A susceptible individual can either die at rate $\mu^S_{t,g}$ or become infected through contact with the opposite sex at rate $\lambda_{r,g}(t)$. An infected individual without treatment can die at rate $\mu^S_{t,g} + \mu^I_{t,g,c}$, begin treatment with probability $\alpha^*_{r,t,g,c}$, or progress to the next disease stage at rate $\iota_{g,c,t}$. Finally, an individual on treatment can die at rate $\mu^S_{t,g} + \mu^A_{t,g,c}$ or interrupt treatment with annual probability $\eta$. Treatment interruption is difficult to measure, so we have fixed $\eta$ to be 6% annually, adjusting a published figure to account for improvements in the treatment programme (Yu et al. 2007).

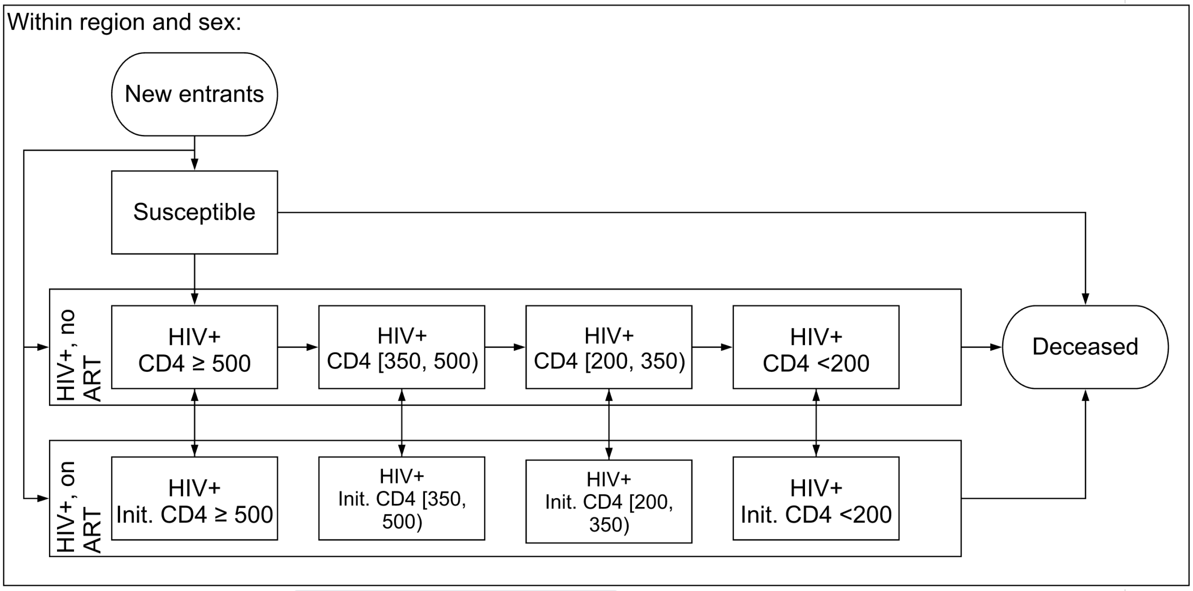

Supplemental Figure 2: Diagram of compartmental model of HIV used in this analysis

We stratify PLHIV with and without treatment into four CD4 categories to accurately model the survival distribution from HIV infection to AIDS-related death and to be able to capture changing eligibility for ART in which treatment was restricted to those with the lowest CD4 counts.

We calculate the CD4 stage progression rates, $\iota_{g,c,t}$, using assumptions described by Johnson and Dorrington (2019). Taking the average time spent in each CD4 category from Table 3.1, we apply Equation 3.1 to the Spectrum-estimated year-/sex-specific average age of PLHIV between 15-49:

$$\iota_{g,c,t} = \iota_c 0.96^g (1 + k)^{(x-30)/10}, \tag{2}$$

where $\iota_c$ is the average annual rate of progression from category $c$ to $c + 1$, 0.96 is the progression rate of women relative to men, and $k = 0.18$ is the proportionate increase in progression associated with a ten-year increase age. Following Table 3.1, we fix $\iota = (3.16, 2.13, 3.20)$ to be the expected number of years spent in CD4 category without treatment for the three highest CD4 categories. The final category is terminal, so its expected duration is not defined. The age distribution of PLHIV from Spectrum is itself an estimate and therefore might be inaccurate or contain uncertainty, which is not considered here; the approach described above is a simple solution to account for limitation that the model does not explicitly represent age structure.

To account for the shifting age distribution of PLHIV and gradual improvement of HIV patient outcomes without treatment in SSA, we calculate age-adjusted mortality rates based on the assumptions and results from EPP-ASM (Eaton et al. 2019). We use input mortality rates and predicted counts of PLHIV by age in each CD4 bin to find year-/age-/sex-/CD4-specific expected death counts. We aggregate the death and population counts to align them with the year-/sex-/CD4 groups used here and recalculate the mortality rates.

Each $E$ term in Equation (1) is the net number of people ageing in or out of the 15-49 year old population. HIV prevalence varies systematically with age in sub-Saharan Africa, so we must consider the possibility that the distributions of people ageing in and out across compartment vary from those of the general population. For example, if the population of PLHIV is ageing, we would expect prevalence among people ageing out to increase over time.

We use population data and national age-/sex-specific estimates of the share of people in each disease compartment from Spectrum to obtain regional estimates of the number of people turning 15 and 50 years old by sex, $E^{15}_{r,t,g}$ and $E^{50}_{r,t,g}$, respectively. The national-level estimates from Spectrum inherently weigh each region proportionately to its population, so if the regional distribution of individuals across compartment is correlated with population, the national-level disease status distributions will represent smaller regions poorly. We therefore apply region-specific adjustments to the estimated Spectrum distributions at each time point. Specifically, we adjust the odds from Spectrum of being in stage $c$ of compartment $C$ relative to being susceptible with the model's current regional prediction. Taking people ageing in as an example, the adjusted odds of an individual being in CD4 bin $c$ relative to not being infected with HIV are:

$$\Delta^{15,C}_{t,g,c} = \frac{C_{r,g,c}(t)}{S_{r,g}(t)} \frac{C^{15,\text{Spec}}_{g,c,t}}{S^{15,\text{Spec}}_{g,t}}, \tag{3}$$

noting that the denominators have cancelled. Then, fixing $o^{15,S}_{t,g} = 1.0$, we solve for the proportion of people in each compartment:

$$\nu^{15,C}_{t,g,c} = \frac{\Delta^{15,C}_{t,g,c}}{1 + \sum_{J \in I,A} \sum_{i=1}^{4} \Delta^{15,J}_{t,g,i}} \tag{4}$$

Finally, we calculate the net numbers of people ageing in and out for a given compartment, $C$ as

$$E^{C}_{r,t,g,c} = \nu^{15,C}_{t,g,c} E^{15}_{r,t,g} - \nu^{50,C}_{t,g,c} E^{50}_{r,t,g}. \tag{5}$$

This method accounts for the possibility that the distributions of people ageing into or out of the population across compartment vary from that of the general population.

We calculate prevalence as

$$\rho_{r,g}(t) = \frac{\sum_{c=1}^{4} [I_{r,g,c}(t) + A_{r,g,c}(t)]}{S_{r,g}(t) + \sum_{c=1}^{4} [I_{r,g,c}(t) + A_{r,g,c}(t)]} \tag{6}$$

and ART coverage as

$$\alpha_{r,g}(t) = \frac{\sum_{c=1}^{4} A_{r,g,c}(t)}{\sum_{c=1}^{4}[I_{r,g,c}(t) + A_{r,g,c}(t)]}. \tag{7}$$

With the compartmental model defined, we will define the models for incidence, ART initiation, and the initial state of the model, denoted $\lambda_{r,g}(t)$, $\alpha^*_{r,t,g,c}$, and $(S_{r,g}(0), I_{r,g}(0), A_{r,g}(0))$, respectively, for all $r \in \{1, ..., R\}$ and $g \in \{0, 1\}$.

##### 1.3.1 Generalised additive models for HIV transmission and ART initiation rates

We assume that a subset of the dynamics governing the compartmental model (mortality, disease progression, etc.) are fixed and drawn from other data sources and models, but three key components (incidence, ART initiation rate, and the initial state) are inferred from data. Each of these quantities is represented by an underlying generalised additive model (Hastie and Tibshirani 1986).

**1.3.1.1 Model of HIV incidence** We model incidence, $\lambda_{r,g}(t)$, as a log-linear function of time-varying transmission rates, opposite-sex prevalence, and ART coverage. Specifically, we have:

$$\begin{aligned} \log \lambda_{r,g,c}(t) \quad &= \quad \log \xi_c + g \cdot (\psi + \upsilon t) + \log \kappa_{r,t} + \\ &\quad\quad (I_{r,g^*,c}(t) + (1-\omega)A_{r,g^*,c}(t))/N_{g,r}. \end{aligned} \tag{8}$$

We use the relative infectiousness ratios listed in Table 3.1 in Johnson and Dorrington (2019) to fix the values of $\xi_c$ and set the following priors on the sex ratio of transmission parameters:

$$\psi, \upsilon \sim \mathrm{N}(0, 5). \tag{9}$$

In Wolock (2022), an alternative specification for this model that allowed for transmission across districts was considered. There was little empirical difference between models that included cross-district spatial transmission dynamics and those that did not. The model specification study performed in Chapter 4 of that thesis indicated slightly that the model without spatial transmission offered the best out-of-sample fit out of a set of candidate models.

Mathematically, the incidence rate, $\lambda_{r,g}(t)$, is only constrained to be greater than zero, but numerical simulation of the system of ODEs in Equation (1) is not well constrained and negative predictions can disrupt the inference procedure. Therefore, we calculate the HIV infection probability during a single time step of duration $h$ attributable to all disease stages combined by aggregating the stage-specific rates and finding the probability of infection:

$$\lambda_{r,g}(t) = 1 - \exp\left[-h \sum_{c=1}^{4} \lambda_{r,g,c}(t)\right]. \tag{10}$$

We use this transformation to avoid numerical problems during the inference procedure; all reported region-level incidence here are per person-year.

**1.3.1.1.1 Spatio-temporal HIV transmission rates** The model allows the transmission rate of HIV to vary by time, region, sex, and transmitting CD4 category. The relative infectiousness by transmitting CD4 category is based on fixed assumptions, and the other dynamics are inferred. We model the log-transformed region-/time-specific HIV transmission rate, $\log \kappa_{r,t}$, with a hierarchical linear model:

$$\begin{aligned}
\log \kappa_{r,t} &= \kappa_0 + \kappa_r + (\gamma_0^\kappa + \gamma_r^\kappa) \cdot t + \textstyle\sum_{i=1}^{K_\kappa+1} \beta_{i,r}^\kappa \phi_i^\kappa(t) \\
\kappa_0 &\sim \mathrm{N}(0, 5) \\
\kappa_r &\sim \mathrm{N}(0, \sigma_\kappa) \\
\gamma_0^\kappa &\sim \mathrm{N}(0, 5) \\
\gamma_r^\kappa &\sim \mathrm{N}(0, \sigma_{\gamma^\kappa}) \\
\beta_{i,r}^\kappa &\sim \mathrm{ARIMA}_{\sigma_{\beta^\kappa}, \theta_{\beta^\kappa}}(1, d, 0) \\
\sigma_\kappa, \sigma_{\gamma^\kappa}, \sigma_{\beta^\kappa} &\sim \mathrm{N}^+(0, 1) \\
\operatorname{logit} \theta_{\beta_\kappa} &\sim \mathrm{N}(0, \sqrt{1/0.15}) \\
\beta_{1,r}^\kappa &= 0.
\end{aligned} \tag{11}$$

where $\kappa_0$ is a shared intercept, $\kappa_r$ is a regional intercept and $\gamma_0^\kappa$ and $\gamma_r^\kappa$ are mean and region-specific slopes with respect to time. The remainder of the model for $\kappa_{r,t}$ defines a spline model with coefficients distributed according to an autoregressive integrated moving average (ARIMA) model (Hyndman and Athanasopoulos 2018). In this model, $K_\kappa$ is the number of knots, $\beta_{i,r}^\kappa$ is a region-specific coefficient for basis function $i$ and $\phi_i^\kappa$ is the $i$'th basis function.

Returning to Equation (11), we specify that only one autoregressive term and no moving average terms may be included but do not specify the order of difference. All else equal, higher order differencing should result in a smoother curve. We assessed the choice of $d$ in the model comparison study (Section 1.5).

The model of incidence contributes the following parameters to $\theta_P$: a transmission rate sex log-ratio, $\psi$, a transmission rate intercept, $\kappa_0$, a set of regional intercepts, $\kappa_r$, a mean transmission rate slope with respect to time, $\beta_0^\kappa$, a set of regional slopes, $\beta_r^\kappa$, and two standard deviations, $\sigma_r^\kappa$ and $\sigma_{\beta^\kappa}$, which are outlined in Table 3.

**1.3.1.2 Model of ART initiation** We use a similar log-linear regression approach to model region-/time-/sex-/substage-specific ART initiation rates (an outcome similarly not observed directly):

Supplemental Table 3: Parameters used in the model of HIV transmission rates. Indexed parameters are estimated for all possible values of that index.

| Param | Size | Description | Prior |
|---|---|---|---|
| $\psi$ | 1 | Log-incidence rate sex ratio | $\mathrm{N}(0, 5)$ |
| $\upsilon$ | 1 | Log-incidence rate sex ratio slope | $\mathrm{N}(0, 5)$ |
| $\kappa_0$ | 1 | Log-transmission rate mean intercept | $\mathrm{N}(0, 5)$ |
| $\kappa_r$ | $R$ | Log-transmission rate region intercept | $\mathrm{N}(0, \sigma_\kappa)$ |
| $\sigma_\kappa$ | 1 | Log-transmission rate region intercept SD | $\mathrm{N}^+(0, 1)$ |
| $\gamma_0^\kappa$ | 1 | Log-transmission rate mean slope | $\mathrm{N}(0, 5)$ |
| $\gamma_r^\kappa$ | $R$ | Log-transmission rate mean regional slope | $\mathrm{N}(0, \sigma_{\beta^\kappa})$ |
| $\sigma_{\gamma^\kappa}$ | 1 | Log-transmission rate region slope SD | $\mathrm{N}^+(0, 1)$ |
| $\beta_{i,r}^\kappa$ | $R \times K_\kappa + 1$ | Log-transmission rate regional spline coefficient | $\mathrm{ARIMA}_{\sigma_{\beta^\kappa}, \theta_{\beta^\kappa}}(1, d, 0)$ |
| $\sigma_{\beta^\kappa}$ | 1 | Log-transmission rate region ARIMA SD | $\mathrm{N}^+(0, 1)$ |
| $\operatorname{logit} \theta_{\beta^\kappa}$ | 1 | Log-transmission rate region ARIMA autocorrelation SD | $\mathrm{N}^+(0, \sqrt{1/0.15})$ |

$$
\begin{aligned}
\log \alpha^\star_{r,t,g,c} &= \zeta_{c,g} + g \cdot \chi + \alpha^\star_0 + \alpha^\star_r + (\gamma_0^{\alpha^\star} + \gamma_r^{\alpha^\star}) \cdot t + \textstyle\sum_{i=1}^{K_\alpha+1} \beta_{i,r}^{\alpha^\star} \phi_i^{\alpha^\star}(t) \\
\alpha^\star_0 &\sim \mathrm{N}(0, 5) \\
\alpha^\star_r &\sim \mathrm{N}(0, \sigma_{\alpha^\star}) \\
\gamma_0^{\alpha^\star} &\sim \mathrm{N}(0, 5) \\
\gamma_r^{\alpha^\star} &\sim \mathrm{N}(0, \sigma_{\gamma^\alpha}) \\
\beta_{i,r}^{\alpha^\star} &\sim \mathrm{ARIMA}_{\sigma_{\beta^\alpha}, \theta_{\beta^\alpha}}(1, 2, 0) \\
\sigma_{\alpha^\star}, \sigma_{\gamma^\alpha}, \sigma_{\beta^\alpha} &\sim \mathrm{N}^+(0, 1) \\
\operatorname{logit} \theta_{\beta_\alpha} &\sim \mathrm{N}(0, \sqrt{1/0.15}) \\
\beta_{1,r}^{\alpha^\star} &= 0.
\end{aligned}
\tag{12}
$$

This model has region-specific log-linear models with respect to time with additional region-specific ARIMA error term. Here, $\zeta_{c,g}$ is a sex-/stage-specific rate of ART initiation, $\chi$ is an inferred intercept among women, $\alpha^\star_0$ is a mean intercept, $\alpha^\star_r$ is a regional intercept, $K_\alpha$ is a number of knots, $\beta_{i,r}^{\alpha^\star}$ is a regional spline coefficient, $\phi_i$ is a spline basis function, and $\sigma_{\alpha^\star}$, $\sigma_{\gamma_{\alpha^\star}}$, and $\sigma_{\beta_r^\alpha}$ are standard deviations. To prevent the autoregressive model from being rank deficient, we fix the first coefficient in the regional splines to be zero. In the results we present here, we set $\phi$ to be an order-two spline with annual knots, effectively linearly interpolating between inferred annual values.

For all $t$ before ART was scaled up in any given region, we fix $\phi_i(t)$ to be zero. The baseline ART initiation rate $\zeta_{c,g}$ is defined as $\mu^I_{c,g}/\mu^I_{1,1}$, the ratio of mortality in CD4 stage $c$ relative to women in stage 1. This encodes an assumption that PLHIV at stage $c$ initiate treatment in proportion to the expected mortality in $c$. This model of ART initiation contributes the following parameters to $\theta_P$: an intercept, $\alpha^\star_0$, a set of region random effects, $\alpha^\star_r$, set of spline coefficient means, mean and region-specific slopes, $\gamma_0^{\alpha^\star}$ and $\gamma_r^{\alpha^\star}$, a set of regional spline coefficients, $\beta_{i,r}^{\alpha^*}$, three standard deviations, $\sigma_{\alpha^*}$, $\sigma_{\gamma^\alpha}$, and $\sigma_{\beta^\alpha}$, and an autoregressive parameter $\theta_{\beta^\alpha}$, which are outlined in Table 4.

Supplemental Table 4: Parameters used in the model of ART initation. Indexed parameters are estimated for all possible values of that index.

| Param | Size | Description | Prior |
|---|---|---|---|
| $\chi$ | 1 | Log-ART initiation rate sex effect | $\mathrm{N}(0,5)$ |
| $\alpha_0^\star$ | 1 | Log-ART initiation rate mean intercept | $\mathrm{N}(0,5)$ |
| $\alpha_r^\star$ | $R$ | Log-ART initiation rate regional intercept | $\mathrm{N}(0,\sigma_{\alpha^\star})$ |
| $\sigma_{\alpha^\star}$ | 1 | Log-ART initiation rate regional intercept SD | $\mathrm{N}^+(0,1)$ |
| $\gamma_0^{\alpha^\star}$ | 1 | Log-ART initation rate mean slope | $\mathrm{N}(0,5)$ |
| $\gamma_r^{\alpha^\star}$ | $R$ | Log-ART initation rate regional slope | $\mathrm{N}(0,\sigma_{\gamma^\alpha})$ |
| $\sigma_{\gamma^\alpha}$ | 1 | Log-transmission rate region slope SD | $\mathrm{N}^+(0,1)$ |
| $\beta_{I,r}^{\alpha^\star}$ | $R{\times}K_\alpha{+}1$ | Log-ART initiation rate regional spline coefficient | $\mathrm{ARIMA}_{\sigma_{\beta^\alpha},\theta_{\beta^\alpha}}(1,2,0)$ |
| $\sigma_{\beta^\alpha}$ | 1 | Log-ART initiation rate region ARIMA SD | $\mathrm{N}^+(0,1)$ |
| $\operatorname{logit}\theta_{\beta^\alpha}$ | 1 | Log-ART initation rate region ARIMA autocorrelation SD | $\mathrm{N}^+(0,\sqrt{1/0.15})$ |
| $\delta_0$ | 1 | Mean ANC bias | $\mathrm{N}(0,5)$ |

**1.3.1.3 Model of initial state** Region-/sex-specific initial prevalence is modelled with a logit-linear model:

$$\begin{aligned} \operatorname{logit}\rho_{r,g}(0) &= \rho_r + g\cdot\epsilon \\ \rho_r &\sim \mathrm{N}(\rho_0,\sigma_\rho) \\ \rho_0 &\sim \mathrm{N}(0,5) \\ \sigma_\rho &\sim \mathrm{N}^+(0,1) \end{aligned} \tag{13}$$

where $\rho_0$ is cross-region logit-transformed mean prevalence at time 0, $\rho_r$ is a regional deviation from $\rho_0$, $\epsilon$ is an intercept for prevalence among women (recalling that $g=1$ among women), and $\sigma_\rho$ is a standard deviation for the random effects. We calculate $\epsilon$ from Spectrum estimates as the log-ratio of female prevalence to male prevalence.

To maintain consistency with other national-level estimates of prevalence, we put a prior on initial prevalence among men:

$$\begin{aligned} \hat{\rho}_{\mathrm{Nat}} &\sim \mathrm{N}(\rho_{\mathrm{Nat}},0.005) \\ \hat{\rho}_{\mathrm{Nat}} &= \frac{1}{P_{\mathrm{Nat},0}(0)}\sum_{r=1}^{R}\frac{P_{r,0}(0)}{1+\exp(-\rho_{r,0})}, \end{aligned} \tag{14}$$

where $P_{r,0}(0)$ is initial male population in region $r$ and $P_{r,0}(0)/(1+\exp(-\rho_{r,0}))$ is estimated male PLHIV in region $r$. This prior encourages the model to match external estimates of initial prevalence, without sacrificing subnational variation. The inverse logit-transformed mean of the random effects, $1/(1+\exp(-\rho_0))$, cannot be compared directly to exogenous initial prevalence $\rho_{\mathrm{Nat}}$, because $\rho_{\mathrm{Nat}}$ is implicitly population-weighted.

We assume that $t=0$ is before ART scale-up, so $A_{r,g,c}(0)=0$ in all cases. Making fixed assumptions about the distribution of PLHIV across disease substage without treatment, we calculate $I_{r,g,c}(0)$ and solve for $S_{r,g,c}(0)$:

Supplemental Table 5: Parameters used in the model of the initial state. Indexed parameters are estimated for all possible values of that index.

| Param | Size | Description | Prior |
|---|---|---|---|
| $\rho_0$ | 1 | Initial mean prevalence | $\mathrm{N}(0, 5)$ |
| $\rho_r$ | $R$ | Initial regional prevalence | $\mathrm{N}(0, \sigma_\rho)$ |
| $\sigma_\rho$ | 1 | Initial prevalence SD | $\mathrm{N}^+(0, 1)$ |

$$\begin{aligned} I_{r,g,c}(0) &= b_{g,c} \cdot \rho_{r,g}(0) \cdot P_{r,g}(0) \\ S_{r,g}(0) &= P_{r,g}(0) - \textstyle\sum_{c=1}^{4}(I_{r,g,c}(0)) \\ A_{r,g,c}(0) &= 0. \end{aligned} \tag{15}$$

where $b_{g,c}$ is the share of PLHIV of sex $g$ in CD4 stage $c$ at time zero derived from Spectrum model estimates. $P_{r,g}(0)$ is the population at time zero for sex $g$ in region $r$, which is assumed to be a fixed known input.

This model adds the following parameters to $\theta_P$: a national mean, $\rho_0$, regional deviations from the means, $\rho_r$, and one standard deviation, $\sigma_\rho$, which are outlined in Table 5.

#### 1.4 Observation model

##### 1.4.1 Household surveys

We assume that national household surveys are probability random samples within each region, so if $s$ is a household survey, $Y^{s,\mathrm{HIV}}_{r,t,g}/T^{s,\mathrm{HIV}}_{r,t,g}$, provides an unbiased estimate of true prevalence in demographic segment $\{r, t, g\}$ where $Y$ and $T$ are the design-weighted effective count and effective sample size, respectively. We therefore assume that $Y^{s,\mathrm{HIV}}_{r,t,g}$ is a sample from a binomial distribution with $T^{s,\mathrm{HIV}}_{r,t,g}$ trials each with a probability of $\rho_{r,g}(t)$:

$$Y^{s,\mathrm{HIV}}_{r,t,g} \sim \mathrm{Binom}(T^{s,\mathrm{HIV}}_{r,t,g}, \rho_{r,g}(t)). \tag{16}$$

Defining a binomial distribution using the effective count and effective sample size is a computationally efficient way to approximate the effect of the complex multi-stage survey design (Chen, Wakefield, and Lumley 2014) and is increasingly common in recent HIV mapping exercises (Eaton et al. 2021; Dwyer-Lindgren et al. 2019).

We make a similar assumption about survey-estimated ART coverage:

$$Y^{s,\mathrm{ART}}_{r,t,g} \sim \mathrm{Binom}(T^{s,\mathrm{ART}}_{r,t,g}, \alpha_{r,g}(t)). \tag{17}$$

HIV recent infection assays (or "recency assays") indicate whether an individual was infected in the recent past, so estimated incidence and prevalence must be combined to estimate the proportions that are recent. We use the estimator from Kassanjee, McWalter, and Welte (2014) as modified by Eaton et al. (2021) to find this proportion:

$$\nu_{r,g}(t) = \frac{\lambda_{r,g}(t) \cdot (1 - \rho_{r,g}(t)) \cdot (\Omega_R - \gamma_R) + \gamma_R \rho_{r,g}(t)}{\rho_{r,g}(t)}, \tag{18}$$

where $\Omega_R$ is the mean duration of recent infection (fixed at 130/365), and $\gamma_R$ is the proportion of positive recency assays that are false positives (fixed at 0). As before, we assume that each $Y_{r,t,g}^{s,\mathrm{Rec}}$ is a sample from a binomial distribution:

$$Y_{r,t,g}^{s,\mathrm{Rec}} \sim \mathrm{Binom}(T_{r,t,g}^{s,\mathrm{Rec}}, \nu_{r,g}(t)). \tag{19}$$

Because the mean duration of recent infection and recency assay false positive rate are fixed, the observation models for survey data do not contribute any parameters to the model.

##### 1.4.2 ANC facility data

HIV prevalence among ANC attendees is not representative of HIV prevalence among the general population, so we cannot estimate $\rho_{r,1}(t)$ with $Y_{r,t,1}^{s,\mathrm{HIV}}/T_{r,t,1}^{s,\mathrm{HIV}}$. Instead, following Bao (2012), we estimate site-specific ANC prevalence as a function of general population prevalence and facility effects

$$\begin{aligned}
\operatorname{logit} \rho_{r,1}^{s}(t) &= \operatorname{logit} \rho_{r,1}(t) + \delta_0 + \delta_s + (\epsilon_0 + \epsilon_s) \cdot t \\
\delta_0, \epsilon_0 &\sim \mathrm{N}(0, 5) \\
\delta_s &\sim \mathrm{N}(0, \sigma_\delta) \\
\epsilon_s &\sim \mathrm{N}(0, \sigma_\epsilon) \\
\sigma_\delta, \sigma_\epsilon &\sim \mathrm{N}(0, 1),
\end{aligned} \tag{20}$$

where $\delta_s$ is a facility-specific random effect, $\delta_0$ is a mean ANC offset, $\epsilon_0$ is a mean slope with respect to time, $\epsilon_s$ is a site-specific slope, and $\sigma_\delta$ and $\sigma_\epsilon$ are standard deviations for the site-specific parameters. Bao (2012) do not include slopes with respect time in their ANC observation. Eaton et al. (2014) found that we cannot assume that the representativeness of ANC facilities is not changing.

Eaton and Bao (2017) report that Gaussian approximations to standard binomial models do not offer adequate posterior predictive coverage when fit to HIV prevalence data from ANC facilities, so this work includes the option to use one of two possible likelihoods. The first is a standard binomial model

$$Y_{r,t,1}^{s,\mathrm{HIV}} \sim \mathrm{Binom}(T_{r,t,1}^{s,\mathrm{HIV}}, \rho_{r,1}^{s}(t)), \tag{21}$$

and the second is a beta-binomial model

Supplemental Table 6: Parameters used in the model of the initial state. Indexed parameters are estimated for all possible values of that index.

| Param | Size | Description | Prior |
|---|---|---|---|
| $\delta_s$ | $S$ | Site-specific ANC bias | $\mathrm{N}(0, \sigma_\delta)$ |
| $\sigma_\delta$ | 1 | ANC bias SD | $\mathrm{N}^+(0, 1)$ |
| $\epsilon_0$ | 1 | Mean ANC slope | $\mathrm{N}(0, 5)$ |
| $\epsilon_s$ | $S$ | Site-specific ANC bias slope | $\mathrm{N}(0, \sigma_\epsilon)$ |
| $\phi_{\text{type}[s]}$ | 2 | Type-specific ANC beta-binomial overdispersion | $\mathrm{N}(-1, 1)$ |
| $\sigma_\epsilon$ | 1 | ANC bias slope SD | $\mathrm{N}^+(0, 1)$ |

$$
\begin{aligned}
Y_{r,t,1}^{s,\text{HIV}} &\sim \text{BetaBinom}(T_{r,t,1}^{s,\text{HIV}}, \rho_{r,1}^{s}(t), \phi_{\text{type}[s]}) \\
\text{logit}\, \phi_{\text{type}[s]} &\sim \mathrm{N}(-1, 1)
\end{aligned}
\tag{22}
$$

where $\phi_{\text{type}[s]} \in (0, 1)$ is a parameter measuring the autocorrelation between each Bernoulli trial. In the beta-binomial case we estimate two separate values of $\phi$: one when $s$ is an individual facility and one when $s$ is an aggregate over multiple facilities. We evaluate the effect of the choice of ANC observation in the model specification study. This model contributes the following parameters to the model: coefficients for the observation model, $\delta_0$, $\delta_s$, $\epsilon_0$, and $\epsilon_s$, and hyperparameters, $\sigma_\delta$, $\sigma_\epsilon$, and $\phi_{\text{type[s]}}$, which are outlined in Table 6.

##### 1.4.3 ART programme data

The final data source used by the model is programmatic ART patient count time series. We use $C_{r,t}$ to denote the total number of adults receiving ART in region $r$ at the end of time $t$. The compartmental model produces estimates of the number of PLHIV living in $r$ that are on treatment, $A_r(t)$, but these estimates are not directly comparable to the corresponding $C_{r,t}$. While large surveys measure individuals in their regions of residence, ART programme data record individuals where they seek treatment. Because we fit directly to survey data and use population estimates defined by residency, we are implicitly modelling individuals in their regions-of-residence, and therefore need to adjust $A_r(t)$ for treatment-seeking dynamics before it can be compared directly to $C_{r,t}$.

Following Eaton et al. (2021), we model the number of PLHIV seeking treatment in region $r$ at time $t$ as

$$
A_r^*(t) = \sum_{\{j \sim r\}} \pi_{j \to r,t} A_j(t), \tag{23}
$$

where $\{j \sim r\}$ is set of regions that are adjacent to $r$ inclusive of $r$, $\pi_{j \to r,t}$ is the time-varying probability an individual residing in $j$ will seek treatment in $r$, and $A_j(t)$ is the number of PLHIV on ART who live in $j$. Note that $A_j(t) = \sum_{g \in \{0,1\}} A_{j,g}(t)$.

We model the odds of moving from $j$ to $r$ relative to staying in $r$ as

$$
\begin{aligned}
\log \frac{\pi_{r\to j,t}}{\pi_{r\to r,t}} &= \log \mu_{r\to j,t} = m_j + m_0 + \beta_\pi t \\
m_j &\sim \mathrm{N}(0, \sigma_m^2) \\
m_0 &\sim \mathrm{N}(-3, 1) \\
\beta_\pi &\sim \mathrm{N}(0, 5) \\
\sigma_m &\sim \mathrm{N}^+(0, 2),
\end{aligned} \tag{24}
$$

where $m_j$ is a region-specific "mass" term, $m_0$ is a mean mass with a prior that ensures that most people will stay in their home regions, and $\beta_\pi$ is a time-specific slope. Following the Naomi model, we place an informative prior on $m_0$ that assumes *a priori* that the majority of people seek treatment in their region of residence. If $m_j$ and $\beta_\pi$ are both fixed to be zero and region $r$ has one neighbour, then $m_0 = -3.0$ implies that approximately 95% of individuals residing in $r$ will seek treatment in $r$.

We allow each $\pi$ to vary with respect to time to account for national-level changes in ART programmes; across-the-board improvements in treatment provision could result in fewer patients needing to travel to receive adequate care and therefore, a negative value of $\beta_\pi$. Naomi was designed to estimate recent trends, so it covers a much shorter time period and does not need to account for long-term changes in ART programmes.

We use the softmax function to solve for $\pi_{r\to j,t}$ with $\mu_{r\to j,t} = 1.0$:

$$
\pi_{r\to j,t} = \frac{\mu_{r\to j,t}}{1 + \sum_{\{k\sim r\}\setminus r} \mu_{r\to k,t}}. \tag{25}
$$

Then we find $\pi_{r\to r,t} = 1 - \sum_{\{k\sim r\}\setminus r} \pi_{r\to k,t}$.

These data do not have a natural denominator, so we cannot treat them as independent binomial samples. Instead, we use a negative binomial model with variance that scales both linearly and quadratically with its mean (Lindén and Mäntyniemi 2011). Let

$$
\begin{aligned}
\mu &= A_r^*(t) \\
\sigma^2 &= \mu + \theta_1 \mu + \theta_2 \mu^2,
\end{aligned} \tag{26}
$$

where $\theta_1, \theta_2 > 0$. We can use $\mu$ and $\sigma^2$ to find the typical negative binomial parameters: $r = \mu^2/(\sigma^2 - \mu)$ and $p = \mu/\sigma^2$. Then we have

$$
C_{r,t} \sim \mathrm{NegBinom}(r, p). \tag{27}
$$

For a fixed value of $\theta_2$, as $\theta_1$ goes to zero, this distribution converges to a negative binomial with overdispersion $\theta_2$. Conversely, for a fixed value of $\theta_1$, as $\theta_2$ goes to zero, it goes to a quasi-Poisson distribution. As both $\theta_1$ and $\theta_2$ go to zero, it returns to Poisson.

Supplemental Table 7: Parameters used in the ART patint count observation model Indexed parameters are estimated for all possible values of that index.

| Param | Size | Description | Prior |
|---|---|---|---|
| $m_0$ | 1 | Mean ART attraction mass | $\mathrm{N}(0, 5)$ |
| $m_r$ | $R$ | Regional ART attraction mass | $\mathrm{N}(0, \sigma_m)$ |
| $\sigma_m$ | 1 | Regional ART attraction mass SD | $\mathrm{N}^{+}(0, 1)$ |
| $\beta_\pi$ | 1 | ART attraction slope | $\mathrm{N}(0, 5)$ |
| $\omega$ | 1 | Linear overdispersion term | $\mathrm{N}^{+}(0, 2)$ |
| $\theta$ | 1 | Quadratic overdispersion term | $\mathrm{N}^{+}(0, 2)$ |

Allowing the variance of $C_{r,t}$ to scale both linearly and quadratically with $\mu$ allows this model to scale appropriately across regions of varying sizes. A one-unit change in $\theta_2$ will result in a much larger change in variance in a high-population region than in a low-population region, even though we do not necessarily expect the measurement of ART patients to be quadratically higher variance in the high-population region.

We set the following priors on $\theta_1$ and $\theta_2$:

$$\log \theta_1, \log \theta_2 \sim \mathrm{N}(0, 2) \tag{28}$$

The observation model for ART patient counts contributes the following parameters to the full set of parameters: $\theta_1$ and $\theta_2$ from the quasi-negative binomial distribution, one mass, $m_r$, per region, a mean mass, $m_0$, a time coefficient, $\beta_t$, and one variance $\sigma_m^2$, which are outlined in Table 7.

#### 1.5 Model selection methods

Because incidence is not measured directly, it is not possible to directly cross-validate model results against withheld observations of incidence, the main outcome of interest. Cross-validation simulates how well a model generalises to new data, so we have designed a strategy focused on forecasting the data sources we expect to continue to acquire (Vollmer et al. 2021). To that end, we evaluated the model's performance on Malawian data using a cross-validation strategy predicts on routinely reported indicators, represented in my model by ART programme data and facility-based ANC prevalence. We constructed cross-validation datasets by holding out all data after one of six forecasting horizons: the first of January in 2015, 2016, 2017, 2018, 2019, and 2020. We compared each model using out-of-sample root mean squared error (RMSE) with respect to observed point estimates and 50%, 80%, and 95% posterior predictive coverage separately for the two datasets (ANC facility data and ART programme data).

##### 1.5.1 Model configurations

We tested every combination of choices for seven different design decisions, outlined in Table 8. First, we allowed the likelihood for the ANC facility data to be either binomial or beta-binomial. As described in Section 1.4.2, individual facility series shared one autocorrelation parameter and aggregate series

Supplemental Table 8: Model configuration variables tested in this chapter with descriptions of each value. Unless otherwise specified, every component refers to the transmission rate model.

| Variable | Value | Description |
|---|---|---|
| ANC observation model | Binomial | Binomial ANC observation model |
| | Beta-binomial | Beta-binomial ANC observation model |
| ARIMA order | 1 | One degree of ARIMA differencing |
| | 2 | Two degrees of ARIMA differencing |
| | 3 | Three degrees of ARIMA differencing |
| Include slope | Yes | Exclude slope w.r.t. time |
| | No | Include slope w.r.t. time |
| Spline interval | 1 | Knots at one-year intervals |
| | 5 | Knots at five-year intervals |
| Spline order | 1 | Piecewise constant design matrix |
| | 2 | Piecewise linear design matrix |
| | 3 | Order-three design matrix |
| Transm. rate model | Constant | Constant w.r.t. time |
| | Linear | Linear w.r.t. time |
| | Latent | Include ARIMA component |
| Use AR | Yes | Include autoregressive term |
| | No | Exclude autoregressive term |

shared another under the beta-binomial model. Second, we tested the value of including a non-linear district-level temporal component in the transmission rate model by fitting Equation (11) with intercepts only, intercepts and linear slopes with respect to time, and intercepts, slopes, and latent components. Among the models with latent components for the HIV transmission rate, we tested the effects of excluding the linear slope with respect to time, the order of the spline basis functions (one, two, or three), the distance between knots in the spline design matrices (one year or five years), the order of ARIMA differencing, and, finally, whether to include an autoregressive term. All valid combinations of these choices resulted in 146 different models, which led to 876 models to fit when combined with the six forecasting horizons. Each of these models were fit using the approximate inference strategy described by Skaug and Fournier (2006) and implemented by Kristensen et al. (2016). In the main text, we present only results from the final, selected model specification.

### 2 Model selection results

We summarise the results of the model specification study here and refer readers to Chapter 3 of Wolock (2022) for a detailed investigation. This study offered a few clear conclusions. First, the beta-binomial model for ANC data offered distinctly better out-of-sample fit to both the ANC data and the ART programme data than the standard binomial model (Supplemental Figure 3). Second, restricting to only models that used a beta-binomial observation model for the ANC data, none of model configuration decisions from Supplemental Table 8 resulted in superior out-of-sample fit. Instead, we were obliged to use more subjective criteria to identify a final model. Supplemental Figure 4 shows that two configurations that included a linear term with respect to time in the transmission rate model resulted in decreasing sex ratios of incidence, which conflicts with widely available evidence (Risher et al. 2021). In the end, we selected a transmission rate model with no linear term with respect to time, one degree of differencing, an order-two spline with five-year intervals between knots, and no autoregressive term.

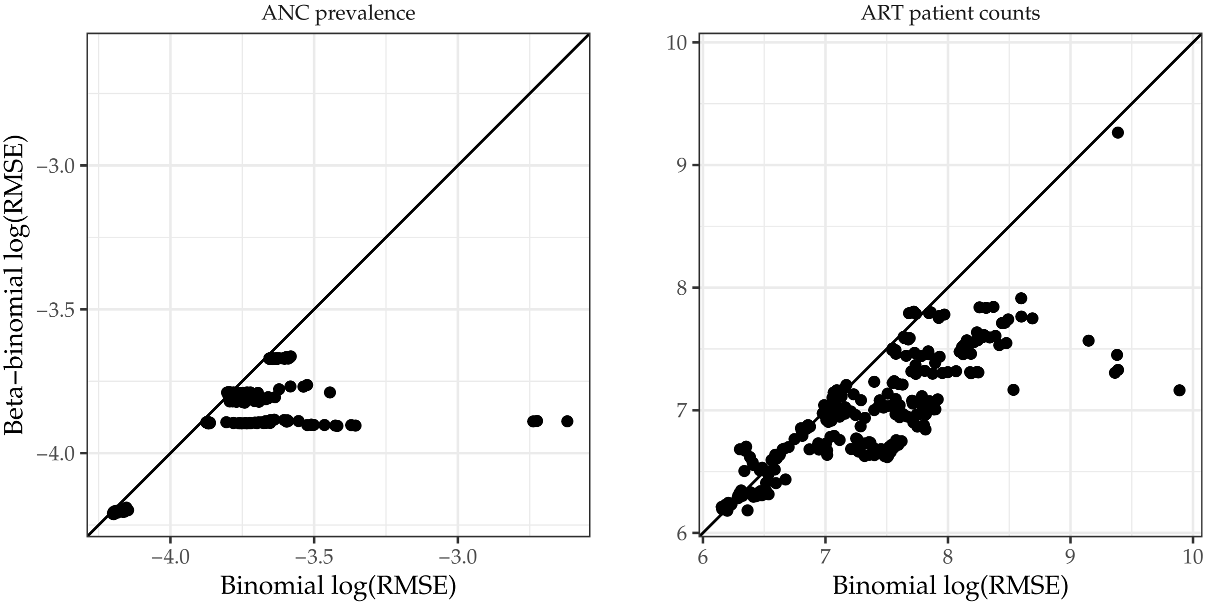

Supplemental Figure 3: Scatter plots of log-transformed out-of-sample RMSE for model configuration pairs that differ only in ANC observation model by dataset. The black line is equality. Points below the line of equality indicate that the beta-binomial observation model was lower and vice versa.

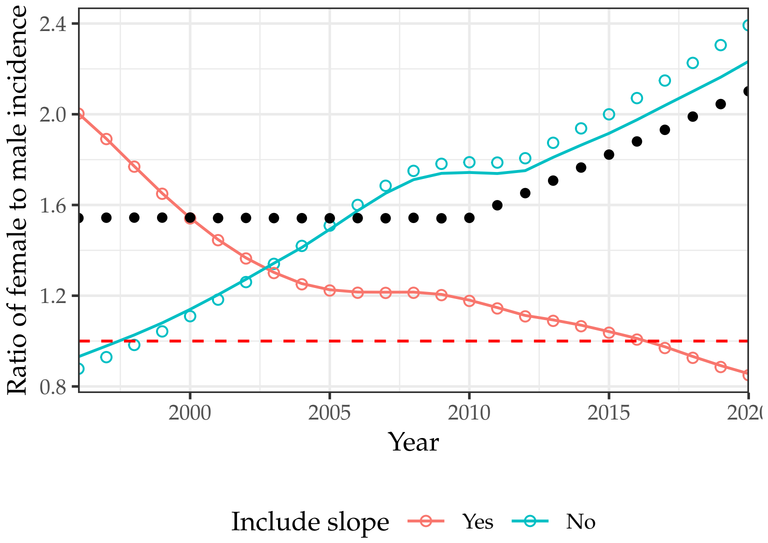

Supplemental Figure 4: The ratio of incidence among women to that among men from four models compared to UNAIDS. Lines correspond models without autoregressive terms, and open circles correspond to models with autoregressive terms. Black points are UNAIDS assumptions.

### 3 Fit in all districts

#### Chitipa District

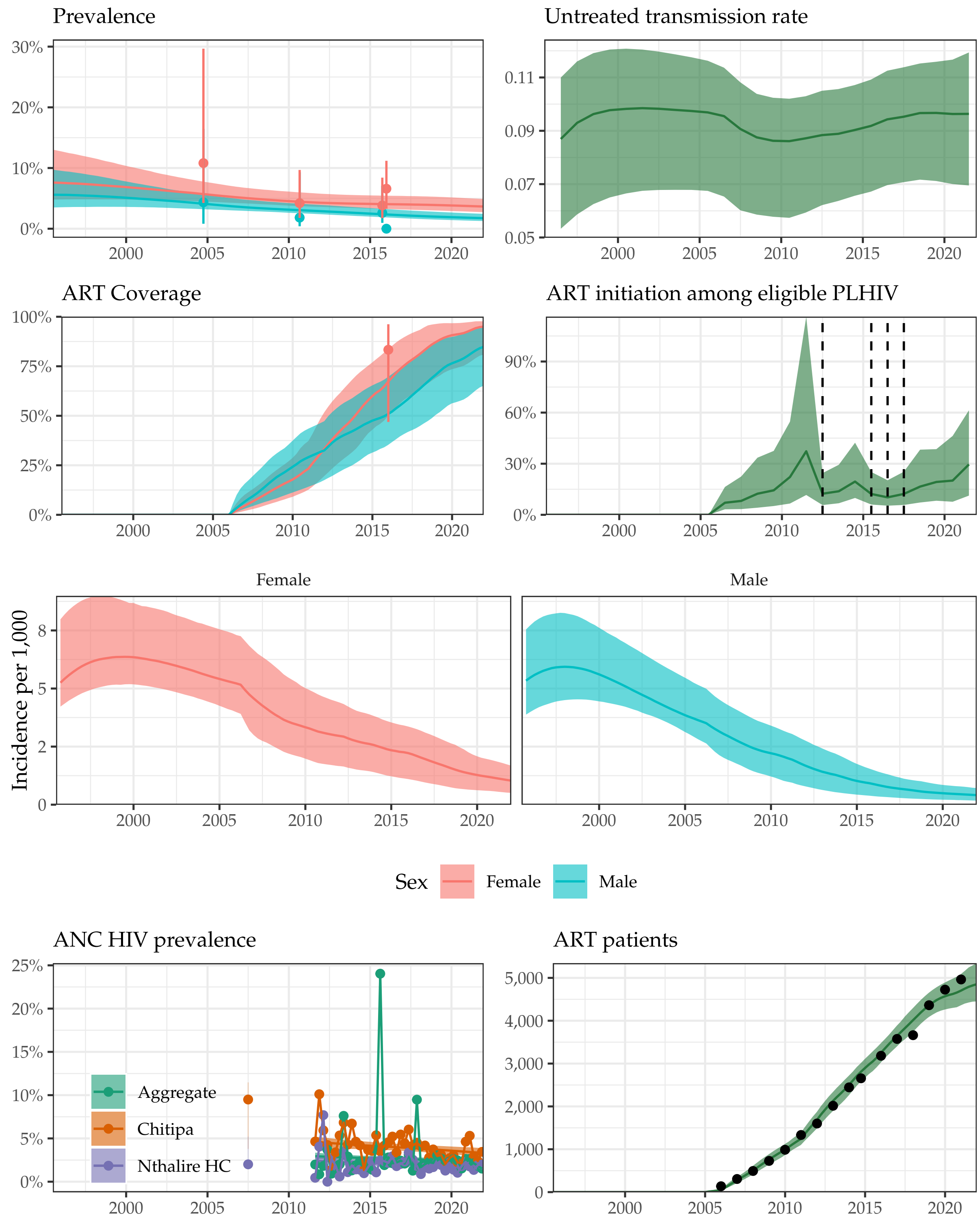

Supplemental Figure 5: **Model fit to HIV data sources in Chitipa District, 1995-2021.** Estimated prevalence, ART coverage, untreated transmission rates, annual ART initiation probabilities, ANC prevalence, and ART patient counts in the Blantyre district in southern Malawi with household survey data (HIV prevalence and ART coverage), HIV prevalence among pregnant women attending ANC facilities, and the number of adults 15-49 receiving ART programmatic reporting data (points). Prevalence, ART coverage, incidence rate, and ART patients reflect adults aged 15-49 years. Vertical dashed lines indicate years of ART eligibility changes. Different colours on panel "ANC prevalence" indicate different ANC facilities.

#### Karonga District

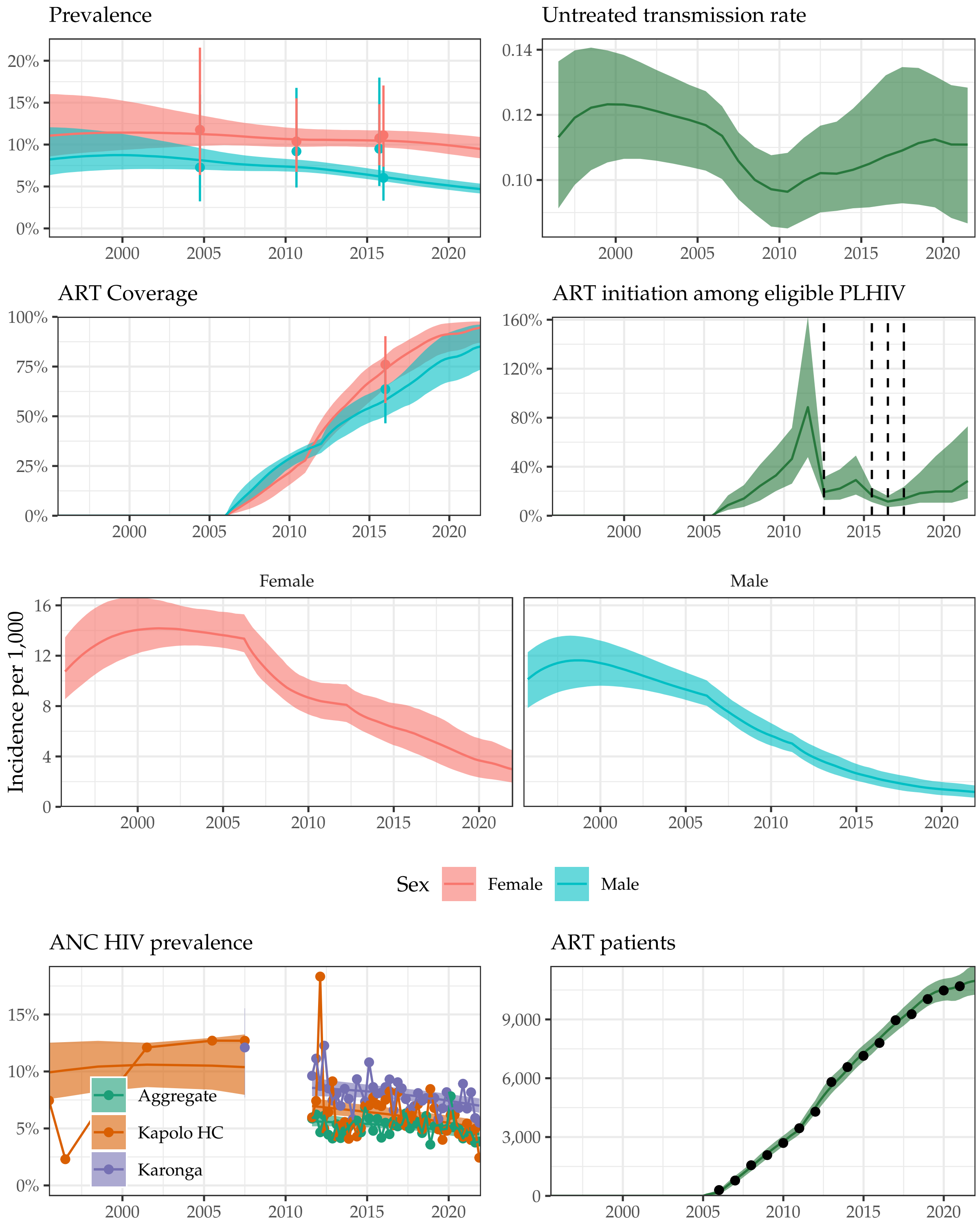

Supplemental Figure 6: **Model fit to HIV data sources in Karonga District, 1995-2021.** Estimated prevalence, ART coverage, untreated transmission rates, annual ART initiation probabilities, ANC prevalence, and ART patient counts in the Blantyre district in southern Malawi with household survey data (HIV prevalence and ART coverage), HIV prevalence among pregnant women attending ANC facilities, and the number of adults 15-49 receiving ART programmatic reporting data (points). Prevalence, ART coverage, incidence rate, and ART patients reflect adults aged 15-49 years. Vertical dashed lines indicate years of ART eligibility changes. Different colours on panel "ANC prevalence" indicate different ANC facilities.

#### Likoma District

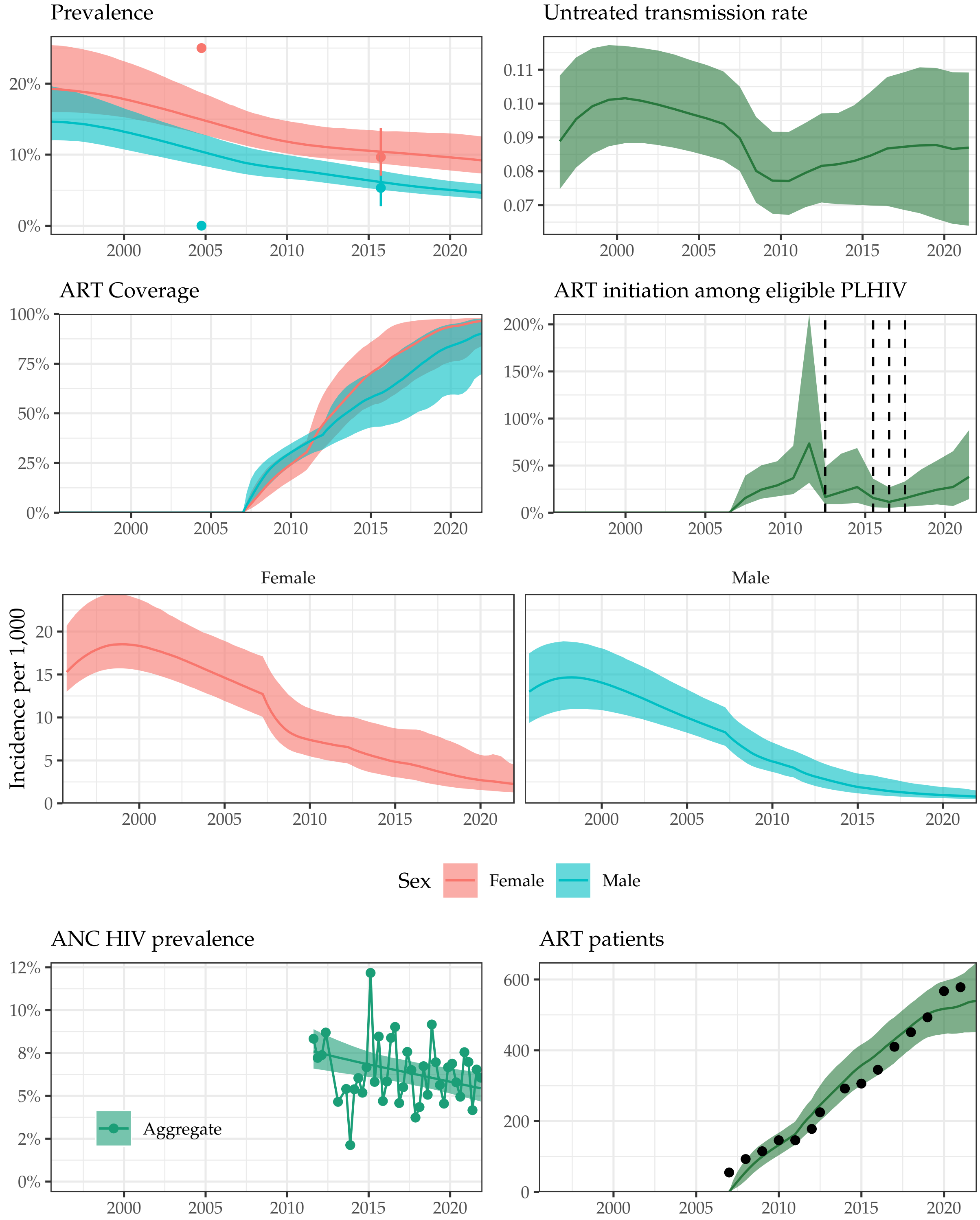

Supplemental Figure 7: **Model fit to HIV data sources in Likoma District, 1995-2021.** Estimated prevalence, ART coverage, untreated transmission rates, annual ART initiation probabilities, ANC prevalence, and ART patient counts in the Blantyre district in southern Malawi with household survey data (HIV prevalence and ART coverage), HIV prevalence among pregnant women attending ANC facilities, and the number of adults 15-49 receiving ART programmatic reporting data (points). Prevalence, ART coverage, incidence rate, and ART patients reflect adults aged 15-49 years. Vertical dashed lines indicate years of ART eligibility changes. Different colours on panel "ANC prevalence" indicate different ANC facilities.

#### Mzimba District

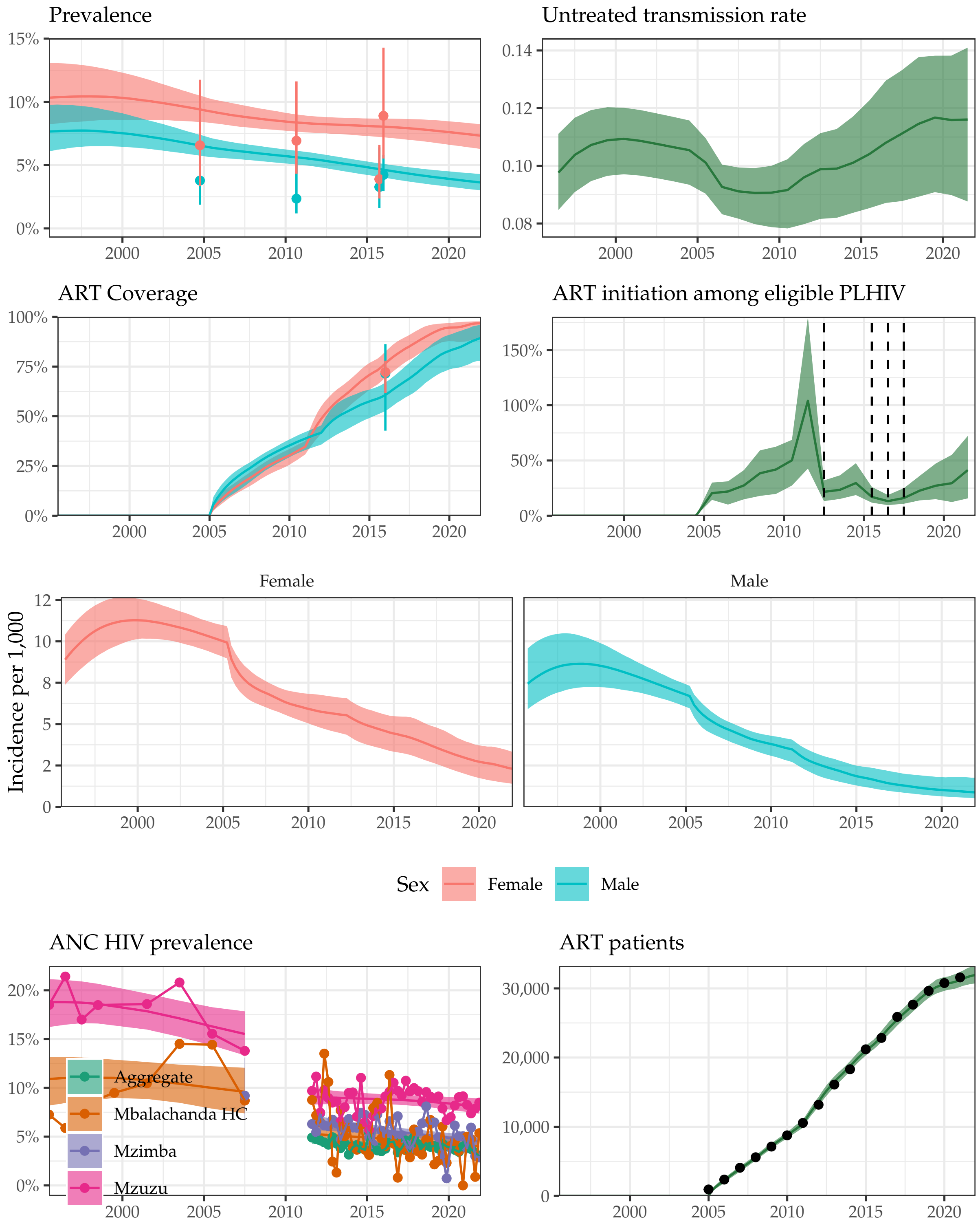

Supplemental Figure 8: **Model fit to HIV data sources in Mzimba District, 1995-2021.** Estimated prevalence, ART coverage, untreated transmission rates, annual ART initiation probabilities, ANC prevalence, and ART patient counts in the Blantyre district in southern Malawi with household survey data (HIV prevalence and ART coverage), HIV prevalence among pregnant women attending ANC facilities, and the number of adults 15-49 receiving ART programmatic reporting data (points). Prevalence, ART coverage, incidence rate, and ART patients reflect adults aged 15-49 years. Vertical dashed lines indicate years of ART eligibility changes. Different colours on panel "ANC prevalence" indicate different ANC facilities.

### Nkhata Bay District

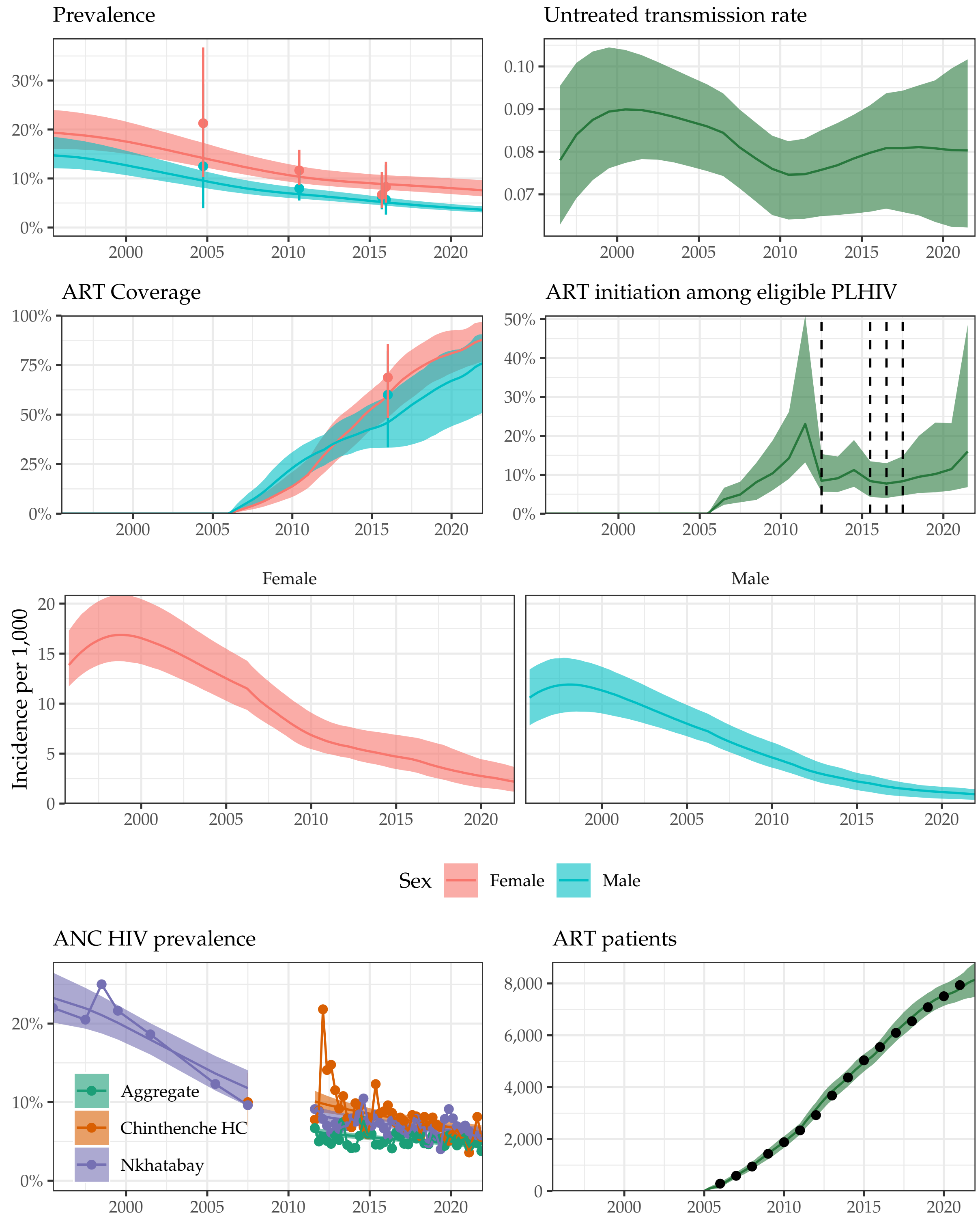

Supplemental Figure 9: **Model fit to HIV data sources in Nkhata Bay District, 1995-2021.** Estimated prevalence, ART coverage, untreated transmission rates, annual ART initiation probabilities, ANC prevalence, and ART patient counts in the Blantyre district in southern Malawi with household survey data (HIV prevalence and ART coverage), HIV prevalence among pregnant women attending ANC facilities, and the number of adults 15-49 receiving ART programmatic reporting data (points). Prevalence, ART coverage, incidence rate, and ART patients reflect adults aged 15-49 years. Vertical dashed lines indicate years of ART eligibility changes. Different colours on panel "ANC prevalence" indicate different ANC facilities.

#### Rumphi District

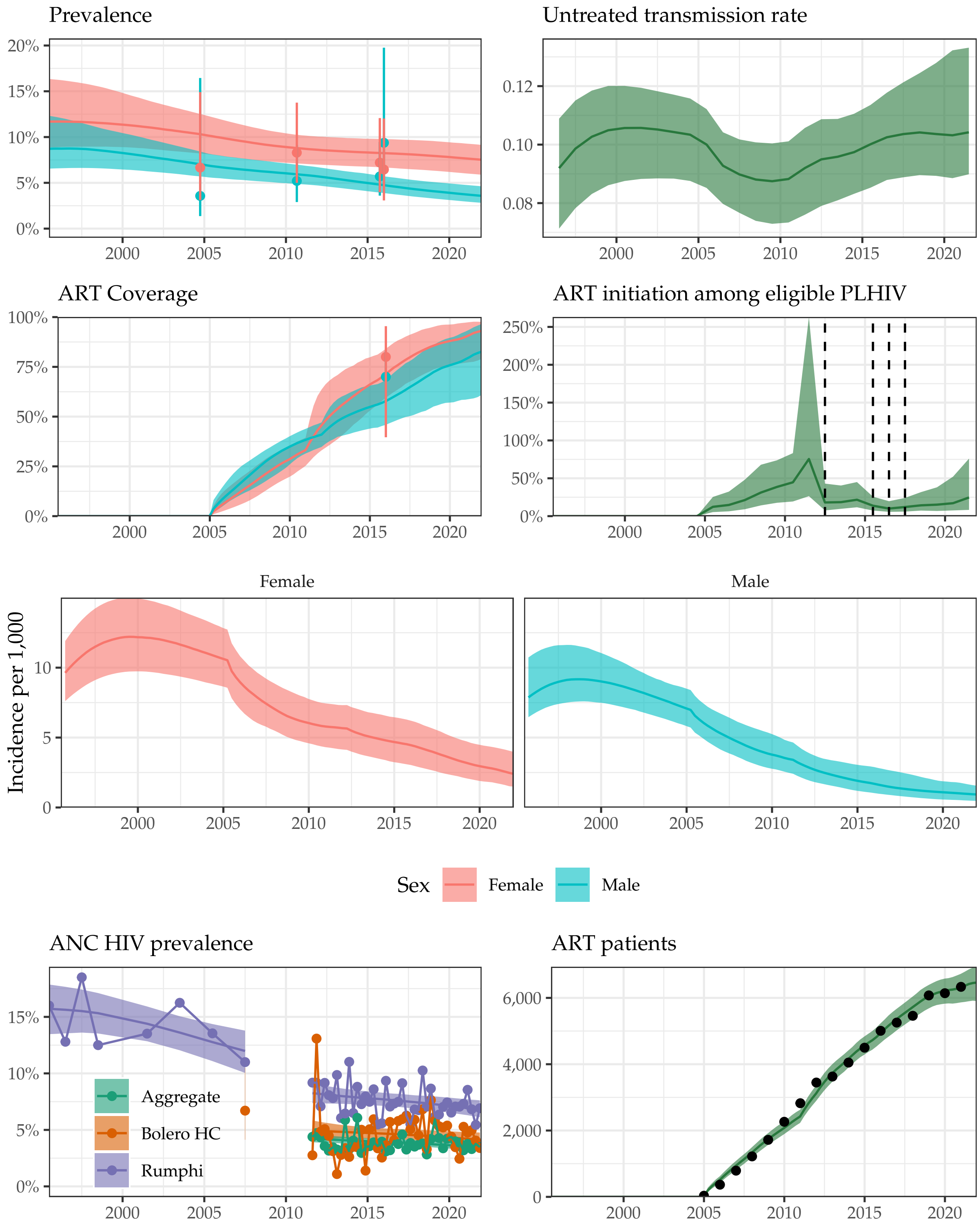

Supplemental Figure 10: **Model fit to HIV data sources in Rumphi District, 1995-2021.** Estimated prevalence, ART coverage, untreated transmission rates, annual ART initiation probabilities, ANC prevalence, and ART patient counts in the Blantyre district in southern Malawi with household survey data (HIV prevalence and ART coverage), HIV prevalence among pregnant women attending ANC facilities, and the number of adults 15-49 receiving ART programmatic reporting data (points). Prevalence, ART coverage, incidence rate, and ART patients reflect adults aged 15-49 years. Vertical dashed lines indicate years of ART eligibility changes. Different colours on panel "ANC prevalence" indicate different ANC facilities.

#### Dedza District

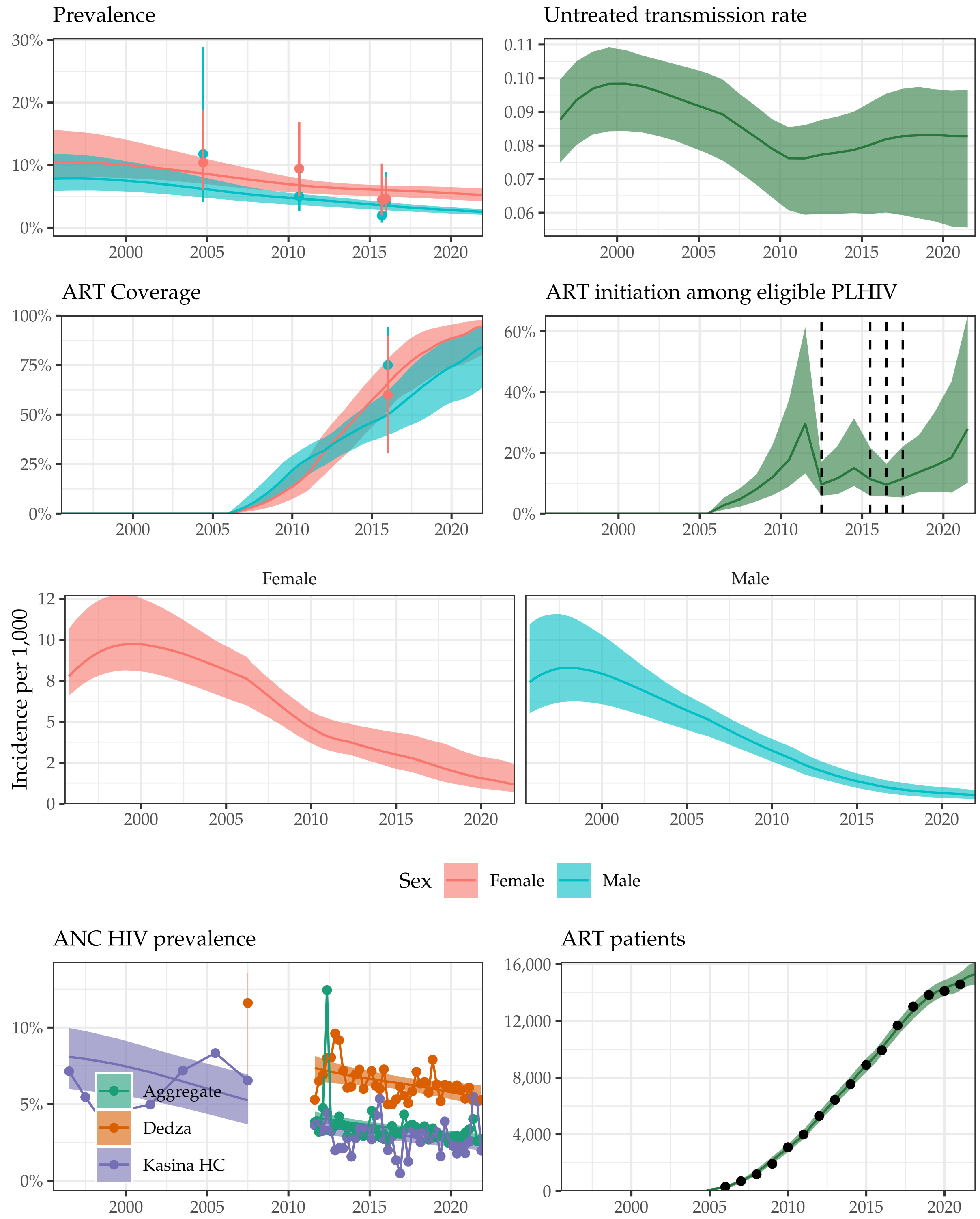

Supplemental Figure 11: **Model fit to HIV data sources in Dedza District, 1995-2021.** Estimated prevalence, ART coverage, untreated transmission rates, annual ART initiation probabilities, ANC prevalence, and ART patient counts in the Blantyre district in southern Malawi with household survey data (HIV prevalence and ART coverage), HIV prevalence among pregnant women attending ANC facilities, and the number of adults 15-49 receiving ART programmatic reporting data (points). Prevalence, ART coverage, incidence rate, and ART patients reflect adults aged 15-49 years. Vertical dashed lines indicate years of ART eligibility changes. Different colours on panel “ANC prevalence” indicate different ANC facilities.

#### Dowa District

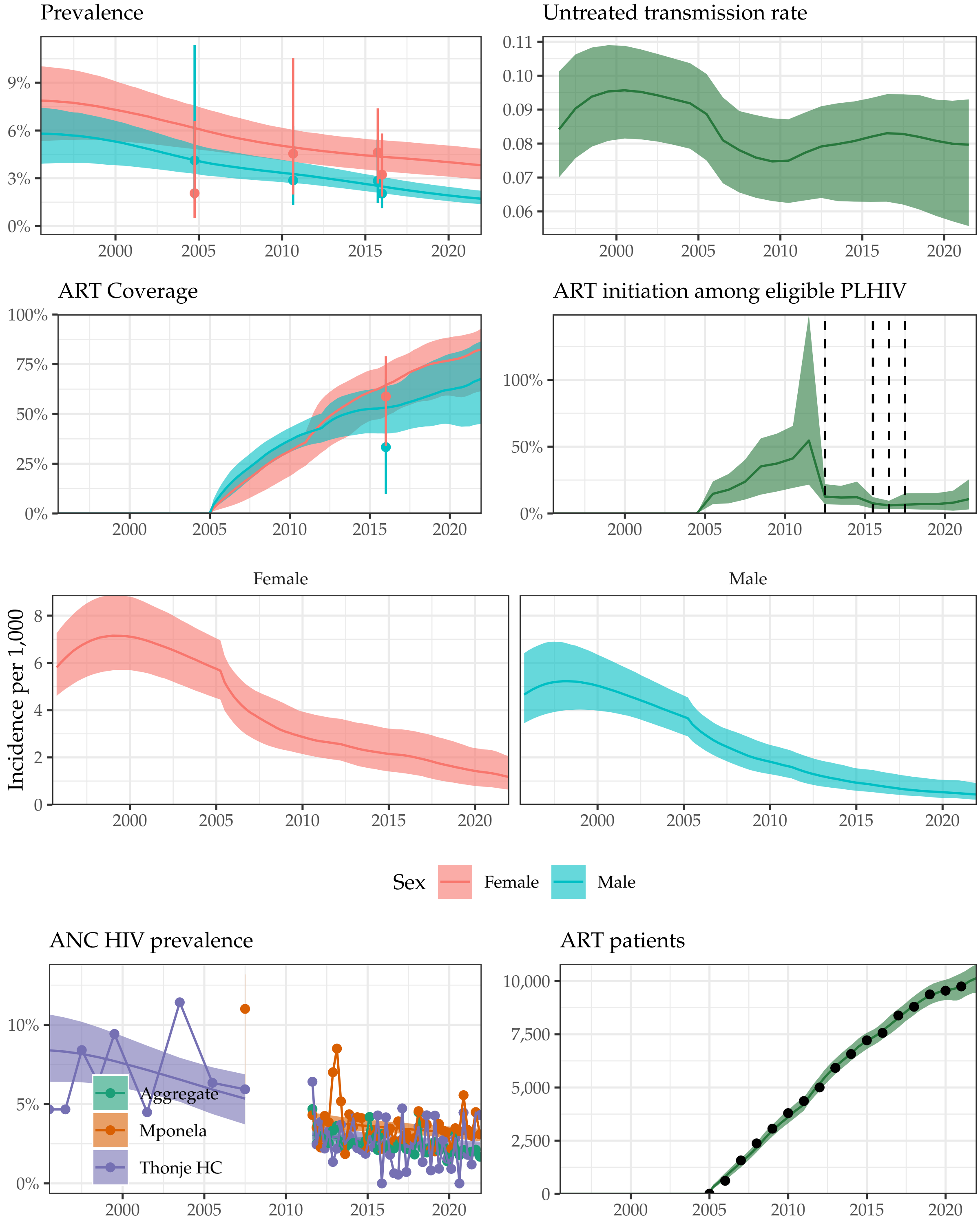

Supplemental Figure 12: **Model fit to HIV data sources in Dowa District, 1995-2021.** Estimated prevalence, ART coverage, untreated transmission rates, annual ART initiation probabilities, ANC prevalence, and ART patient counts in the Blantyre district in southern Malawi with household survey data (HIV prevalence and ART coverage), HIV prevalence among pregnant women attending ANC facilities, and the number of adults 15-49 receiving ART programmatic reporting data (points). Prevalence, ART coverage, incidence rate, and ART patients reflect adults aged 15-49 years. Vertical dashed lines indicate years of ART eligibility changes. Different colours on panel "ANC prevalence" indicate different ANC facilities.

#### Kasungu District

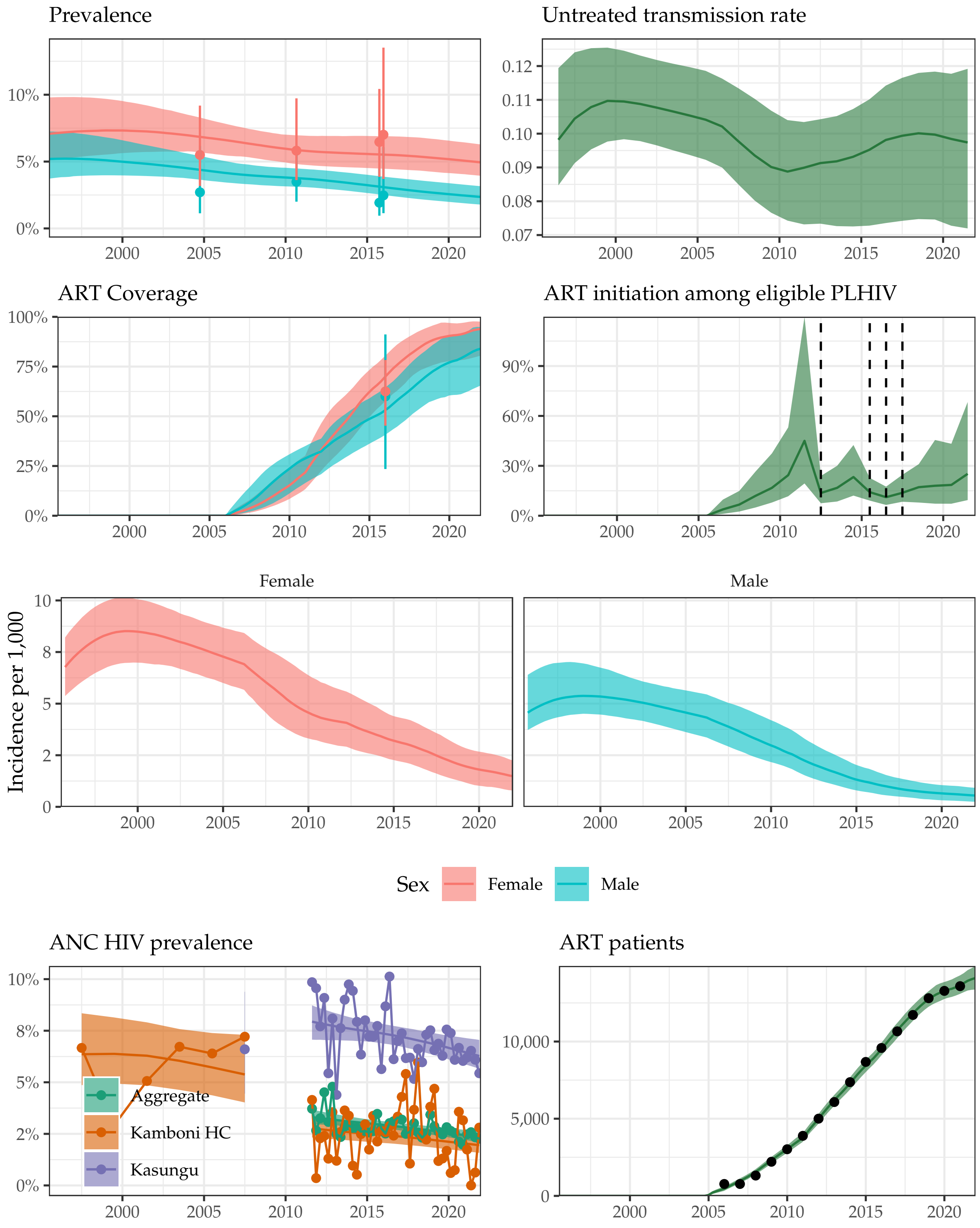

Supplemental Figure 13: **Model fit to HIV data sources in Kasungu District, 1995-2021.** Estimated prevalence, ART coverage, untreated transmission rates, annual ART initiation probabilities, ANC prevalence, and ART patient counts in the Blantyre district in southern Malawi with household survey data (HIV prevalence and ART coverage), HIV prevalence among pregnant women attending ANC facilities, and the number of adults 15-49 receiving ART programmatic reporting data (points). Prevalence, ART coverage, incidence rate, and ART patients reflect adults aged 15-49 years. Vertical dashed lines indicate years of ART eligibility changes. Different colours on panel "ANC prevalence" indicate different ANC facilities.

#### Lilongwe District

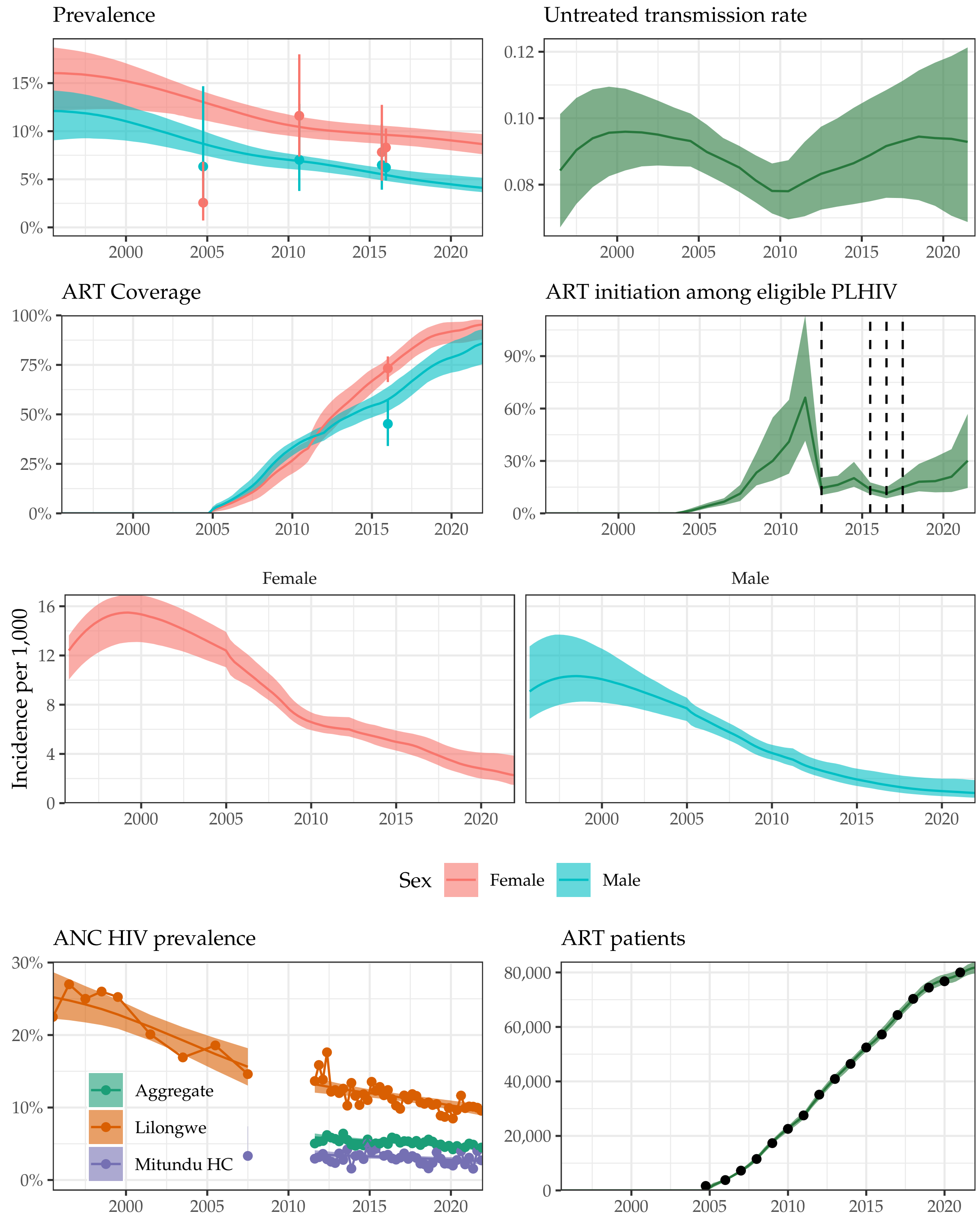

Supplemental Figure 14: **Model fit to HIV data sources in Lilongwe District, 1995-2021.** Estimated prevalence, ART coverage, untreated transmission rates, annual ART initiation probabilities, ANC prevalence, and ART patient counts in the Blantyre district in southern Malawi with household survey data (HIV prevalence and ART coverage), HIV prevalence among pregnant women attending ANC facilities, and the number of adults 15-49 receiving ART programmatic reporting data (points). Prevalence, ART coverage, incidence rate, and ART patients reflect adults aged 15-49 years. Vertical dashed lines indicate years of ART eligibility changes. Different colours on panel "ANC prevalence" indicate different ANC facilities.

#### Mchinji District

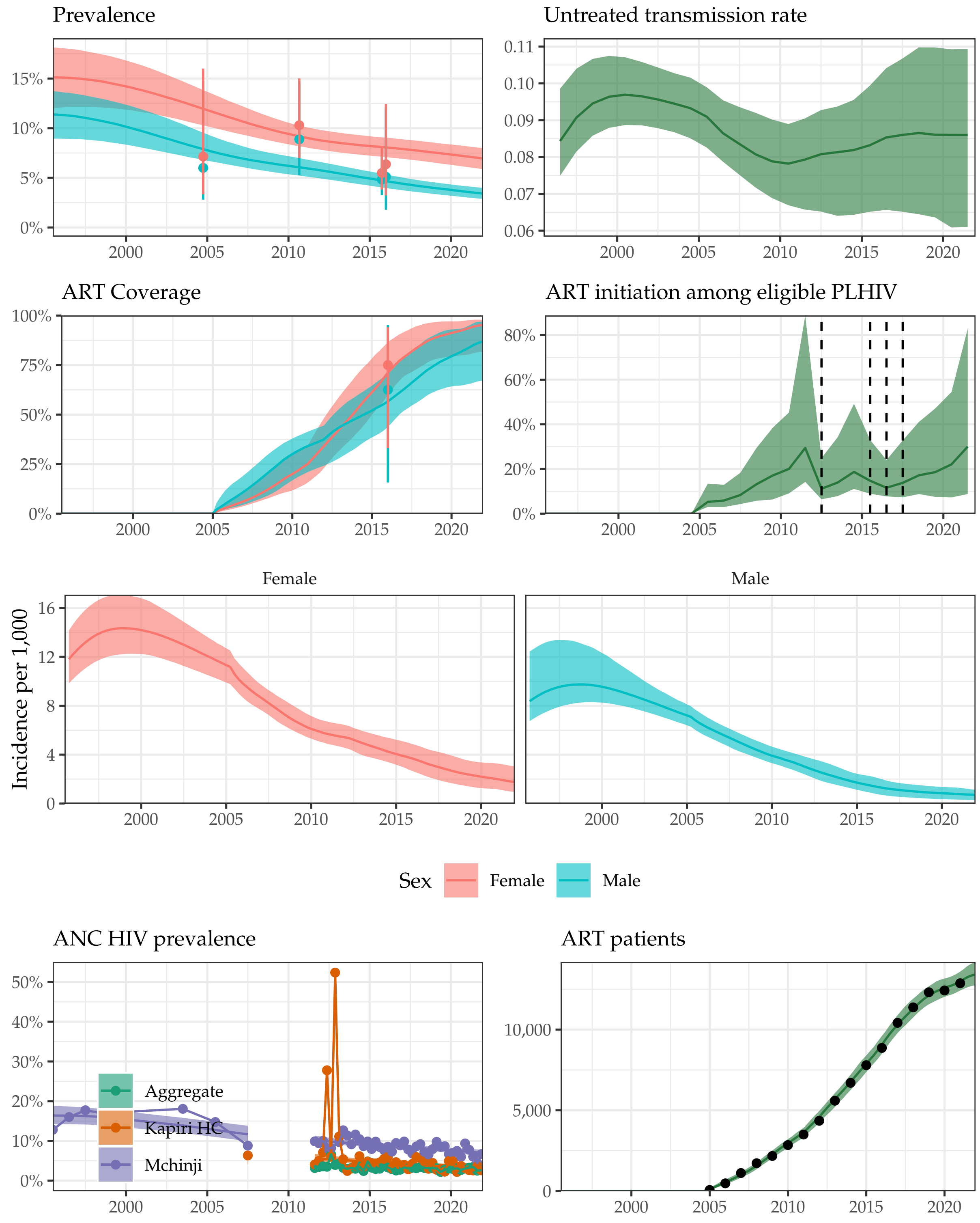

Supplemental Figure 15: **Model fit to HIV data sources in Mchinji District, 1995-2021.** Estimated prevalence, ART coverage, untreated transmission rates, annual ART initiation probabilities, ANC prevalence, and ART patient counts in the Blantyre district in southern Malawi with household survey data (HIV prevalence and ART coverage), HIV prevalence among pregnant women attending ANC facilities, and the number of adults 15-49 receiving ART programmatic reporting data (points). Prevalence, ART coverage, incidence rate, and ART patients reflect adults aged 15-49 years. Vertical dashed lines indicate years of ART eligibility changes. Different colours on panel "ANC prevalence" indicate different ANC facilities.

#### Nkhotakota District

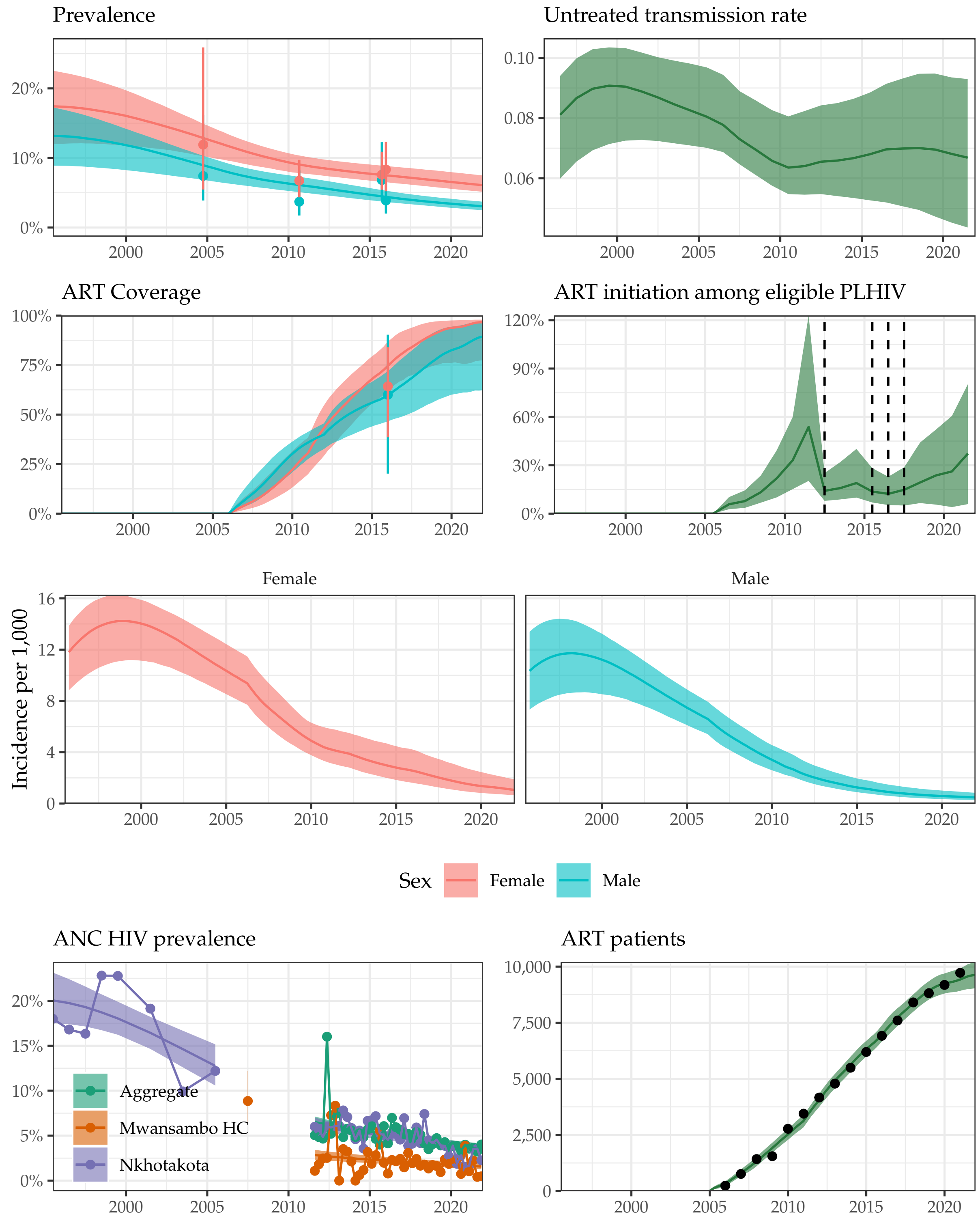

Supplemental Figure 16: **Model fit to HIV data sources in Nkhotakota District, 1995-2021.** Estimated prevalence, ART coverage, untreated transmission rates, annual ART initiation probabilities, ANC prevalence, and ART patient counts in the Blantyre district in southern Malawi with household survey data (HIV prevalence and ART coverage), HIV prevalence among pregnant women attending ANC facilities, and the number of adults 15-49 receiving ART programmatic reporting data (points). Prevalence, ART coverage, incidence rate, and ART patients reflect adults aged 15-49 years. Vertical dashed lines indicate years of ART eligibility changes. Different colours on panel "ANC prevalence" indicate different ANC facilities.

#### Ntcheu District

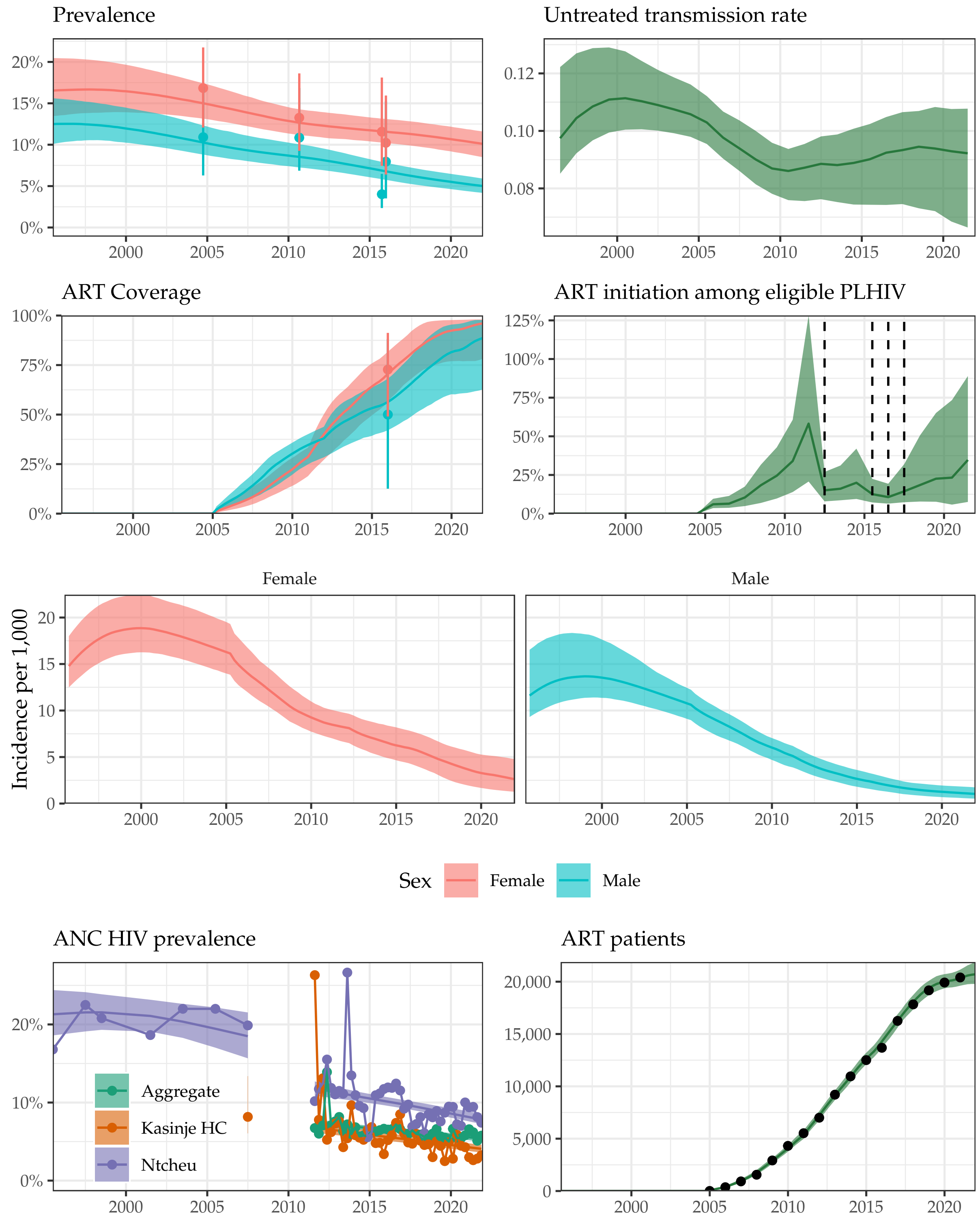

Supplemental Figure 17: **Model fit to HIV data sources in Ntcheu District, 1995-2021.** Estimated prevalence, ART coverage, untreated transmission rates, annual ART initiation probabilities, ANC prevalence, and ART patient counts in the Blantyre district in southern Malawi with household survey data (HIV prevalence and ART coverage), HIV prevalence among pregnant women attending ANC facilities, and the number of adults 15-49 receiving ART programmatic reporting data (points). Prevalence, ART coverage, incidence rate, and ART patients reflect adults aged 15-49 years. Vertical dashed lines indicate years of ART eligibility changes. Different colours on panel "ANC prevalence" indicate different ANC facilities.

#### Ntchisi District

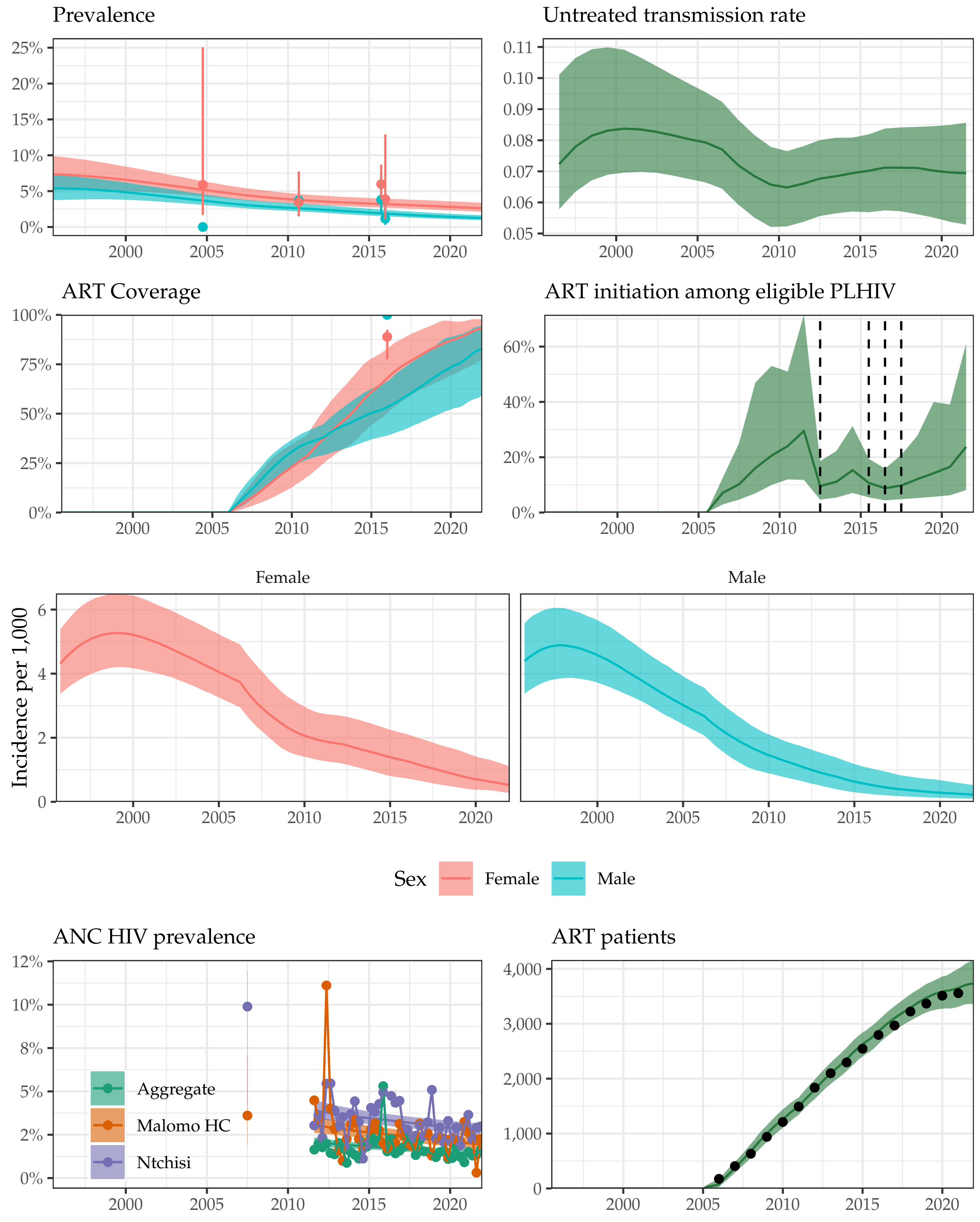

Supplemental Figure 18: **Model fit to HIV data sources in Ntchisi District, 1995-2021.** Estimated prevalence, ART coverage, untreated transmission rates, annual ART initiation probabilities, ANC prevalence, and ART patient counts in the Blantyre district in southern Malawi with household survey data (HIV prevalence and ART coverage), HIV prevalence among pregnant women attending ANC facilities, and the number of adults 15-49 receiving ART programmatic reporting data (points). Prevalence, ART coverage, incidence rate, and ART patients reflect adults aged 15-49 years. Vertical dashed lines indicate years of ART eligibility changes. Different colours on panel "ANC prevalence" indicate different ANC facilities.

#### Salima District

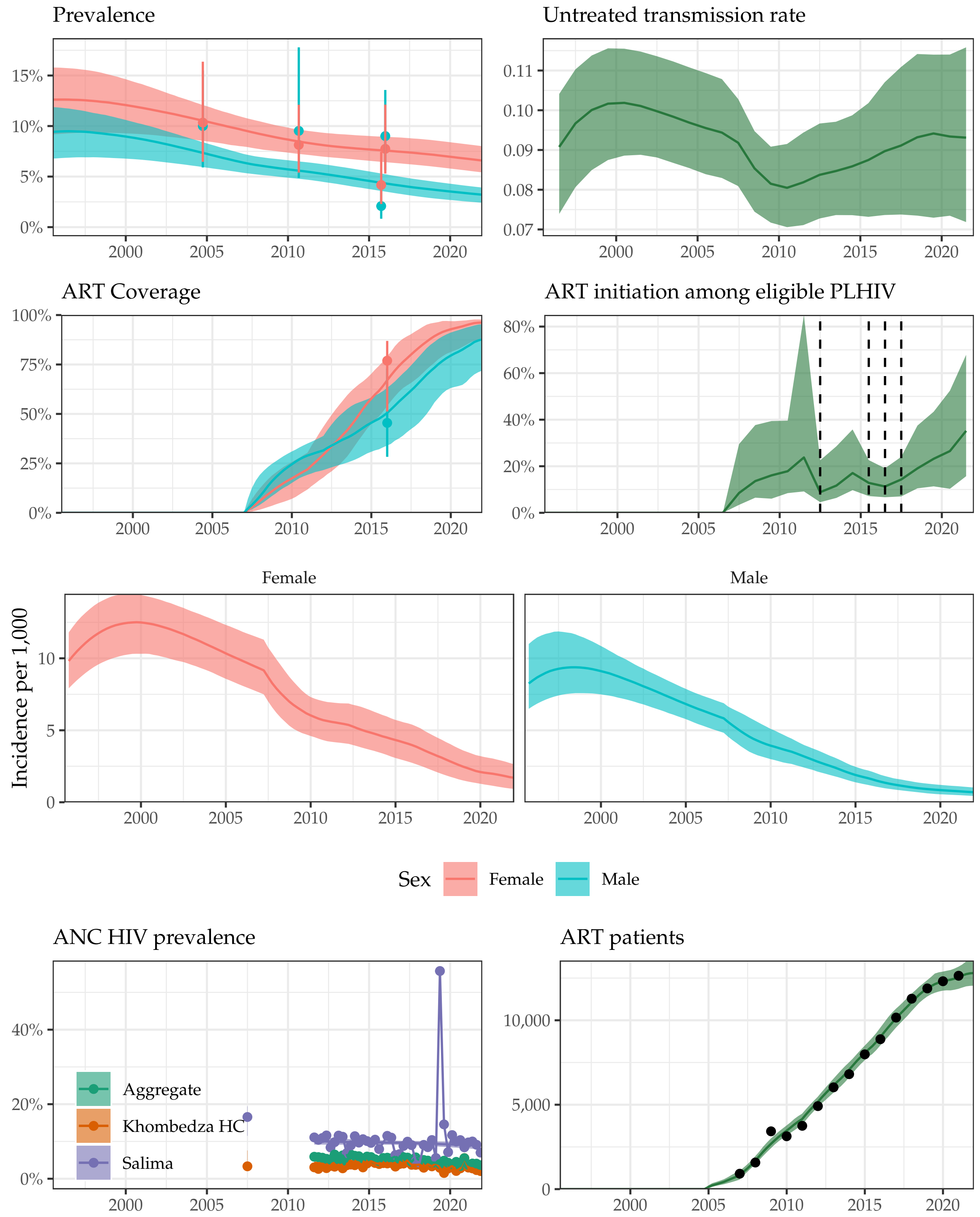

Supplemental Figure 19: **Model fit to HIV data sources in Salima District, 1995-2021.** Estimated prevalence, ART coverage, untreated transmission rates, annual ART initiation probabilities, ANC prevalence, and ART patient counts in the Blantyre district in southern Malawi with household survey data (HIV prevalence and ART coverage), HIV prevalence among pregnant women attending ANC facilities, and the number of adults 15-49 receiving ART programmatic reporting data (points). Prevalence, ART coverage, incidence rate, and ART patients reflect adults aged 15-49 years. Vertical dashed lines indicate years of ART eligibility changes. Different colours on panel "ANC prevalence" indicate different ANC facilities.

#### Balaka District

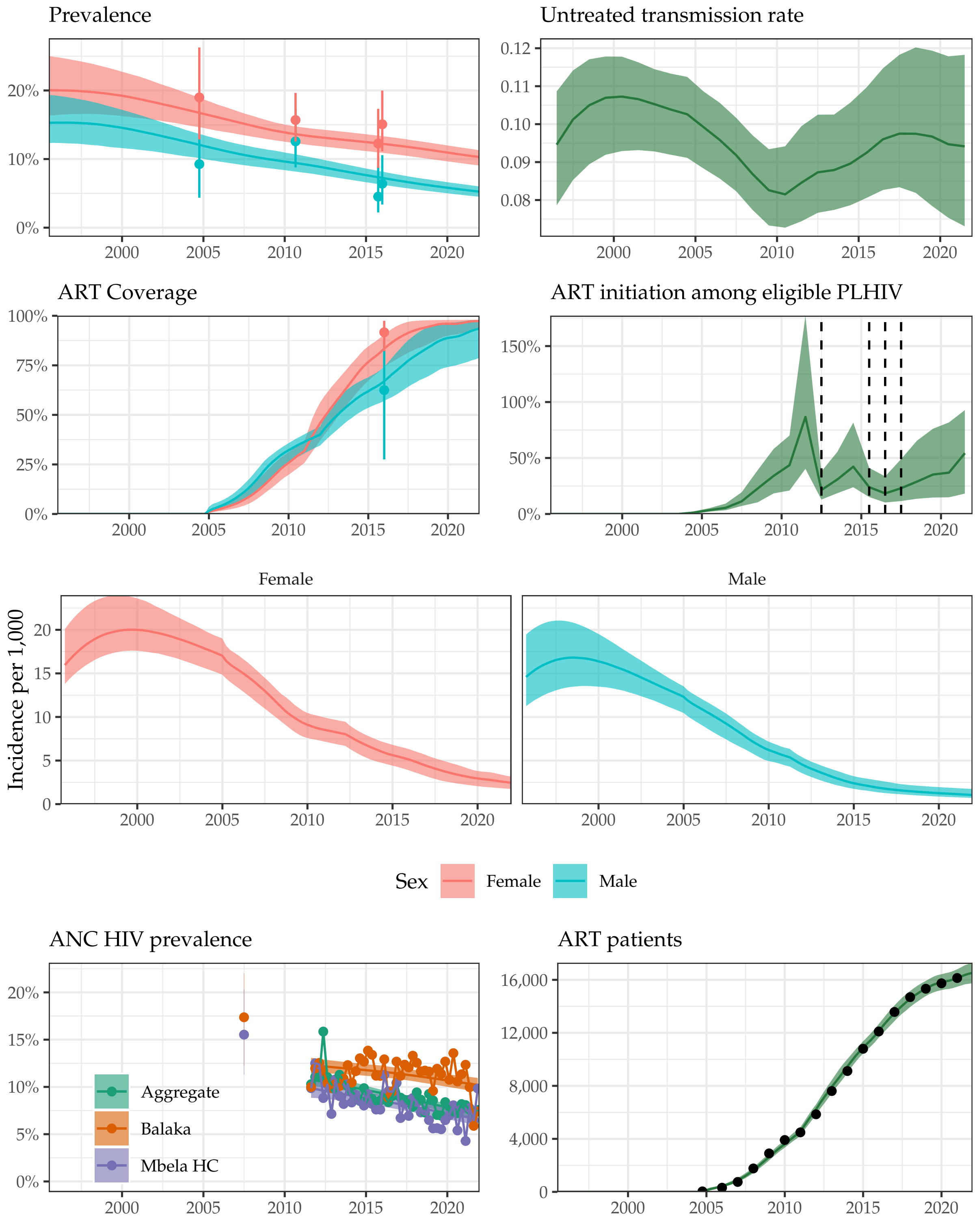

Supplemental Figure 20: **Model fit to HIV data sources in Balaka District, 1995-2021.** Estimated prevalence, ART coverage, untreated transmission rates, annual ART initiation probabilities, ANC prevalence, and ART patient counts in the Blantyre district in southern Malawi with household survey data (HIV prevalence and ART coverage), HIV prevalence among pregnant women attending ANC facilities, and the number of adults 15-49 receiving ART programmatic reporting data (points). Prevalence, ART coverage, incidence rate, and ART patients reflect adults aged 15-49 years. Vertical dashed lines indicate years of ART eligibility changes. Different colours on panel "ANC prevalence" indicate different ANC facilities.

#### Blantyre District

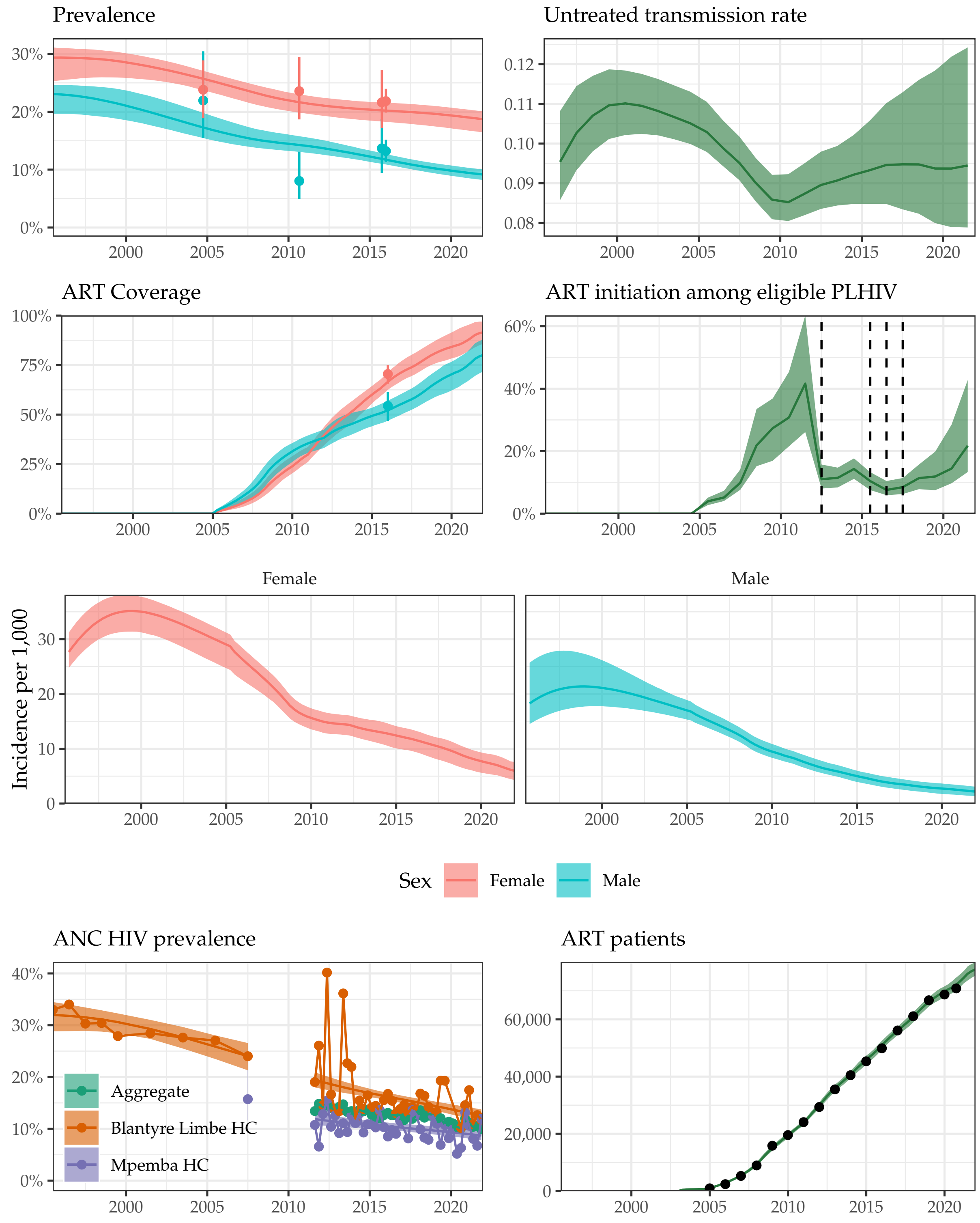

Supplemental Figure 21: **Model fit to HIV data sources in Blantyre District, 1995-2021.** Estimated prevalence, ART coverage, untreated transmission rates, annual ART initiation probabilities, ANC prevalence, and ART patient counts in the Blantyre district in southern Malawi with household survey data (HIV prevalence and ART coverage), HIV prevalence among pregnant women attending ANC facilities, and the number of adults 15-49 receiving ART programmatic reporting data (points). Prevalence, ART coverage, incidence rate, and ART patients reflect adults aged 15-49 years. Vertical dashed lines indicate years of ART eligibility changes. Different colours on panel "ANC prevalence" indicate different ANC facilities.

#### Chikwawa District

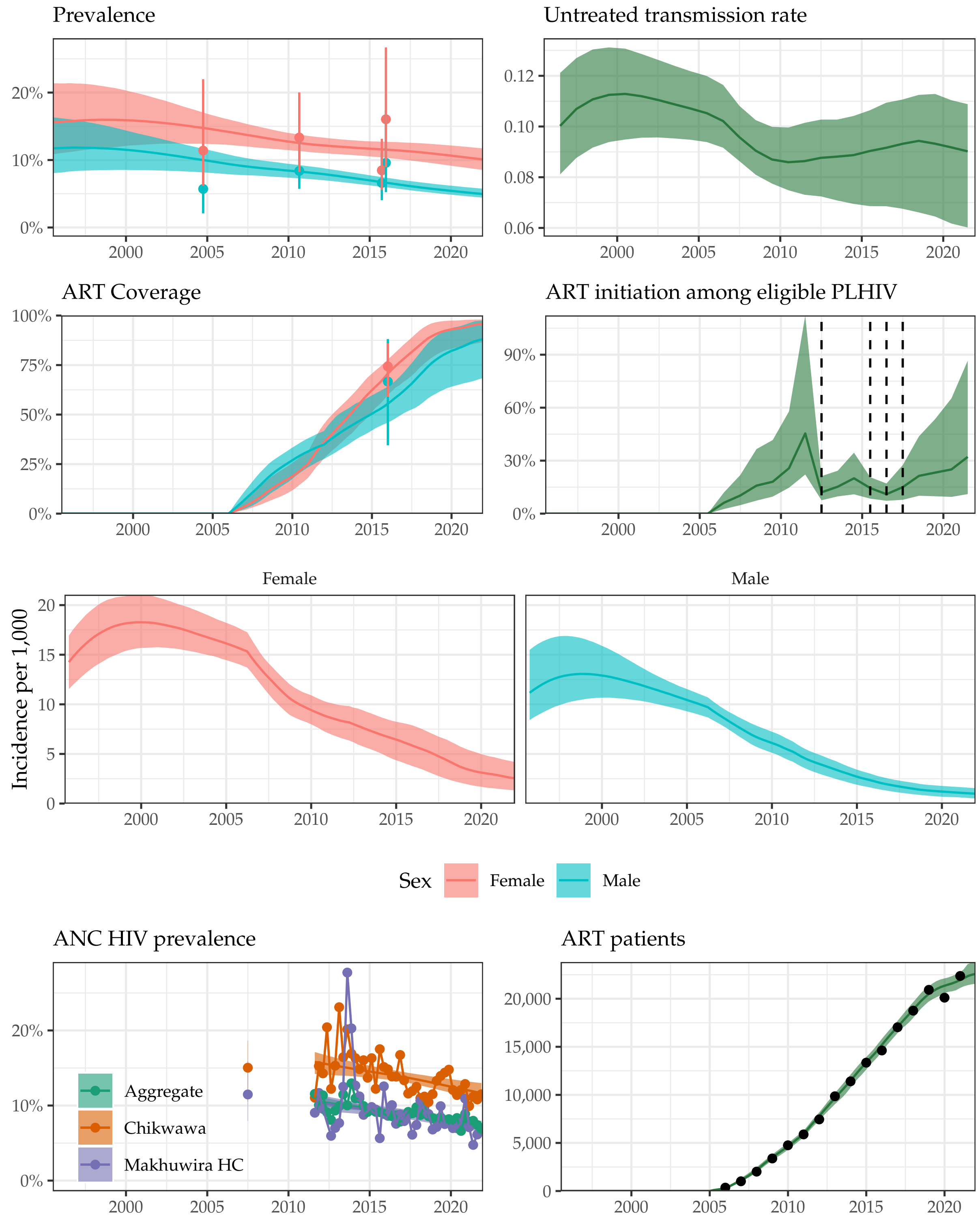

Supplemental Figure 22: **Model fit to HIV data sources in Chikwawa District, 1995-2021.** Estimated prevalence, ART coverage, untreated transmission rates, annual ART initiation probabilities, ANC prevalence, and ART patient counts in the Blantyre district in southern Malawi with household survey data (HIV prevalence and ART coverage), HIV prevalence among pregnant women attending ANC facilities, and the number of adults 15-49 receiving ART programmatic reporting data (points). Prevalence, ART coverage, incidence rate, and ART patients reflect adults aged 15-49 years. Vertical dashed lines indicate years of ART eligibility changes. Different colours on panel "ANC prevalence" indicate different ANC facilities.

#### Chiradzulu District

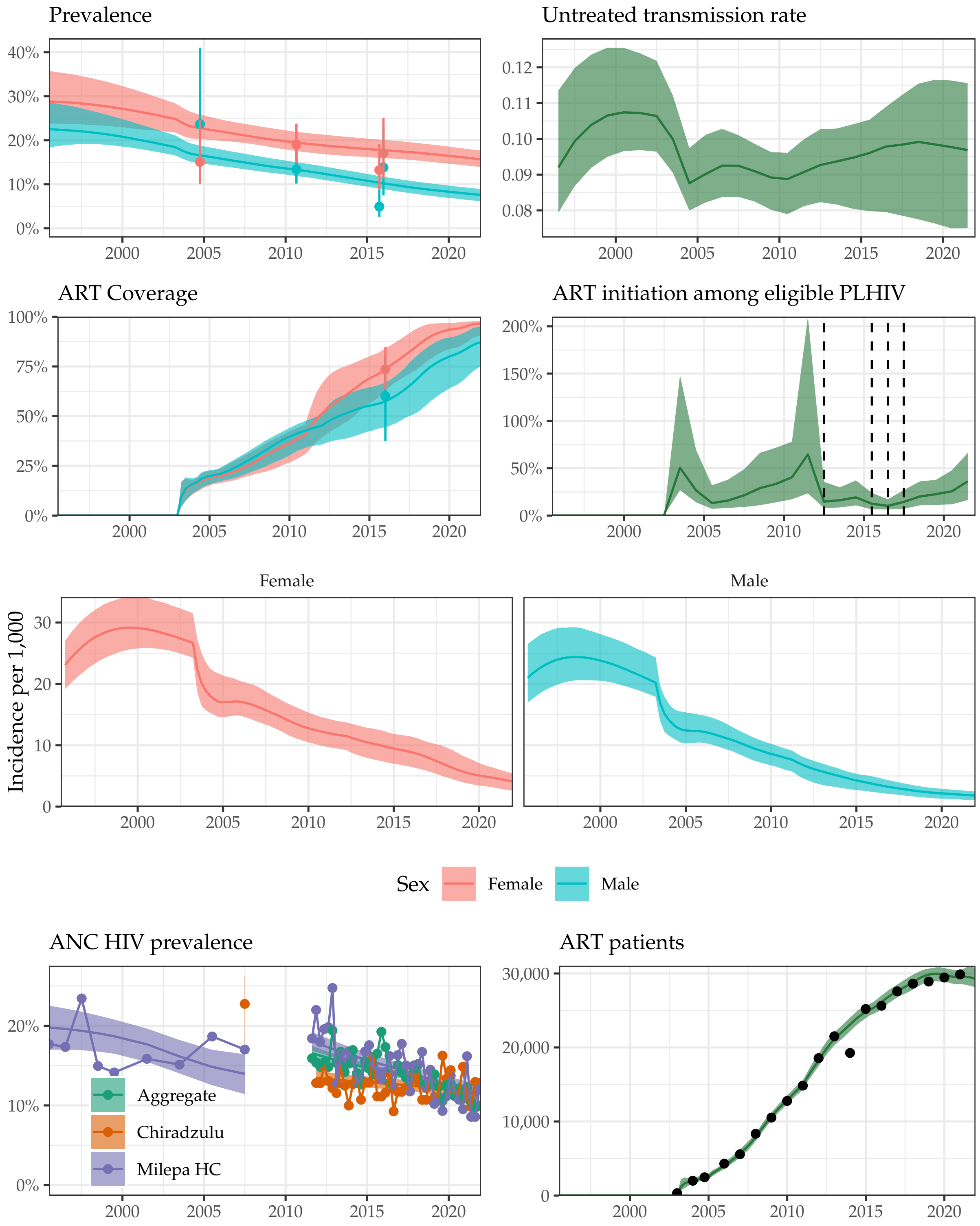

Supplemental Figure 23: **Model fit to HIV data sources in Chiradzulu District, 1995-2021.** Estimated prevalence, ART coverage, untreated transmission rates, annual ART initiation probabilities, ANC prevalence, and ART patient counts in the Blantyre district in southern Malawi with household survey data (HIV prevalence and ART coverage), HIV prevalence among pregnant women attending ANC facilities, and the number of adults 15-49 receiving ART programmatic reporting data (points). Prevalence, ART coverage, incidence rate, and ART patients reflect adults aged 15-49 years. Vertical dashed lines indicate years of ART eligibility changes. Different colours on panel "ANC prevalence" indicate different ANC facilities.

#### Machinga District

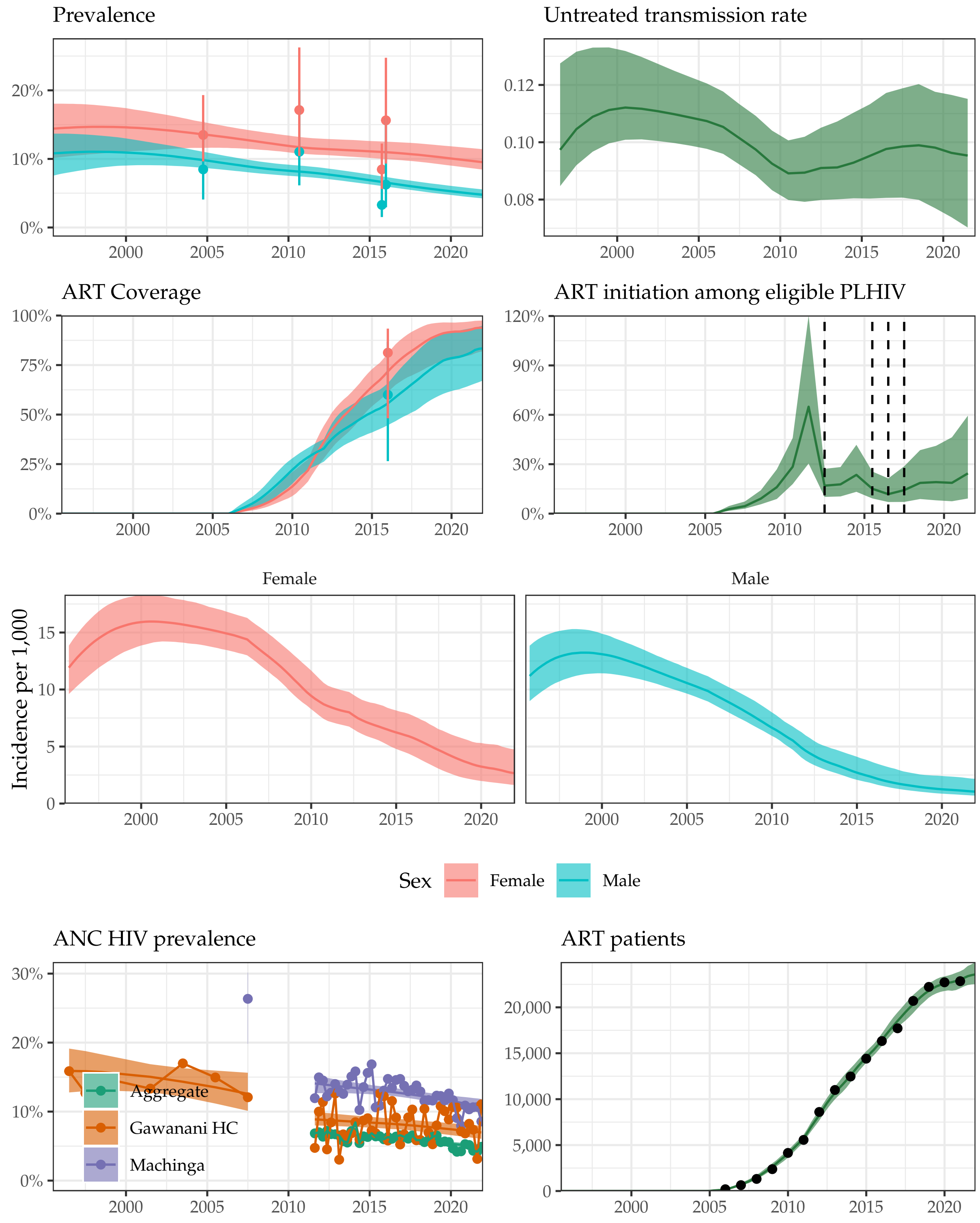

Supplemental Figure 24: **Model fit to HIV data sources in Machinga District, 1995-2021.** Estimated prevalence, ART coverage, untreated transmission rates, annual ART initiation probabilities, ANC prevalence, and ART patient counts in the Blantyre district in southern Malawi with household survey data (HIV prevalence and ART coverage), HIV prevalence among pregnant women attending ANC facilities, and the number of adults 15-49 receiving ART programmatic reporting data (points). Prevalence, ART coverage, incidence rate, and ART patients reflect adults aged 15-49 years. Vertical dashed lines indicate years of ART eligibility changes. Different colours on panel "ANC prevalence" indicate different ANC facilities.

#### Mangochi District

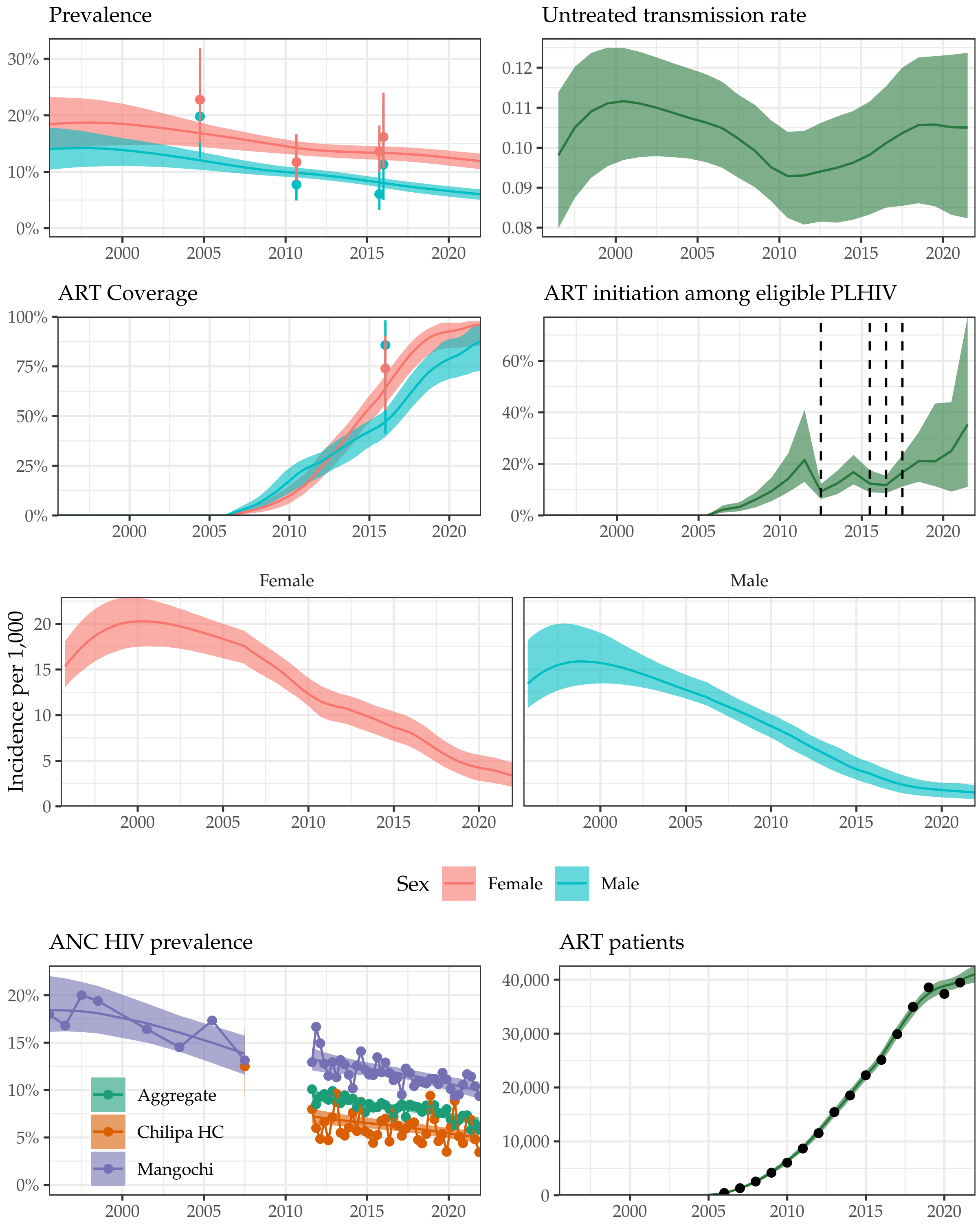

Supplemental Figure 25: **Model fit to HIV data sources in Mangochi District, 1995-2021.** Estimated prevalence, ART coverage, untreated transmission rates, annual ART initiation probabilities, ANC prevalence, and ART patient counts in the Blantyre district in southern Malawi with household survey data (HIV prevalence and ART coverage), HIV prevalence among pregnant women attending ANC facilities, and the number of adults 15-49 receiving ART programmatic reporting data (points). Prevalence, ART coverage, incidence rate, and ART patients reflect adults aged 15-49 years. Vertical dashed lines indicate years of ART eligibility changes. Different colours on panel "ANC prevalence" indicate different ANC facilities.

#### Mulanje District

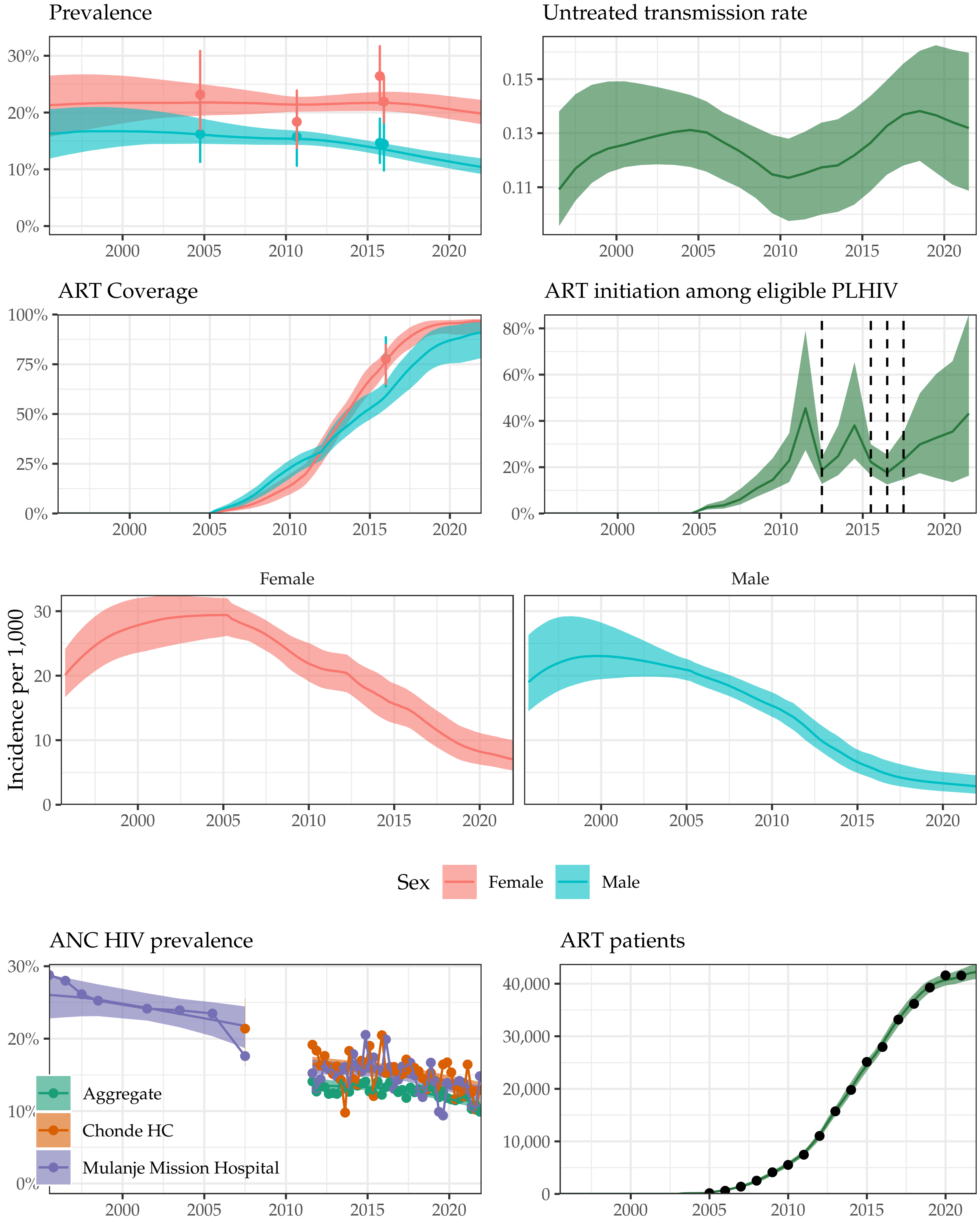

Supplemental Figure 26: **Model fit to HIV data sources in Mulanje District, 1995-2021.** Estimated prevalence, ART coverage, untreated transmission rates, annual ART initiation probabilities, ANC prevalence, and ART patient counts in the Blantyre district in southern Malawi with household survey data (HIV prevalence and ART coverage), HIV prevalence among pregnant women attending ANC facilities, and the number of adults 15-49 receiving ART programmatic reporting data (points). Prevalence, ART coverage, incidence rate, and ART patients reflect adults aged 15-49 years. Vertical dashed lines indicate years of ART eligibility changes. Different colours on panel "ANC prevalence" indicate different ANC facilities.

#### Mwanza District

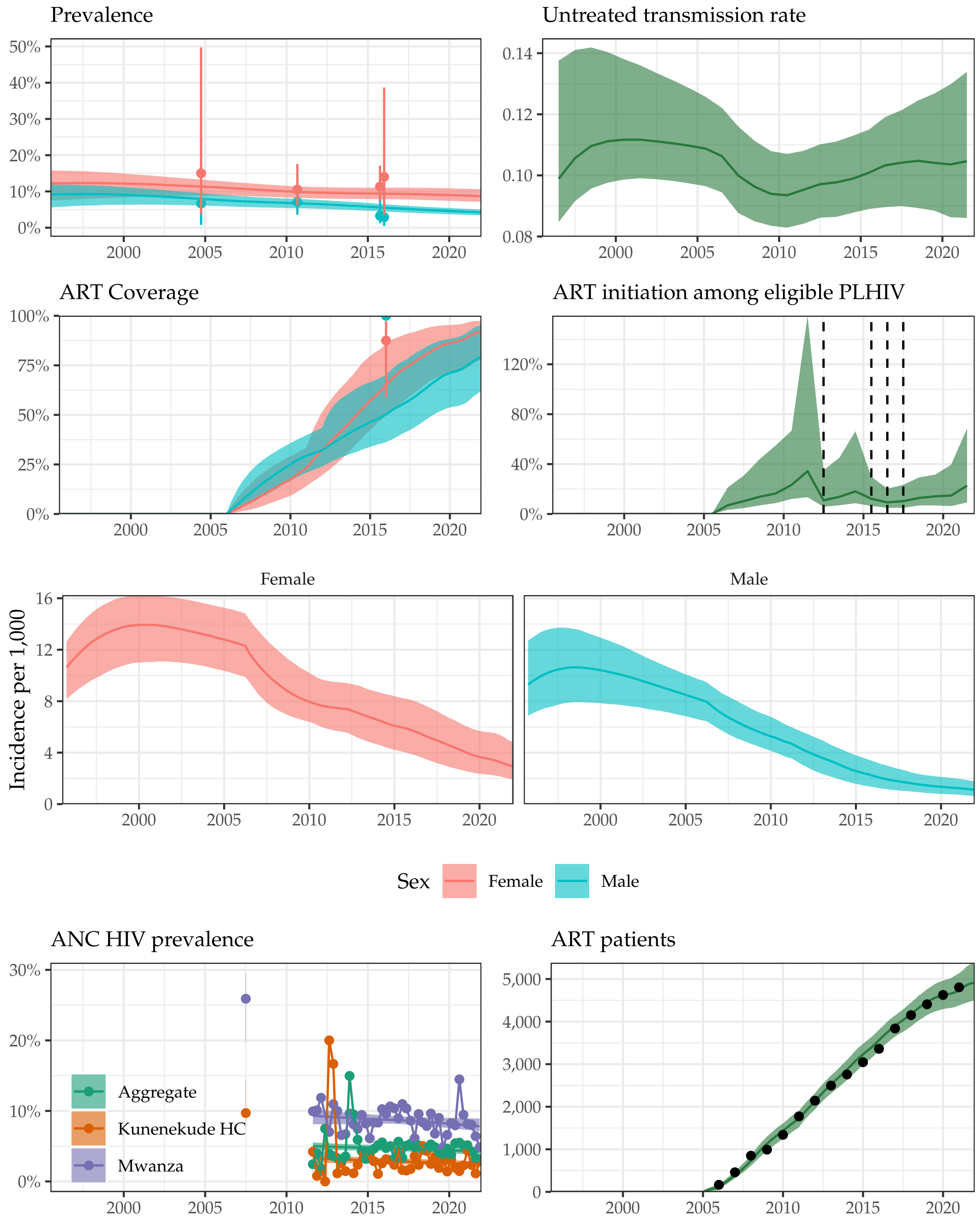

Supplemental Figure 27: **Model fit to HIV data sources in Mwanza District, 1995-2021.** Estimated prevalence, ART coverage, untreated transmission rates, annual ART initiation probabilities, ANC prevalence, and ART patient counts in the Blantyre district in southern Malawi with household survey data (HIV prevalence and ART coverage), HIV prevalence among pregnant women attending ANC facilities, and the number of adults 15-49 receiving ART programmatic reporting data (points). Prevalence, ART coverage, incidence rate, and ART patients reflect adults aged 15-49 years. Vertical dashed lines indicate years of ART eligibility changes. Different colours on panel "ANC prevalence" indicate different ANC facilities.

#### Neno District

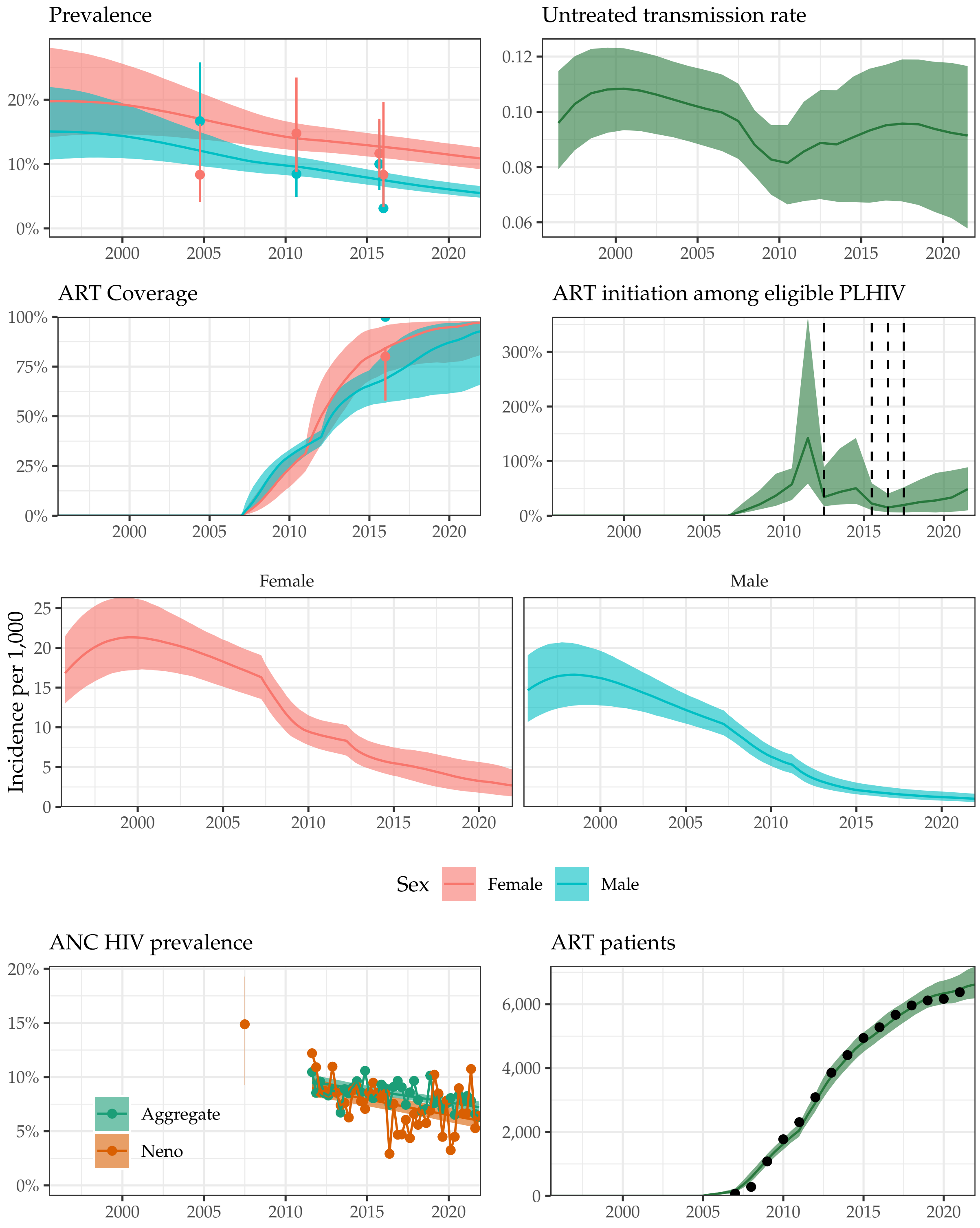

Supplemental Figure 28: **Model fit to HIV data sources in Neno District, 1995-2021.** Estimated prevalence, ART coverage, untreated transmission rates, annual ART initiation probabilities, ANC prevalence, and ART patient counts in the Blantyre district in southern Malawi with household survey data (HIV prevalence and ART coverage), HIV prevalence among pregnant women attending ANC facilities, and the number of adults 15-49 receiving ART programmatic reporting data (points). Prevalence, ART coverage, incidence rate, and ART patients reflect adults aged 15-49 years. Vertical dashed lines indicate years of ART eligibility changes. Different colours on panel "ANC prevalence" indicate different ANC facilities.

#### Nsanje District

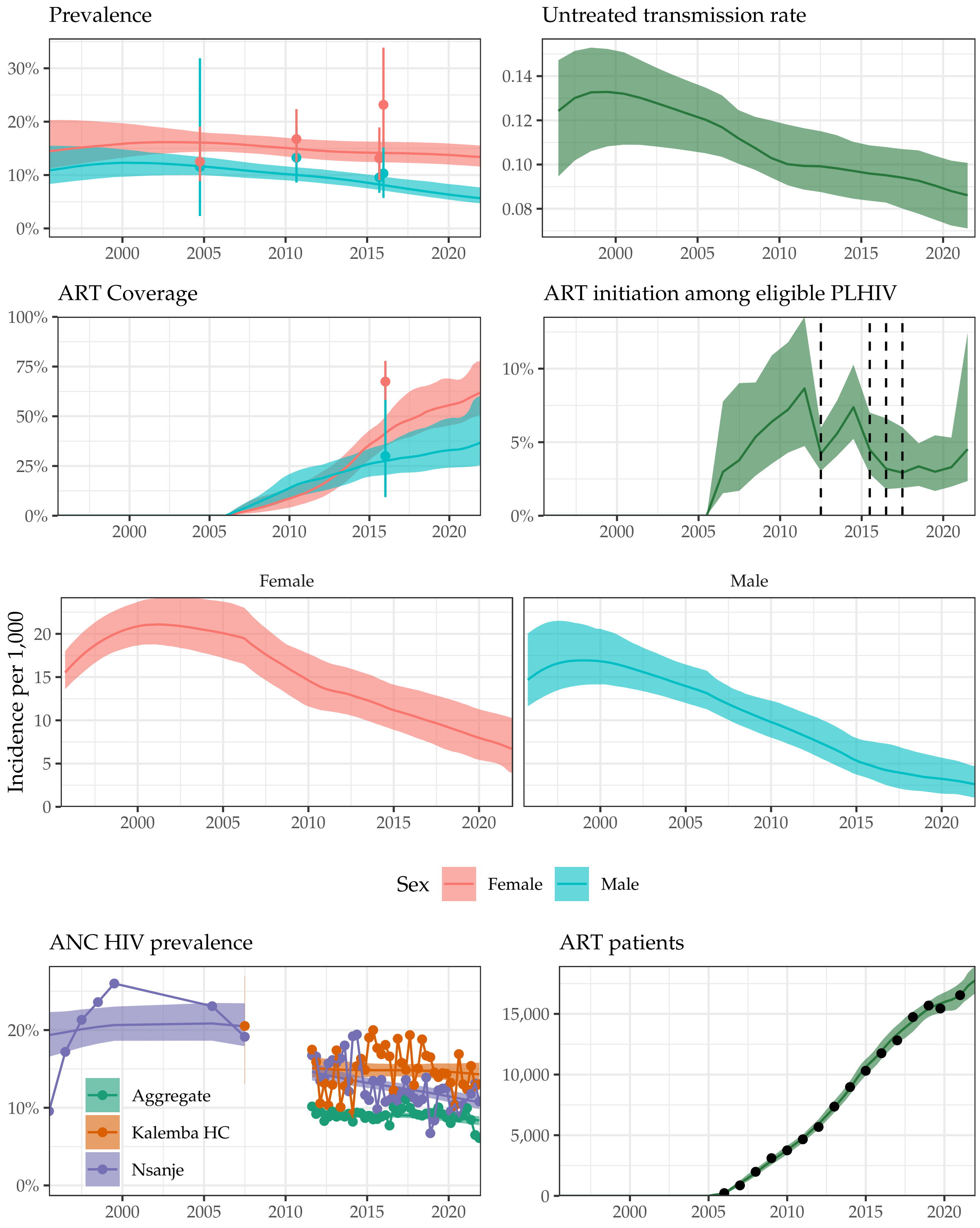

Supplemental Figure 29: **Model fit to HIV data sources in Nsanje District, 1995-2021.** Estimated prevalence, ART coverage, untreated transmission rates, annual ART initiation probabilities, ANC prevalence, and ART patient counts in the Blantyre district in southern Malawi with household survey data (HIV prevalence and ART coverage), HIV prevalence among pregnant women attending ANC facilities, and the number of adults 15-49 receiving ART programmatic reporting data (points). Prevalence, ART coverage, incidence rate, and ART patients reflect adults aged 15-49 years. Vertical dashed lines indicate years of ART eligibility changes. Different colours on panel "ANC prevalence" indicate different ANC facilities.

#### Phalombe District

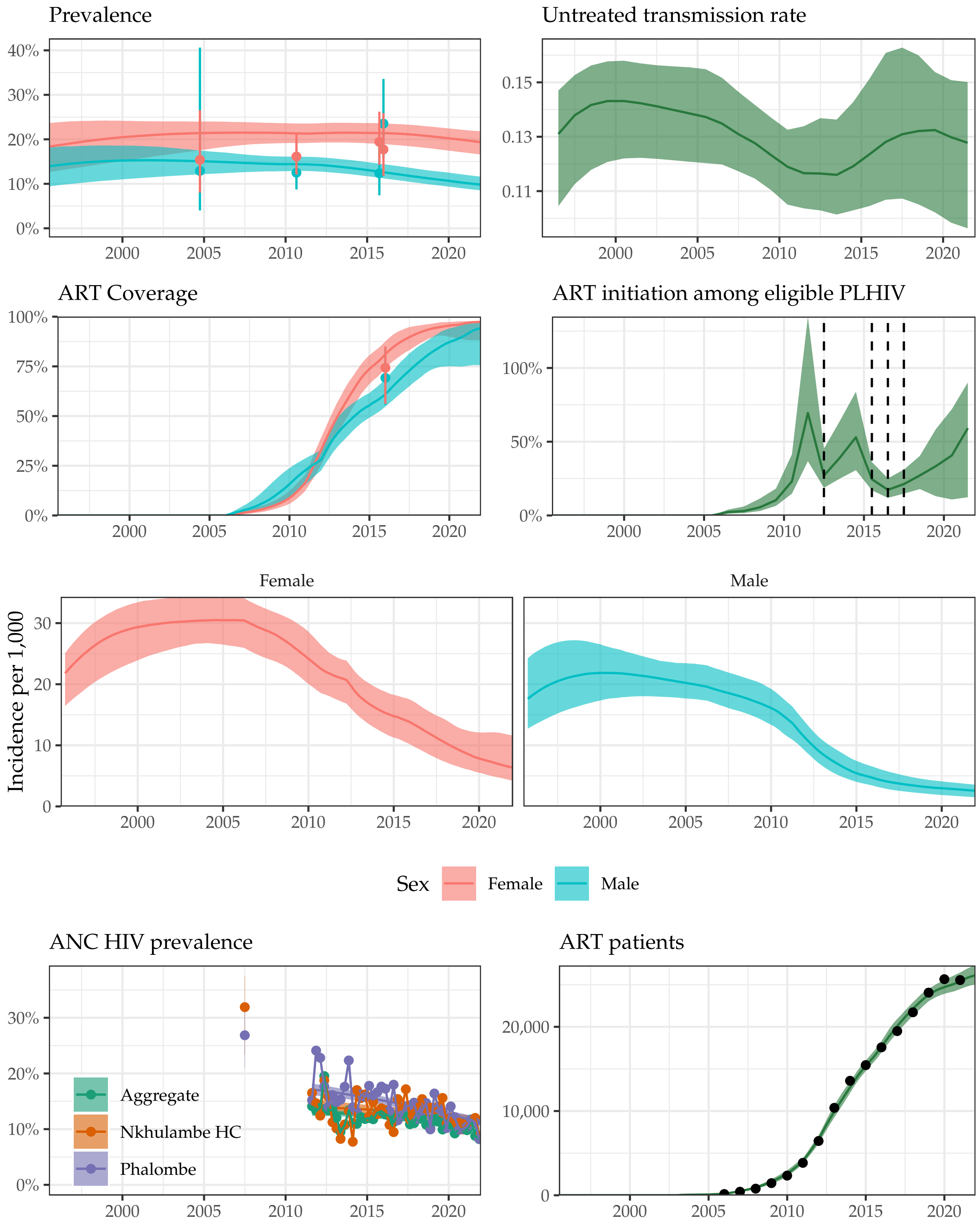

Supplemental Figure 30: **Model fit to HIV data sources in Phalombe District, 1995-2021.** Estimated prevalence, ART coverage, untreated transmission rates, annual ART initiation probabilities, ANC prevalence, and ART patient counts in the Blantyre district in southern Malawi with household survey data (HIV prevalence and ART coverage), HIV prevalence among pregnant women attending ANC facilities, and the number of adults 15-49 receiving ART programmatic reporting data (points). Prevalence, ART coverage, incidence rate, and ART patients reflect adults aged 15-49 years. Vertical dashed lines indicate years of ART eligibility changes. Different colours on panel "ANC prevalence" indicate different ANC facilities.

#### Thyolo District

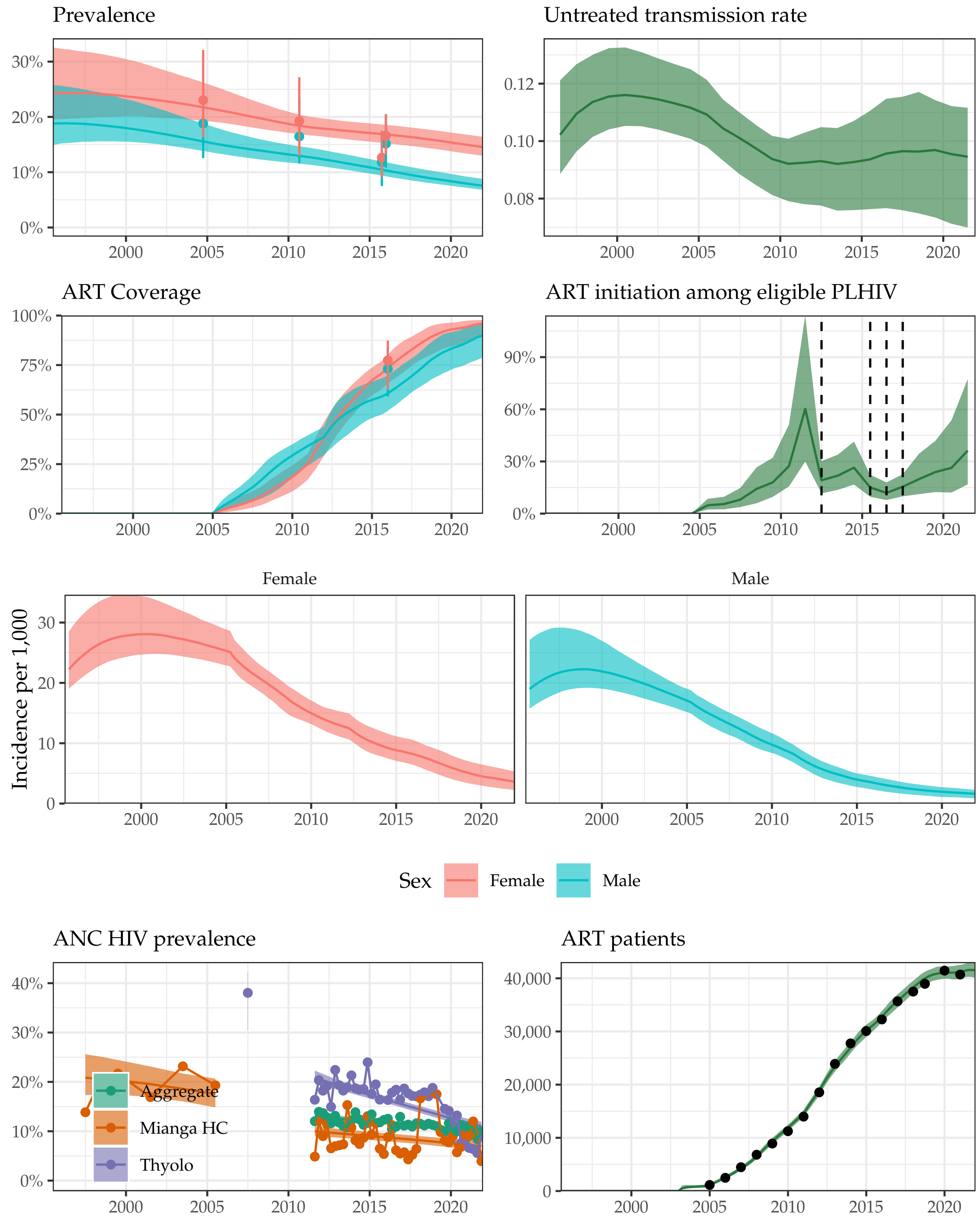

Supplemental Figure 31: **Model fit to HIV data sources in Thyolo District, 1995-2021.** Estimated prevalence, ART coverage, untreated transmission rates, annual ART initiation probabilities, ANC prevalence, and ART patient counts in the Blantyre district in southern Malawi with household survey data (HIV prevalence and ART coverage), HIV prevalence among pregnant women attending ANC facilities, and the number of adults 15-49 receiving ART programmatic reporting data (points). Prevalence, ART coverage, incidence rate, and ART patients reflect adults aged 15-49 years. Vertical dashed lines indicate years of ART eligibility changes. Different colours on panel “ANC prevalence” indicate different ANC facilities.

#### Zomba District

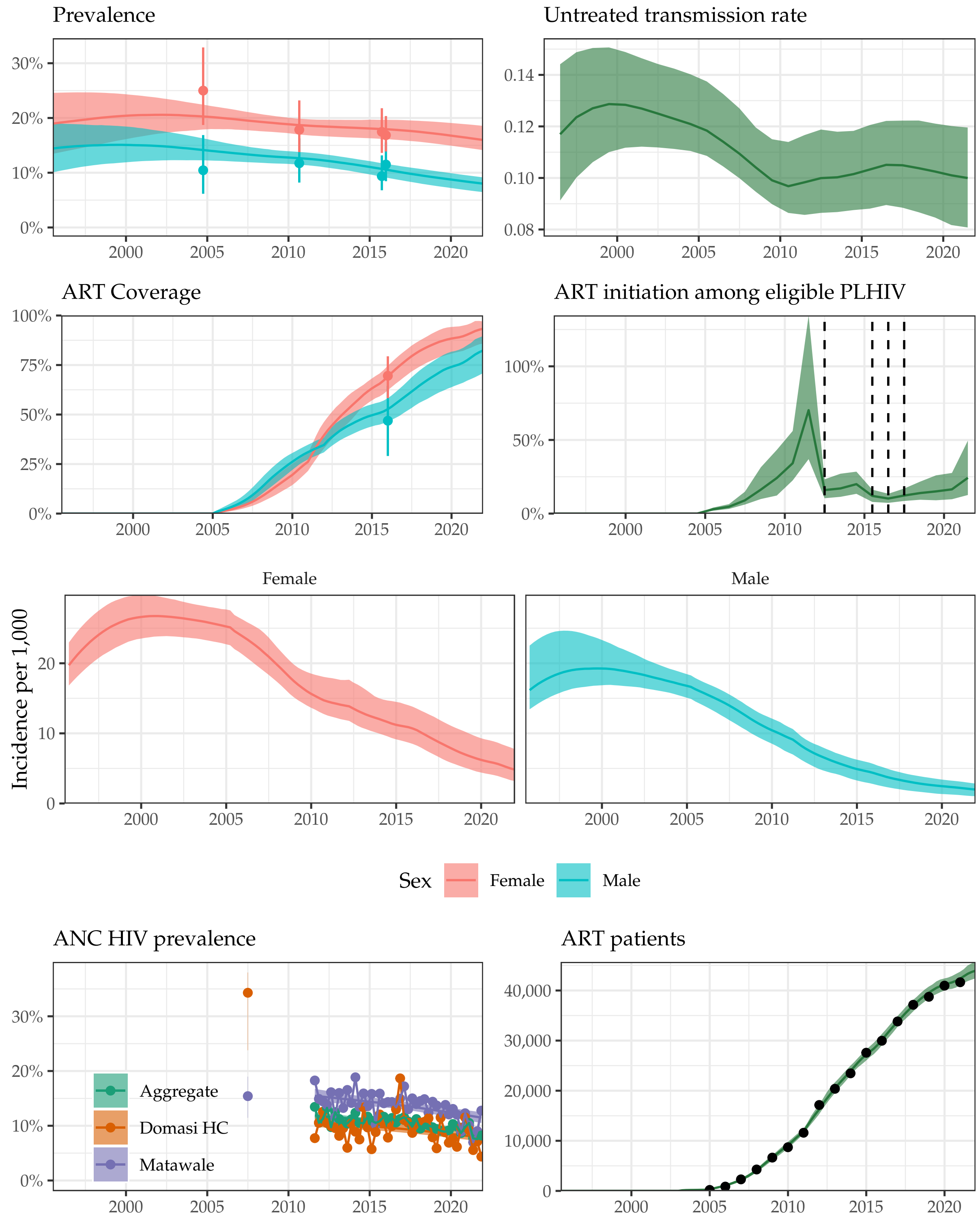

Supplemental Figure 32: **Model fit to HIV data sources in Zomba District, 1995-2021.** Estimated prevalence, ART coverage, untreated transmission rates, annual ART initiation probabilities, ANC prevalence, and ART patient counts in the Blantyre district in southern Malawi with household survey data (HIV prevalence and ART coverage), HIV prevalence among pregnant women attending ANC facilities, and the number of adults 15-49 receiving ART programmatic reporting data (points). Prevalence, ART coverage, incidence rate, and ART patients reflect adults aged 15-49 years. Vertical dashed lines indicate years of ART eligibility changes. Different colours on panel "ANC prevalence" indicate different ANC facilities.

297 # 4 Comparison to UNAIDS 2021 estimates (UNAIDS 2021)

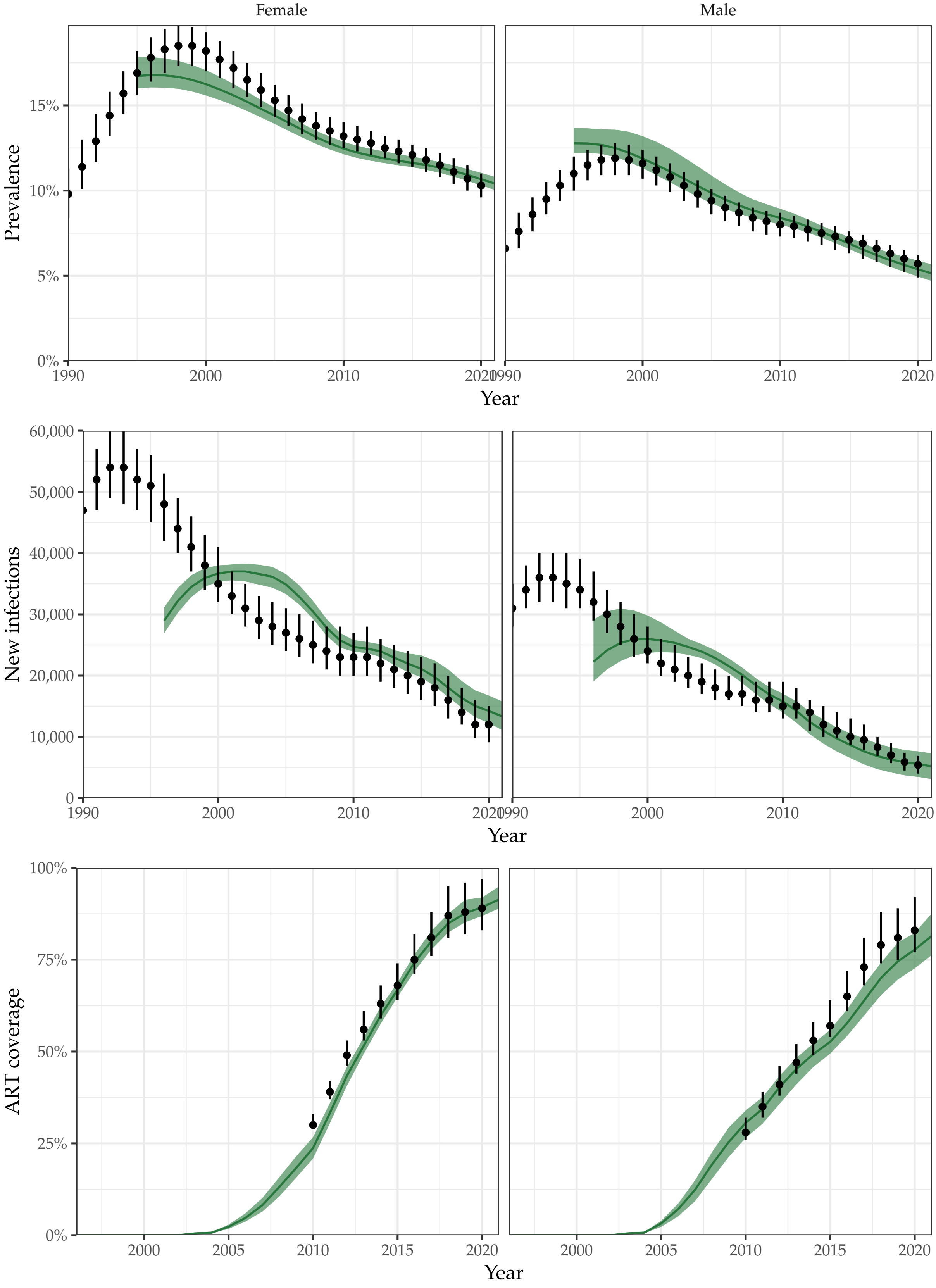

Supplemental Figure 33: Comparison of estimated national-level annual prevalence, new infections, and ART coverage between UNAIDS 2021 estimates (point ranges) and the model presented here (green regions). Note that UNAIDS ART coverage is among all adults.

Hyndman, Rob J., and George Athanasopoulos. 2018. *Forecasting: Principles and Practice*. OTexts.

Johnson, Leigh, and Rob Dorrington. 2019. *Thembisa Version 4.2: A Model for Evaluating the Impact of HIV/AIDS in South Africa*. https://www.thembisa.org/content/downloadPage/Thembisa4_2report.

Kassanjee, Reshma, Thomas A. McWalter, and Alex Welte. 2014. "Short Communication: Defining Optimality of a Test for Recent Infection for HIV Incidence Surveillance." *AIDS Research and Human Retroviruses* 30 (1): 45–49. https://doi.org/10.1089/aid.2013.0113.

Kristensen, Kasper, Anders Nielsen, Casper W. Berg, Hans Skaug, and Brad Bell. 2016. "TMB: Automatic Differentiation and Laplace Approximation." *Journal of Statistical Software* 70 (5). https://doi.org/10.18637/jss.v070.i05.

Lindén, Andreas, and Samu Mäntyniemi. 2011. "Using the Negative Binomial Distribution to Model Overdispersion in Ecological Count Data." *Ecology* 92 (7): 1414–21. https://doi.org/10.1890/10-1831.1.

Risher, Kathryn A, Anne Cori, Georges Reniers, Milly Marston, Clara Calvert, Amelia Crampin,

Tawanda Dadirai, et al. 2021. "Age Patterns of HIV Incidence in Eastern and Southern Africa: A Modelling Analysis of Observational Population-Based Cohort Studies." *The Lancet HIV* 8 (7): e429–39. https://doi.org/10.1016/S2352-3018(21)00069-2.

Skaug, Hans J., and David A. Fournier. 2006. "Automatic Approximation of the Marginal Likelihood in Non-Gaussian Hierarchical Models." *Computational Statistics & Data Analysis* 51 (2): 699–709. https://doi.org/10.1016/j.csda.2006.03.005.

UNAIDS. 2021. "Global HIV & AIDS Statistics — Fact Sheet." 2021. https://www.unaids.org/en/resources/fact-sheet.

Vollmer, Michaela A. C., Ben Glampson, Thomas Mellan, Swapnil Mishra, Luca Mercuri, Ceire Costello, Robert Klaber, Graham Cooke, Seth Flaxman, and Samir Bhatt. 2021. "A Unified Machine Learning Approach to Time Series Forecasting Applied to Demand at Emergency Departments." *BMC Emergency Medicine* 21 (1): 9. https://doi.org/10.1186/s12873-020-00395-y.

Wolock, Timothy M. 2022. "Bayesian Epidemic Models to Infer Spatio-Temporal HIV Incidence with Applications to Malawi." PhD thesis, London, UK: Imperial College London. http://spiral.imperial.ac.uk/handle/10044/1/98107.

Yu, Joseph Kwong-Leung, Solomon Chih-Cheng Chen, Kuo-Yang Wang, Chao-Sung Chang, Simon D. Makombe, Erik J. Schouten, and Anthony D. Harries. 2007. "True Outcomes for Patients on Antiretroviral Therapy Who Are 'Lost to Follow-up' in Malawi." *Bulletin of the World Health Organization* 85 (7): 550. https://doi.org/10.2471/BLT.06.037739.
